# Multi-omics signatures of the human early life exposome

**DOI:** 10.1101/2021.05.04.21256605

**Authors:** Léa Maitre, Mariona Bustamante, Carles Hernández-Ferrer, Denise Thiel, Chung-Ho Lau, Alexandros Siskos, Marta Vives-Usano, Carlos Ruiz-Arenas, Oliver Robinson, Dan Mason, John Wright, Solène Cadiou, Rémy Slama, Barbara Heude, Marta Gallego-Paüls, Maribel Casas, Jordi Sunyer, Eleni Z. Papadopoulou, Kristine B. Gutzkow, Sandra Andrusaityte, Regina Grazuleviciene, Marina Vafeiadi, Leda Chatzi, Amrit K. Sakhi, Cathrine Thomsen, Ibon Tamayo, Mark Nieuwenhuijsen, Jose Urquiza, Eva Borràs, Eduard Sabidó, Inés Quintela, Ángel Carracedo, Xavier Estivill, Muireann Coen, Juan R. González, Hector C. Keun, Martine Vrijheid

**Author notes:** Equally contributed to the work (Léa Maitre, Mariona Bustamante). Equally contributed to the work (Juan R. González, Hector C. Keun, Martine Vrijheid).

## Abstract

Environmental exposures during early life play a critical role in life-course health, yet the molecular phenotypes underlying environmental effects on health are poorly understood. In the Human Early Life Exposome (HELIX) project, a multi-centre cohort of 1,301 mother-child pairs, we associated individual exposomes consisting of >100 chemical, physical and lifestyle exposures assessed in pregnancy and childhood, with multi-omics profiles (methylome, transcriptome, metabolome and proteins) in childhood. We identified 1,170 associations, 249 in pregnancy and 921 in childhood, which revealed potential biological responses and sources of exposure. The methylome best captures the persistent influence of pregnancy exposures, including maternal smoking; while childhood exposures were associated with features from all omics layers, revealing novel signatures for indoor air quality, essential trace elements, endocrine disruptors and weather conditions. This study provides a unique resource (https://helixomics.isglobal.org/) to guide future investigation on the biological effects of the early life exposome.

## Introduction

A large proportion of environmental risk factors remain unknown or poorly defined, although their contribution to disease risk is estimated to be 70–90% (Lim et al., 2012; Rappaport and Smith, 2010). More than a decade ago, the term “exposome” was coined to encompass all environmental factors (i.e. non-genetic factors) to which humans are exposed throughout the life-course (Wild, 2005). Historically, environmental health studies focused almost exclusively on single exposure factors, such as air pollution, lead, or pesticides. The central tenet of the exposome concept is a call for a holistic and systematic approach to assessing the impacts of environment on health. Moreover, the exposome includes not only external exposures, but also the internal biological responses to these exposures through the interrogation of high-dimensional molecular data (Niedzwiecki et al., 2019; Vermeulen et al., 2020; Wild, 2005, 2012).

Of particular interest is the early detection of physiological changes at the molecular level related to environmental exposures before the manifestation of clinical symptoms in healthy populations. Such information may support the biological plausibility of environment-health associations in population studies, help to understand toxicological mechanisms or elucidate how multiple exposures may be grouped based on their common influence on biological pathways (e.g. inflammation) or their source of exposure (e.g. diet), help to identify exposure biomarkers to predict current and past exposures, and will ultimately contribute to preventing environmental health-related disease. Integrative personal omics profiling studies, gathering high-throughput data on multiple molecular layers, have demonstrated that personal molecular profiles may be particularly useful to assess disease risk, detect early preclinical conditions and initiate preventive strategies (Contrepois et al., 2020; Li-Pook-Than and Snyder, 2013; Schüssler-Fiorenza Rose et al., 2019).

Fetal and childhood development has life-long consequences and is critical for many chronic diseases including obesity, cardiometabolic diseases (Franks et al., 2010; Hardy et al., 2015; Juonala et al., 2011), attention-deficit and hyperactivity disorders (ADHD) (Arango et al., 2018) and lung function (Bui et al., 2018). Therefore, early life is a particularly important period to study the early biological triggers of disease: exposures during these developmentally vulnerable periods may have pronounced effects at the molecular level that may remain clinically undetectable until adulthood.

The molecular mechanisms through which early-life environmental exposures may impact birth outcomes and long-term health in humans have primarily been studied through the lens of epigenetics. It is thought that the epigenome orchestrates cellular responses to environmental perturbations and provides cell memory and plasticity (Cavalli and Heard, 2019). The prenatal exposure most studied has been maternal smoking during pregnancy. Offspring of smoker mothers have altered methylation patterns at birth and at older ages (Joubert et al., 2016), which have been linked to later diseases (Bauer et al., 2016) and used to develop epigenetic biomarkers of past exposure (Rauschert et al., 2020; Reese et al., 2017). To a lesser extent, other diverse exposures, from metals to socio-economic factors, have also been linked to differential methylation and are catalogued in public databases (Li et al., 2019)(http://www.ewascatalog.org/). Although epigenetic marks regulate gene transcription and thus the proteome, the relationships between these and the exposome are less studied (Everson and Marsit, 2018). The metabolome, which can reflect physiological responses and microbiome activity as well as the direct internalization of exposures, has received particular attention in exposome research (Athersuch, 2012; Gauglitz et al., 2020; Niedzwiecki et al., 2019; Rappaport et al., 2014). However, there is still a lack of large-scale studies where multi-omics data have been contextualised and integrated to study environmental health in early life. In this work, we aimed to associate the personal early life exposome, measured in 1,301 mother-child pairs of the Human Early Life Exposome (HELIX) project, with deep molecular phenotype data assessed in childhood and defined by the blood methylome and transcriptome, plasma proteins, and serum and urinary metabolites (Maitre et al., 2018a). By systematically documenting all associations between the exposome and the molecular phenotypes, we provide a unique resource (https://helixomics.isglobal.org/) for the identification of novel exposure biomarkers and early biological effects during developmentally vulnerable life periods.

## Results

### Building the early life exposome and measuring multi-omics phenotypes in children

We assessed a broad spectrum of environmental exposures in 1,301 mother–child pairs from the HELIX project, a multi-centre longitudinal population-based cohort study in 6 locations in Europe (Spain, UK, France, Lithuania, Norway and Greece) **(Figure 1A; Supplemental Experimental Procedures)** (Maitre et al., 2018a).

**Figure 1.**
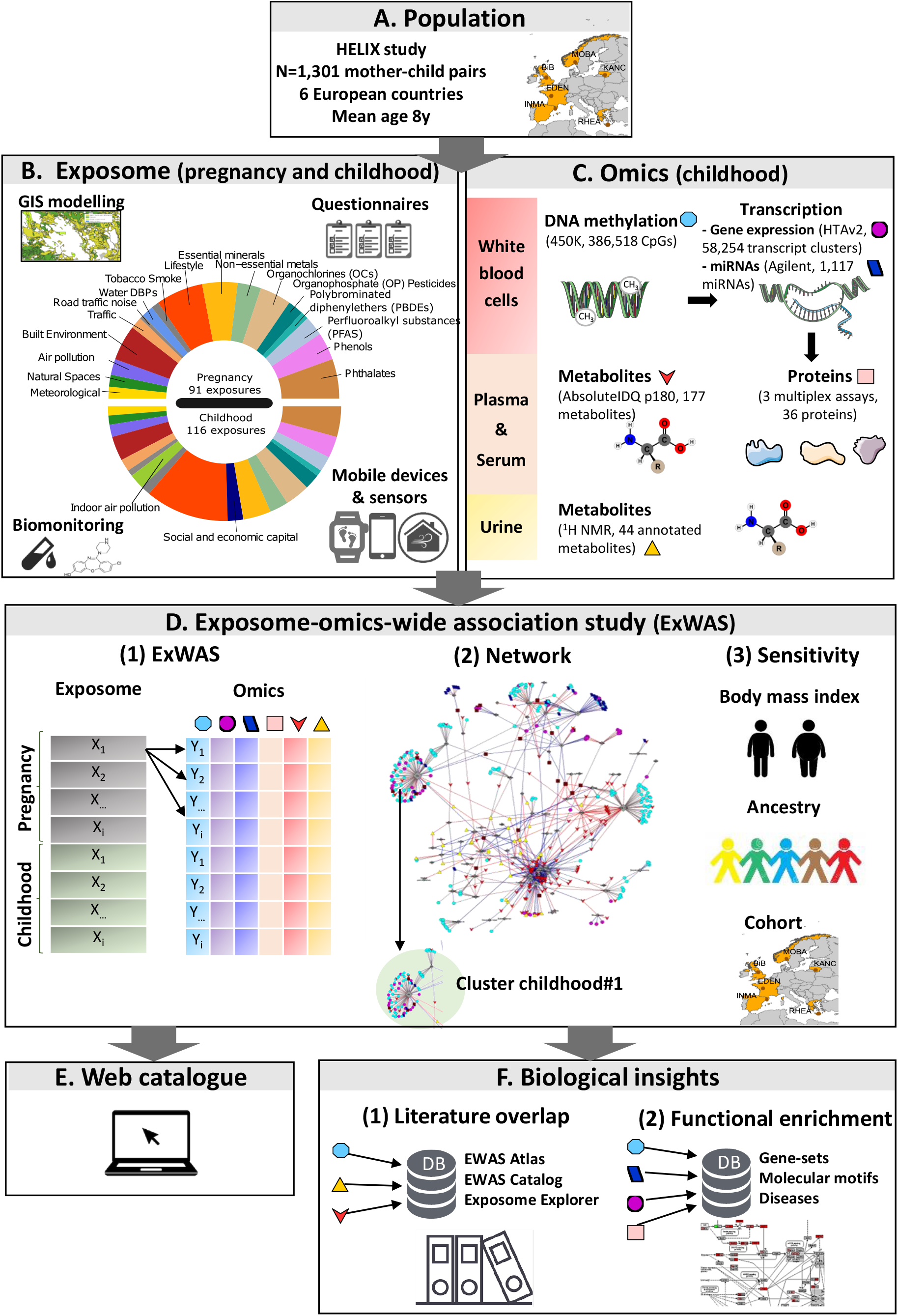
An overview of the early life exposome and multi-omics signature study. **A)** The birth cohorts participating in the study. **B)** The methods used to measure the exposome in pregnancy and childhood. The pie chart represents the exposure families and their proportion corresponds to the number of different exposures measured. **C)** The methods used to measure child molecular phenotypes. Six different platforms were used in three biological matrices. **D)** An exposome-omics-wide association study (ExWAS) was applied, modelling exposure-omics one by one (1). These associations were visualized through network and clusters were identified as depicted in (2). Sensitivity analyses were undertaken running ExWAS again without adjusting for child BMI, only in European ancestry children and by cohorts (3). **E)** All summarized results can be found in https://helixomics.isglobal.org/. **F)** Exposure-omics associations and clusters were interpreted through literature overlap (1) and functional enrichment analyses (2).

Exposure assessment tools included mass-spectrometry based measurement of biomarkers of chemical exposure in urine and blood, exposure monitors, remote sensing and geospatial methods, and self-reports to trained interviewers **(Figure 1B; Supplemental Experimental Procedures)**. The early life exposome consisted of 91 environmental exposure in pregnancy and 116 in childhood, covering 18 exposure families: meteorological factors, proximity to natural spaces, indoor and outdoor air pollution, built environment, road traffic, noise, water disinfection by-products, tobacco smoking, lifestyle factors (diet, physical activity), social and economic capital, essential minerals and chemical pollutants (non-essential metals, organochlorines, organophosphate pesticides, polybrominated diphenylethers, perfluoralkyl substances, phenols and phthalates). Exposure levels and correlation patterns between exposure variables in the HELIX cohort are described further elsewhere (Haug et al., 2018; Robinson et al., 2018; Tamayo-Uria et al., 2019).

For these same children, we performed in-depth multi-omics molecular phenotyping, including measurement of blood DNA methylation (450K, Illumina), blood gene expression (HTA v2.0, Affymetrix), blood miRNA expression (SurePrint Human miRNA rel 21, Agilent), plasma proteins (Luminex), serum metabolites (AbsoluteIDQ p180 kit, Biocrates), and urinary metabolites (^1^H NMR spectroscopy) **(Figure 1C; Supplemental Experimental Procedures)**. Around 91% of the children had molecular data from at least 4 of the omics platforms. While blood DNA methylation and transcriptomics were measured genome-wide, the other omics followed a semi-targeted or targeted approach. Details of the plasma proteins and the urinary and serum metabolomic analyses can be found elsewhere (Lau et al., 2018; Vives-Usano et al., 2020).

### The methylome best captures the persistent influence of pregnancy exposures, while childhood exposures are associated with features from all omics layers

We first systematically tested the association between each exposure variable and each molecular feature (Exposome-omics-wide association analysis, ExWAS) by fitting linear regression models adjusted for cohort, child’s age, sex, zBMI, ancestry, maternal education and omics specific covariates **(Figure 1D; Supplemental Experimental Procedures)**. Out of ∼10 M tested associations, 1,170 were statistically significant after multiple-testing correction considering the number of molecular features within each omics platform (see **Supplemental Experimental Procedures; Table S1**). Significant associations are displayed by family of exposure in Miami plots in **Figure 2A1 and 2B1**. All results can be viewed in the HELIX-exp-omics web catalogue: https://helixomics.isglobal.org/ (for genome-wide assays only results with p-values <0.01 were included).

**Figure 2.**
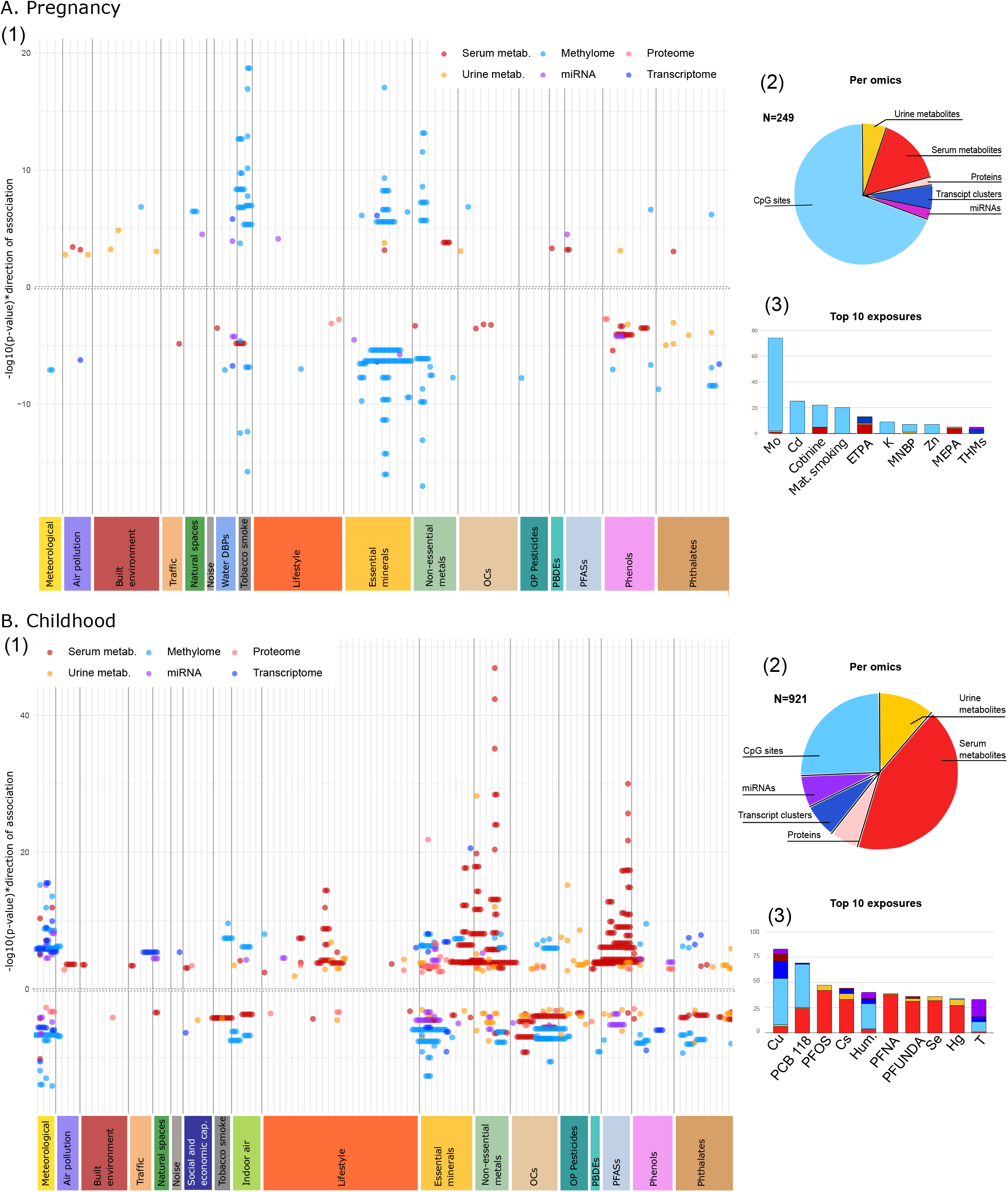
Exposome-omics-wide association study (ExWAS) for the pregnancy and childhood exposome. **A)** Summary of the pregnancy exposome-omics associations: Miami plot (1); pie charts showing the proportion of associations with the different molecular layers (2); and top 10 pregnancy exposures (3). **B)** Summary of the childhood exposome-omics associations: Miami plot (1); pie charts showing the proportion of associations with the different molecular layers (2); and top 10 childhood exposures (3). In Miami plots, each point corresponds to an exposure-omics association; the y-axes show the - log10p-values multiplied by the direction of the association (sign of the regression coefficient); and the x-axis groups exposures along the 19 exposure families. In the pie-chart and histogram, colours indicate the molecular layer.

Associations between the pregnancy exposome and molecular phenotypes totalled 249, including 52 unique exposures and 209 unique molecular features, while the 921 associations with the childhood exposome corresponded to 84 unique exposures and 454 unique molecular features. The pregnancy exposome was predominantly associated with child DNA methylation (70% of associations observed) **(Figure 2A2)**; in contrast, the childhood exposome was associated with all molecular layers, with the serum metabolome showing the highest number of associations (43% of associations observed) **(Figure 2B2)**. The top 10 exposures in terms of numbers of associations included essential elements, heavy metals, tobacco smoking, parabens, and phthalates for the pregnancy exposome **(Figure 2A3)**, and essential elements, heavy metals, persistent pollutants (PFASs and PCBs) and meteorological factors for the childhood exposome **(Figure 2B3)**. We observed fewer omics associations for outdoor air pollution, built environment, road traffic, noise and water disinfection by-products **(Figure 2)**.

#### Exposome-omics networks reveal distinct exposure clusters and the different nature of their underlying molecular signatures

Given the large number of significant associations, we visualized the results in two networks for the pregnancy and childhood exposomes **(Figures 3 and 4; Supplemental Experimental Procedures)**. In these networks, an exposure and a molecular feature were connected (N_nodes_=538, either molecular features or exposures) if their association was statistically significant (N_edges_=1,170) and only connected components with at least two molecular features were displayed. The pregnancy exposome network was mostly composed of CpGs and very disconnected. It contained 3 main connected components (referred to as clusters named as “preg#…”), the largest of which contained less than 30% of all nodes **(Figure 3; Table S2)**. This lack of connectivity can be explained by the wide-spacing along the genome of the CpG sites captured with the 450K array and their low correlation. These 3 clusters varied greatly by their size, their number of exposures and the type of omics composing them **(Table 1)**.

**Table 1.** Pregnancy and childhood exposome-omics clusters

| Exposome | Cluster | Name | Exposures, ordered by degree | N exposures | N omics | N methy | N trans | N miRNA | N prot | N met_s | N met_u | N ann. gene |
| --- | --- | --- | --- | --- | --- | --- | --- | --- | --- | --- | --- | --- |
| Pregnancy | Preg#1 | Tobacco smoke | Cd, Cotinine, Mat. Smoking, Hg, PFUNDA | 5 | 48 | 39 | 0 | 1 | 0 | 8 | 0 | 19 |
|  | Preg#2 | Molybdenum (Mo) | Mo | 1 | 74 | 72 | 0 | 0 | 0 | 1 | 1 | 47 |
|  | Preg#3 | Phtalates and parabens | ETPA, MEPA, PRPA, MEHHP, BUPA, MECPP, MEOHP | 7 | 20 | 1 | 0 | 5 | 0 | 10 | 4 | 6 |
| Childhood (postnatal) | Childhood#1 | Organochlorine chemicals | PCB 118, HCB, PCB 138, PCB 153, PM2.5, PCB 180, House crowding, PCB 170, Bread, DDE, Diet fat, MEP, MIBP | 13 | 74 | 43 | 1 | 1 | 3 | 24 | 2 | 41 |
|  | Childhood#2 | Copper (Cu) | Cu, Pb, land use | 3 | 89 | 52 | 5 | 17 | 6 | 8 | 1 | 69 |
|  | Childhood#3 | Fish and contaminants | PFOS, Cs, PFNA, Se, PFUNDA, Hg, As, Fish, PFHXS, Cotinine, Dairy, PBDE 153, Fastfood, ETS, BTEX in, Sweets | 16 | 71 | 2 | 0 | 3 | 2 | 56 | 8 | 5 |
|  | Childhood#4 | Weather | Hum., T, UV, K, PFOA, Mg, BPA, TI, PM10 | 9 | 78 | 30 | 21 | 18 | 5 | 4 | 0 | 65 |
|  | Childhood#5 | Phthalates (DEHP metabolites) | MEOHP, MEHHP, MECPP, MEHP, Bakery prod | 5 | 16 | 8 | 1 | 0 | 0 | 7 | 0 | 10 |
|  | Childhood#6 | Non-persistent chemicals and diet | OXOMINP, KIDMED, DMTP, Co, BUPA, DETP, OHMiNP, Cd, Fruits, DMDTP, DEP, Vegetables, Meat, Cereals, Social participation, DMP | 16 | 38 | 13 | 0 | 1 | 2 | 1 | 21 | 15 |
|  | Childhood#7 | Indoor air | PM2.5 in, Smoking, Blue space, MNBP | 4 | 22 | 13 | 0 | 4 | 0 | 5 | 0 | 12 |
|  | Childhood#8 | Manganese (Mn) and Molybdenum (Mo) | Mn, Mo | 2 | 10 | 1 | 6 | 0 | 1 | 2 | 0 | 8 |
|  | Childhood#9 | Benzene | Benzene in | 1 | 9 | 9 | 0 | 0 | 0 | 0 | 0 | 7 |
|  | **Childhood#10** | Triclozan | TRCS | 1 | 5 | 0 | 0 | 2 | 3 | 0 | 0 | 5 |
|  | **Childhood#11** | Distance road | Distance road | 1 | 2 | 0 | 0 | 0 | 0 | 2 | 0 | 0 |

Abbreviations: methy = DNA methylation (CpGs); trans = gene expression (TCs); miRNA = miRNA expression; prot = plasma proteins; met\_s = serum metabolites; met\_u = urine metabolites; N ann. gene = number of genes annotated to molecular features

**Figure 3.**
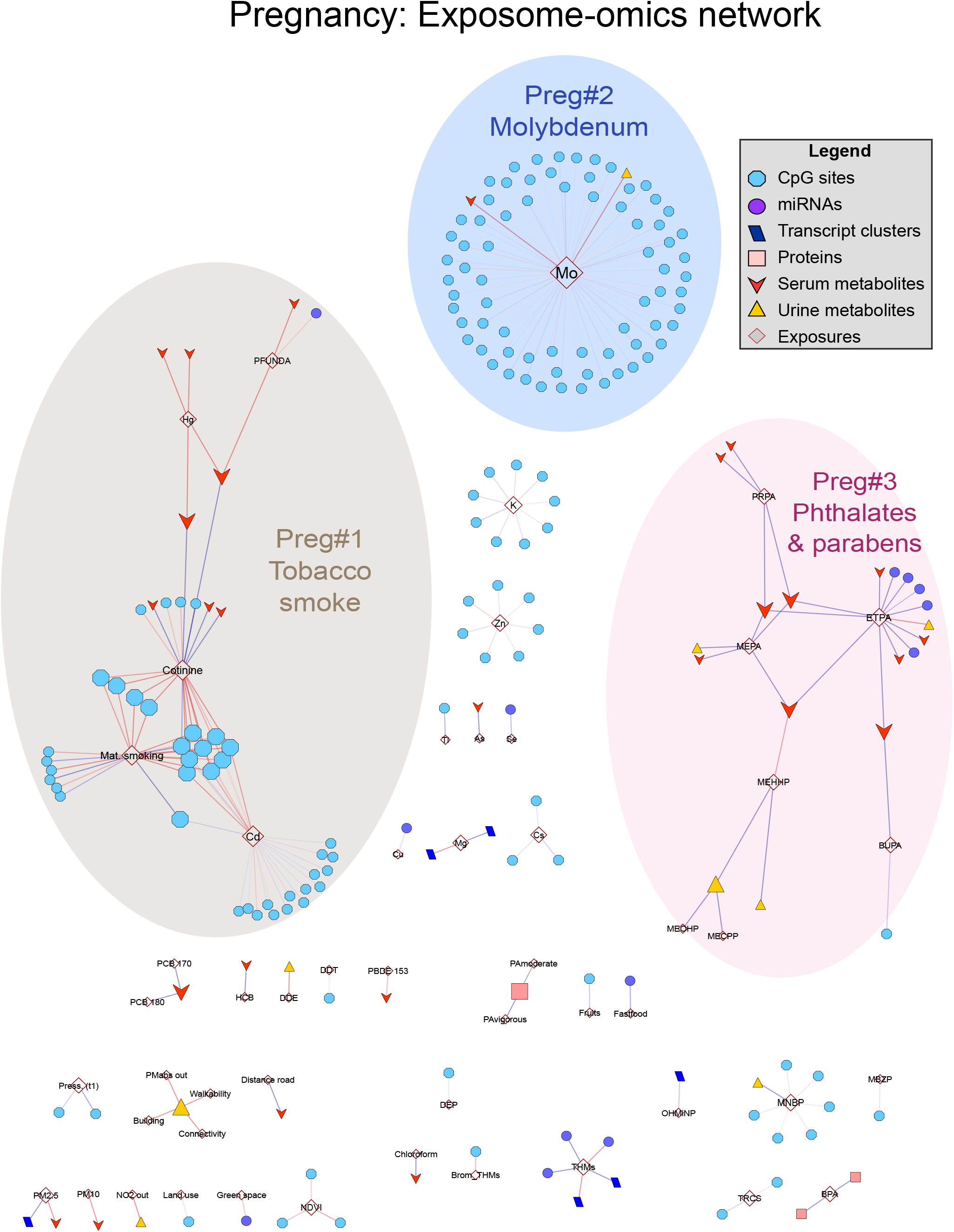
Network map of the multi-omics signatures of the pregnancy Exposome. Network visualization of the pregnancy exposome-omics-wide association analysis (ExWAS). An exposure and a molecular feature were connected if their association was statistically significant and only connected components with at least two molecular features were displayed. Three main connected components were annotated, which varied greatly by their size, their number of exposures and the type of omics composing them.

**Figure 4.**
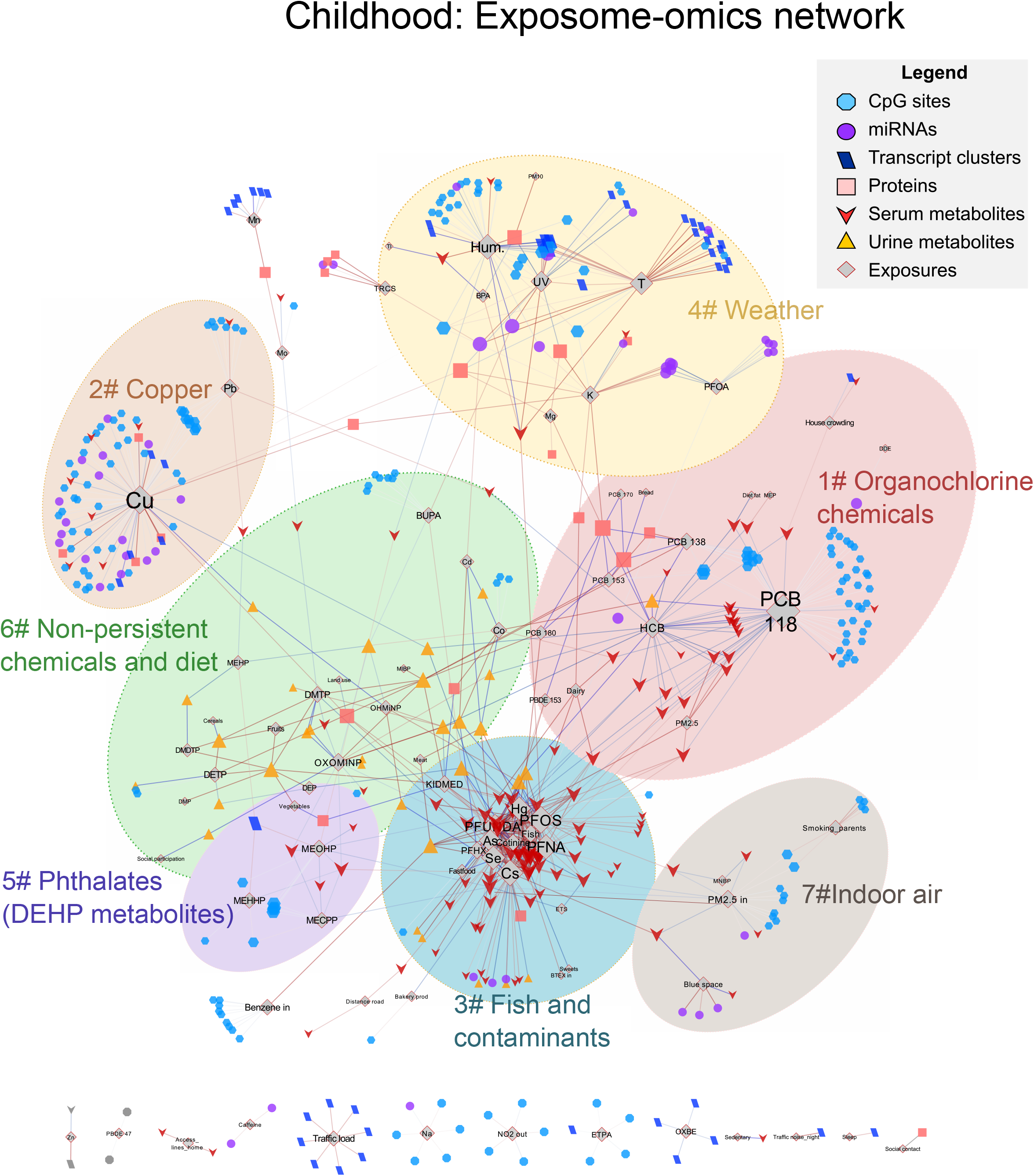
Network map of the multi-omics signatures of the childhood Exposome. Network visualization of the childhood exposome-omics-wide association analysis (ExWAS). An exposure and a molecular feature were connected if their association was statistically significant and only connected components with at least two molecular features were displayed. The childhood exposome network was diverse in terms of omics features represented and the level of interconnection, with the biggest connected component containing 90% of all nodes. Within this network, 11 interconnected subcomponents (i.e. clusters) were identified using an unsupervised structural cluster analysis, seven of them were annotated.

The childhood exposome network was more diverse in terms of omics features represented and the level of interconnection, with the biggest connected component containing 90% of all nodes **(Figure 4; Table S2)**. This high connectivity highlights the correlated nature of the serum and urine metabolome. Within this network, we identified 11 interconnected subcomponents (i.e. clusters, named as “childhood#…”) using an unsupervised structural cluster analysis **(Table 1)** (Newman and Girvan, 2003; Su et al., 2010).

In summary, over the two exposome periods, we found 14 major exposome-omics clusters spanning many diverse exposure families. The clusters reveal both potential biological pathways and potential routes or sources of exposure, as further described below.

#### Maternal smoking shows robust and long-lasting effects in the child methylome and we detect novel signatures for prenatal cadmium

Biological signatures for smoking have been relatively well documented, particularly for methylation signatures: maternal smoking during pregnancy has been associated with altered patterns of blood DNA methylation at birth (Bauer et al., 2016; Joubert et al., 2012, 2016), and some loci have shown persistent dysregulation until childhood (Joubert et al., 2016; Richmond et al., 2015), adolescence (Lee et al., 2015; Richmond et al., 2015) or even adulthood (Tehranifar et al., 2018). In our analysis, maternal smoking during pregnancy reported from questionnaires and urinary maternal cotinine levels (cluster preg#1) associated with 24 unique CpGs, representing 9 unique loci (2 Mb) annotated to 8 genes, that largely corroborate previous findings described in the EWAS Atlas/Catalog **(Supplemental Experimental Procedures; Figure 5A; Table S3)**. Additionally, we observed that child exposure to second-hand smoke (cluster childhood#7) overlapped with existing literature, but to a lesser extent than maternal smoking **(Figure 5C; Table S3)**. Period specific effects have been investigated elsewhere in more detail (Vives-Usano et al., 2020). In order to understand which pathways were perturbed due to exposure to tobacco smoke, we performed functional enrichment analysis of genes annotated to molecular features associated with pregnancy or childhood exposure **(Supplemental Experimental Procedures)**. The analysis identified the following functions: axon development, cognition, cholinergic synapse, insulin signalling, and several types of cancer **(Figures 5B and 5D; Table S4**, highlighted in yellow). These pathways are in line with the effects of maternal smoking on health detected in HELIX children: higher blood pressure (Warembourg et al., 2019) and BMI (Vrijheid et al., 2020), and increased behavioural problems (Maitre *et al*. under review).

**Figure 5.**
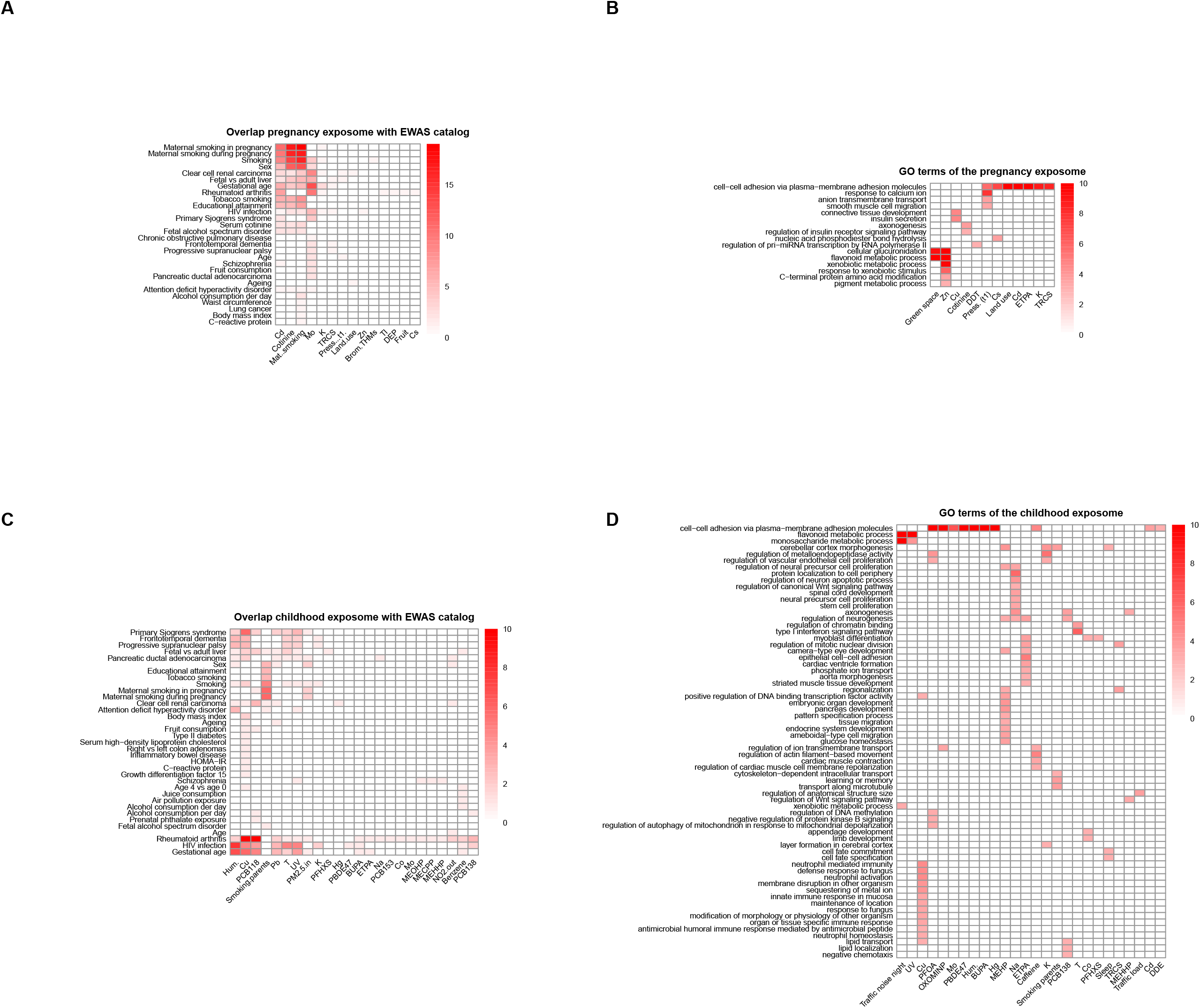
Biological interpretation of the exposome-omics associations through literature trait overlap and functional enrichment. **A)** Overlap of CpGs associated with the pregnancy exposome (columns) with CpGs associated with traits/exposures in the EWAS catalog (rows). **B)** Functional enrichment analyses of the pregnancy exposome (columns) for Gene Ontology (GO) terms (rows). **C)** Overlap of CpGs associated with the childhood exposome (columns) with CpGs associated with traits/exposures in the EWAS catalog (rows). **D)** Functional enrichment analyses of the childhood exposome (columns) for GO terms (rows). Exposure variables, traits/exposures of the EWAS catalog, and GO terms are ordered according to a hierarchical clustering. For the overlap with the EWAS catalog, colour indicates the number of overlapping CpGs. For the functional enrichment analyses, colour indicates the –log10 adjusted p-value of the enrichment. To facilitate visualization, we eliminated related GO terms and –log10 adjusted p-values >10 are coded as 10.

We further observed that exposure during pregnancy to the heavy metal cadmium (Cd) was associated with child blood methylation, and clustered with maternal smoking in cluster preg#1. This could be partially explained by the fact that Cd is a component of tobacco (Satarug, 2018) and in our dataset smoker mothers showed almost twice the level of Cd compared to non-smokers. However, we identified 14 additional CpGs that were unique to Cd; these did not overlap with known smoking effects, nor with CpGs associated with maternal Cd in the placenta **(Figure S1B; Table S5)**(Everson et al., 2018). When restricting our analysis of maternal Cd to non-smoker mothers (N=998), 51 CpGs (48 loci) were identified, which did not overlap with smoking effects, but did correspond to CpGs previously associated with conditions such as asthma, gestational diabetes, and gestational age **(Figure S1D; Table S5)**. Besides tobacco smoke, the main sources of Cd exposure are contaminated foods such as rice, potatoes and wheat, when frequently consumed in large quantities (Järup and Åkesson, 2009).

#### Indoor air quality during childhood is related to biomarkers of obesity and insulin resistance

We found several associations for indoor air quality during childhood (clusters childhood#7 for PM2.5 absorbance, and childhood#9 for benzene), in contrast to the few associations found for outdoor air pollution (5 CpGs with NO_2_ and 5 serum phosphatidylcholines with PM_2.5_ that do not overlap with indoor air pollution signatures). Individual indoor air pollution levels were estimated through prediction models trained from real measures with air monitors installed in the homes and self-reported data on home characteristics and smoking habits of the parents, among other factors (Tamayo-Uria et al., 2019).

Home indoor air pollution exposure to benzene was associated with 9 CpGs, one of them previously related to PM2.5 levels **(Figure S5C; Table S3)**. Indoor levels of PM2.5 absorbance, a marker of black/elemental carbon originating from combustion, were associated with methylation of 9 CpGs, including two in common with tobacco exposure **(Figure S5C; Table S3)**, and with decreased levels of serum branched amino acids (aka BCAA: Ile, Leu, and Val), acylcarnitine C4 (butyrylcarnitine) and two sphingolipids. There is emerging, although somewhat conflicting evidence of metabolic changes related to outdoor air pollution in humans and animals (Brower et al., 2016; Miller et al., 2015; van Veldhoven et al., 2019). Similar to our results, lower BCAA and acylcarnitines were detected in young obese participants exposed to near-roadway air pollution (Chen et al., 2019). In sensitivity analyses, where models were not adjusted for child BMI, the associations of indoor PM2.5abs with acylcarnitine C4 and three BCAA were even stronger (15-20% fold change in effect size). Associations between dysregulated metabolism of BCAAs and acylcarnitines with obesity and insulin resistance have been widely observed in animal and adult human studies (Newgard, 2017). We propose that altered BCAA and acylcarnitine metabolism may be an important biomarker to study further in relation to indoor air pollution and subsequent development of cardio-metabolic disease in later life. An association between indoor air pollution and increased child BMI was previously reported in the HELIX study, independently of correlated exposures such as second-hand smoke and lower social class status (Vrijheid et al., 2020).

#### The serum and urinary metabolome reveal principal dietary routes of exposure to chemical pollutants

The metabolome is a key molecular layer to detect intermediary physiological changes in response to environmental influences, but it is also composed of exogenous compounds which are internalized through different routes: principally diet, but also via airways or skin absorption. We observed the childhood exposome to be associated with the metabolome in a dense, interconnected manner **(Figures 4)**. Three clusters contained most of the serum and urine metabolites: childhood#1 contained persistent chemicals (see next section); cluster childhood#3 **(Figure 6A)** contained fish intake, toxic metals (mercury (Hg) and arsenic (As)), the persistent PFASs, and non-toxic essential elements (selenium (Se) and caesium (Cs)); and cluster childhood#6 **(Figure 6B)** contained fruit and vegetable diet-related variables and organophosphate (OP) pesticides.

**Figure 6.**
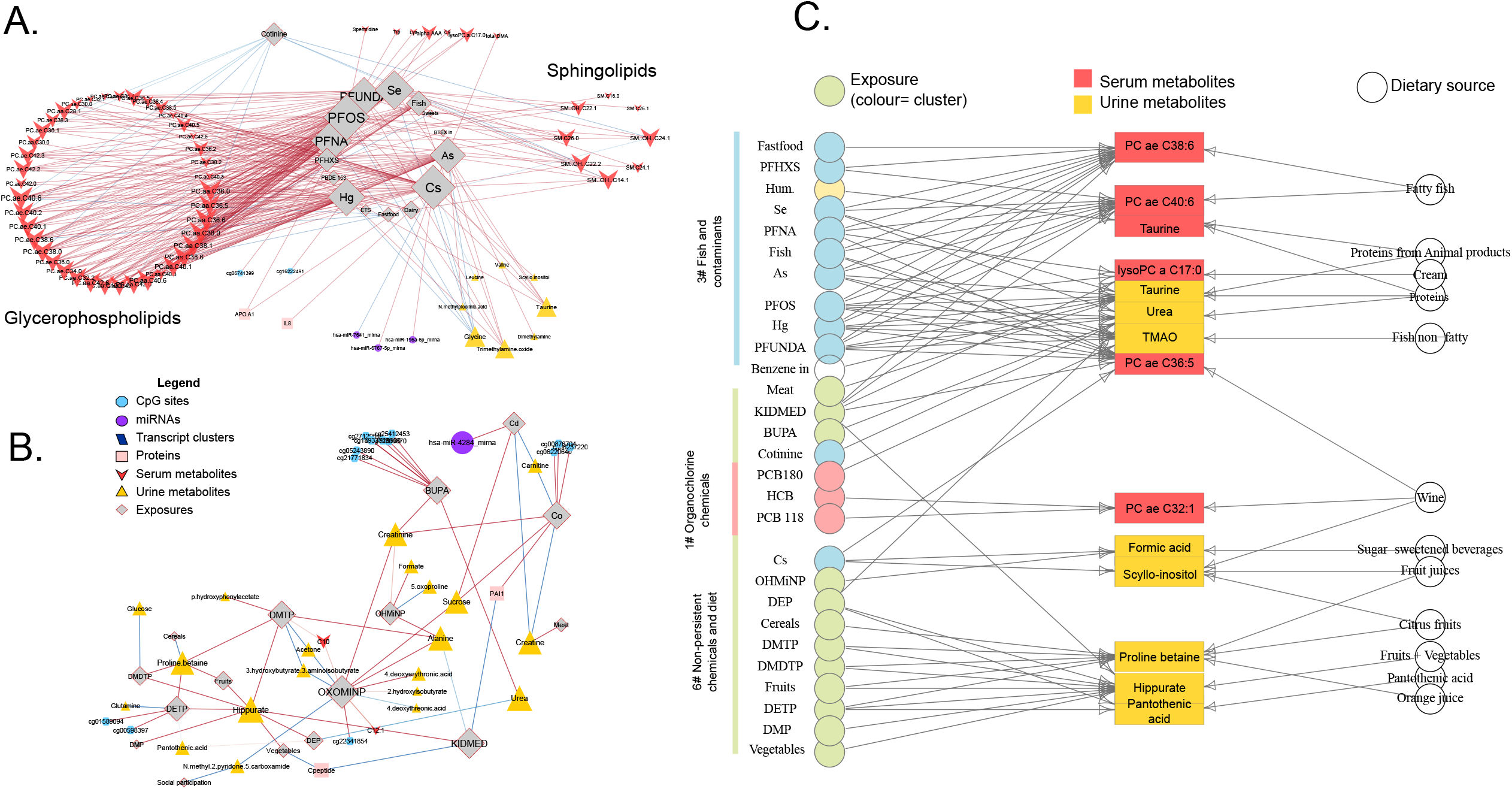
Metabolite signatures of the childhood exposome and dietary sources. **A)** Cluster childhood#3, which includes the associations between fish and contaminants (As, Hg, PFOS) and serum metabolites (mainly glycerophospholipids); **B)** Cluster childhood#6, which includes the associations between diet (vegetables, fruit, cereals) and OP pesticides with urinary metabolites; **C)** Previous literature linking urine and serum metabolites with food items, based on the database Exposome Explorer (http://exposome-explorer.iarc.fr/).

Cluster childhood#3 “fish and contaminants” was enriched for metabolic biomarkers of fish intake such as serum lipids containing polyunsaturated fatty acids (PCaaC40:6, PCaaC36:5, PCaaC36:6 and PCaaC38:6) and urinary trimethylamine N-oxide (TMAO), dimethylamine and homarine (*aka. N*-methylpicolinic acid). Fish is a source of essential elements (selenium (Se), caesium (Cs), but it also bio-accumulates toxic metals (mercury (Hg) and arsenic (As)) and the persistent PFASs) (Avella-Garcia and Julvez, 2014; Christensen et al., 2017), which were also found in the cluster. These fish metabolites were previously found to be associated with Hg and As in pregnant women (Maitre et al., 2018b).

Cluster childhood#6 “fruit and vegetables” contained mostly urinary metabolites (21 out of 44 measured), including hippurate, proline betaine and *N*-methylnicotinic acid (trigonelline) which are known biomarkers of fruit and vegetable intake (Heinzmann et al., 2015; Lau et al., 2018). Interestingly, these metabolites were associated with organophosphate (OP) pesticides (DEP, DETP, DMP, DMTP and DMDTP measured in urine), a class of insecticides still applied in agriculture for insect control on food crops. Intake of fruits and vegetables is a main determinant of OP urinary metabolites in children and pregnant women, as previously found in the HELIX cohort (Papadopoulou et al., 2019). This suggests that the observed association between child OP levels and known biomarkers of fruit and vegetable intake is likely to arise from a higher fresh fruit and vegetable intake.

Using ExposomeExplorer (Neveu et al., 2017), a catalogue of 908 dietary and pollutant biomarkers measured in population studies **(Supplemental Experimental Procedures)**, we were able to confirm the dietary origin of many exposure-metabolite associations observed in our dataset **(Figure 6C)**. These findings demonstrate the ability of metabolomics to accurately reflect dietary patterns.

#### Essential trace elements are key components of the exposome

Essential trace elements are required by living organisms to ensure normal fetal development and growth, and maintenance of biological functions, but can also be toxic when present in excess. We measured 9 essential elements (Co, Cu, Mn, Mo, Na, K, Mg, Zn, Se) and found a remarkable number of omics associations with maternal molybdenum (Mo), and child copper (Cu).

Molybdenum (Mo), the exposure in pregnancy with the most associations (74, including 72 CpGs from 63 loci; cluster preg#2) **(Figure 3)**, acts as a co-factor of 4 human enzymes (sulphite oxidase, xanthine oxidoreductase, aldehyde oxidase and mitochondrial amidoxime reductase) which are involved in various key reactions of aldehydes, purines, and sulfur metabolism (Schwarz, 2016). Interestingly, sulphite oxidase is involved in the catabolism of sulphur amino acids such as methionine, which serves as a donor of methyl groups for DNA methylation (Clare et al., 2019). In our data, we found that >70% of the significant CpGs were hypo-methylated in relation to maternal Mo, and that maternal Mo was associated with higher methionine levels in childhood. Thirteen of the CpGs associated with maternal Mo have previously been related to gestational age **(Figure S5A; Table S3)**. Human exposure to Mo may occur via diet, water consumption, inhalation from molybdenum fertilizer, nanoparticles, and/or occupational exposure from mining operations or other industrial uses (Mohamed et al., 2020; Novotny and Peterson, 2018; Vázquez-Salas et al., 2014). It is generally believed that Mo is safe for human health (Novotny and Peterson, 2018). However, there is growing evidence that excess of Mo is associated with some adverse health outcomes in the general population (Meeker et al., 2008, 2010) and with developmental effects *in utero* (Gauglitz et al., 2020; Vázquez-Salas et al., 2014; Yin et al., 2020; Zheng et al., 2020). Reproductive and genotoxic effects of Mo have been reported in animals with retarded fetal growth (Mohamed et al., 2020; Tallkvist and Oskarsson, 2015). This points to the possible importance of Mo exposure during pregnancy and DNA methylation patterns with possible long-lasting impact on the child molecular phenotype.

During childhood, essential trace elements were associated with multiple omics layers, with little overlap across molecular features **(Figure 5)**. Copper (Cu) (cluster childhood#2), was associated with 89 molecular features, distributed across different omics layers. One of the most robust (lowest p-value) Cu associations was with increased levels of the C-reactive protein (CRP), an inflammatory biomarker. Cu-associated CpGs have previously been described in relation to obesity, type 2 diabetes and rheumatoid arthritis, a chronic inflammatory disorder, among others **(Figure 5C; Table S3)**. Enriched gene-sets included: immune response, lipid storage and sequestering of metal ions **(Figure 5D; Table S4**, highlighted in green). Cu is an essential trace element required for numerous cellular processes, including mitochondrial respiration, antioxidant defense, neurotransmitter synthesis, connective tissue formation, pigmentation, peptide amidation and iron metabolism (De Bie et al., 2007). In line with the numerous Cu biological functions, we found in HELIX that child Cu was associated with poorer lung function (Agier et al., 2019), higher BMI (Cadiou et al., 2020; Vrijheid et al., 2020) and higher blood pressure (Warembourg et al., 2019), and increased prevalence of behavioural problems and ADHD symptoms (Maitre *et al*. under review).

Some of the other essential trace element associations validated previous knowledge of their regulatory role on protein synthesis/enzymatic activity. We found that zinc (Zn) was related to higher transcription of *CA1* (*Carbonic anhydrase 1*), whose expression is known to be influenced by Zn^2+^ availability (Lionetto et al., 2016) and which uses Zn^2+^ as a cofactor. Also, cobalt (Co) was associated with higher levels of PAI1 protein (Plasminogen Activator Inhibitor Type 1), consistent with previous studies which show that CoCl_2_ induces PAI-1 mRNA and regulates its activity (Long and Schafer, 2008; Thompson et al., 2011).

Overall, our data highlights the importance of essential trace elements as key components of the exposome. We corroborated known regulatory effects of Co and Zn, and in addition, we discovered novel biological interactions for maternal Mo and child Cu. Since only trace amounts of Mo and Cu from diet are required to achieve optimal health, further studies on the potential long-term consequences of excess of these elements, rather than deficiency, are warranted.

#### Molecular signatures of persistent organic pollutants implicate body mass and hepatic metabolic alterations in children

Developmental exposure to persistent organic pollutants (POPs), so called because they persist in the environment and bio-accumulate through the food chain in human and animal fatty tissues, has consistently been associated with adverse birth outcomes and neurotoxicological outcomes (Vrijheid et al., 2016). However, evidence for child health effects associated with childhood exposure is inconsistent.

We found that POPs in children, especially dioxin-like PCB118 (69 associations), HCB (28) and PCB138 (14), were associated with DNA methylation, serum metabolites and plasma proteins (IL1B and leptin) grouped in cluster childhood#1. CpGs in this cluster have previously been reported to be related to the inflammatory disease rheumatoid arthritis **(Figure 5C; Table S3)**. Several serum sphingolipids and glycerophospholipids, inversely associated with POPs, were previously found to be related to higher BMI in HELIX children (Lau et al., 2018). Similarly, POPs were related to lower levels of proteins that are normally enhanced in obesity. These included, which is mainly produced by fat cells of the adipose tissue and regulates appetite and energy expenditure (Fain, 2010), and IL1β, an inflammatory adipokine primarily released by the non-fat cells of adipose tissue (Yao et al., 2014). These associations were even stronger when models were not adjusted for child BMI (mean relative change +192%), indicating a strong influence of BMI **(Figure S2)**. Indeed, in HELIX, we also observed an inverse relationship between POPs and adiposity and BMI (Vrijheid et al., 2020). The serum levels of POPs depend on exposure dose and timing, but also, as they accumulate in adipose tissue, on the amount and type of fat tissue and on recent changes in body weight/composition (Jackson et al., 2017; Wolff et al., 2007; Wood et al., 2016). Thus, future studies, especially in growing children, should consider including pharmacokinetic-pharmacodynamic (PKPD) modeling to elucidate the role of fat tissue distribution in observed biological effects of POPs.

Besides associations likely linked to fat distribution in children, we also observed a positive association of PCB180 and TMAO, a product of gut microbiota and liver hepatic flavin containing monooxygenase (FMO3) enzyme activity. This association was previously reported in animals and humans and appeared independent of potential common dietary sources of PCBs and TMAO and of BMI (Petriello et al., 2018). Currently, TMAO is proposed as a causative agent of cardio-vascular disease (Zhu et al., 2017) but further investigations on the mechanistic link between PCBs, FMO3 activity/expression and cardio-vascular outcomes are needed. In addition, we observed a downregulation of fatty acids associated with HCB and PCB118 possibly indicating a perturbation of global hepatic lipid metabolism as previously suggested in general population and adolescents with severe obesity exposed to organochlorine compounds (Carrizo et al., 2017; Salihovic et al., 2016; Valvi et al., 2020).

#### Endocrine disruptors, phthalates and parabens are associated with altered sphingomyelin levels

Phthalates are synthetic compounds widely found in personal care products, drugs, food packaging, building products and polyvinylchloride (PVC) derivates. They are rapidly metabolized in the body in <24h and suspected of being endocrine disruptors (Braun, 2017). We found associations mainly with metabolites of the high molecular weight phthalates DEHP and DiNP measured during childhood, but not with the low molecular weight congeners. DEHP and DiNP exposure occurs mostly through diet, dust ingestion, and to a less extent through inhalation (Giovanoulis et al., 2018).

The oxidized forms of DiNP (OHMiNP and OXOMiNP) mapped in cluster childhood#6 with many serum and urine metabolites. These metabolic signatures (particularly for creatinine in serum and urine) may be indicative of potential renal excretion patterns, in line with the fact that the oxidized forms are more polar, hydrophilic phthalate metabolites, which would lead to their rapid renal excretion. On the other hand, the metabolized forms of DEHP (MEOHP, MEHHP, MECPP, MEHP) mapped in cluster childhood#5 and were associated with 13 CpGs, with no clear overlap with reported traits/exposures **(Figure 5C; Table S3)**. MEOHP and MECPP were also negatively associated with a number of serum sphingomyelins (SM.C16.0, SM.C18.0, SM.C18.1, SM.C20.2, SM(OH)C14.1 and SM(OH)C16.1), which are important structural lipid components of cell membranes, are involved in signalling pathways and are implicated in many disorders.

Pregnancy exposure to DEHP metabolites and parabens (PRPA, ETPA, MEPA), another class of endocrine disruptors present in personal care products, showed negative associations with sphingomyelins (SM(OH)C16.1) and valine in children. Intermediates of sphingosine biosynthesis and valine have been reported to be upregulated in pregnant women exposed to phthalates (Zhou et al., 2018) and parabens (Zhao et al., 2020). Another study suggested persistent sex-specific effects of fetal exposure to phthalates and bisphenols on childhood lipid concentrations and glucose metabolism at a mean age of 9.7 years (Sol et al., 2020). There is increasing epidemiological evidence suggesting that prenatal exposure to phthalate contribute to childhood adverse health outcomes in a sex-specific manner (Haug et al., 2018). However, typically these exposures are hard to monitor in human population due to their rapid elimination in urine. Repeated sampling strategies and longitudinal designs, exploring the critical windows of phthalate and paraben exposures in pregnancy and early childhood might help to disentangle the persistent metabolic effects suggested in our study.

#### Weather conditions are associated with signatures in all omics layers

Weather conditions or meteorological factors (temperature, humidity, cloud coverage and atmospheric pressure), in particular when extreme, are strong determinants of health and mortality (EEA, 2017). However, there are no studies systematically assessing their influence on molecular phenotypes. We estimated weather conditions through geographical information coupled with data from meteorological stations, and in childhood averaged over the month before the omics measurement. In childhood, these were associated with all molecular layers (cluster childhood#4), except for the urinary metabolome. Serum metabolites associated with meteorological variables in cluster childhood#4, including taurine, asymmetric dimethylarginine (ADMA), acylcarnitine C5, and serotonin, have been previously reported as biomarkers of sleep deprivation, circadian rythm and in the aetiology of depression (Davies et al., 2014; Nasca et al., 2018; Selley, 2004). This cluster also included three proteins (adiponectin, MCP1 and HGF), several miRNAs, small nucleolar RNAs, and CpGs. Interestingly, adiponectin, an essential regulator of thermogenesis (Jankovic et al., 2013; Wei et al., 2017), increased with humidity (which is higher in winter in Europe) and decreased with ultraviolet radiation (higher in summer). This is in line with previous studies showing that exposure to cold temperatures for 2h increases adiponectin plasma levels (Imbeault et al., 2009).

The CpGs associated with weather conditions in this cluster overlapped with CpGs reported for gestational age and infections, among others **(Figure 5C; Table S3)**; and genes related to temperature were enriched for cellular response to type I interferon **(Figure 5D; Table S4**, highlighted in blue). Infectious diseases follow seasonal patterns and are more prevalent under particular meteorological conditions (Abhimanyu and Coussens, 2017; Lowen and Steel, 2014). Our findings provide evidence that, besides the direct effect of meteorological variables on human physiology (e.g. regulation of thermogenesis), they can indirectly affect child molecular profiles and potentially disease status by determining other exposures (e.g. virus survival), or they can represent proxies of other variables (e.g. hours of daylight). As far as we know, we are the first in reporting genome-wide and multi-omics signatures related to meteorological variables, and thus replication in larger longitudinal omics datasets covering seasonal patterns is needed to elucidate causal mechanisms.

#### Exposure periods, biological matrices and molecular layers capture different molecular properties of the exposome

We evaluated the overlap of the reported associations in our data across different exposome periods (pregnancy and childhood), biological matrices (serum and urine) and different molecular layers (DNA methylation, gene and miRNA expression), in order to better inform the design of future exposome studies.

Although a substantial number of exposures (83) were assessed both in pregnancy and childhood **(Supplemental Experimental Procedures)**, only 14 exposure-omics pairs were statistically significant in the two periods: 6 CpGs related to tobacco smoking, and several long chain fatty acids related to cotinine, hexachlorobenzene (HCB), perfluoroundecanoate (PFUnDA) and Hg **(Table S6)**. The inter-period correlation for these exposures was low (mean: 0.24). We can thus conclude that biological signatures of the pregnancy and childhood exposomes had minimal overlap.

Regarding biological matrices, we compared the overlap of exposure associations for 12 metabolites (amino acids, glucose, carnitine and creatinine) that were measured in both urine and serum. In urine, 9 out of the common 12 metabolites were associated with the childhood exposome with a total of 33 significant associations; and in serum, 7 out of the 12 metabolites were associated with exposures giving a total of 14 associations **(Tables S7)**. At nominal significance, 27.3% of the urine associations replicated in serum and 7% of the serum associations replicated in urine. Assuming similar quality of metabolic data between platforms, these results suggest that a substantial proportion of associations with the exposome are biofluid specific.

Next, we investigated whether different molecular layers (DNA methylation, gene and miRNA expression) in the same biological matrix (blood cells) pointed to the same biological pathways. For each CpG we searched for cis expression quantitative trait methylations (eQTMs), meaning correlations between gene expression and DNA methylation in 1 Mb window **(Supplemental Experimental Procedures)**. Out of the 187 CpGs associated with the childhood exposome, 9 showed eQTMs for a total of 11 genes **(Table S8)**, however the expression of none of these was nominally associated with the same exposures as the CpG site. We also searched for experimentally validated targets of the 49 miRNAs associated with the childhood exposome using the miRwalk v3 tool (Sticht et al., 2018), and identified 1,267 targeted genes, representing 44 unique mature miRNAs **(Table S8)**. Only 17 of these genes were associated with the same exposure as the original miRNA and in the expected direction (higher miRNA levels - lower gene expression) in a total of 18 associations **(Table S8)**. They encompassed 7 unique exposures (Cd, Cu, K, PFOA, blue spaces and meteorological factors) and 9 unique miRNAs. Overall, the low correspondence between the methylome and miRNAome with the transcriptome highlights the high complexity of transcriptional regulation and suggests that each molecular layer might capture a piece of the effects of the exposome.

#### Robustness of results with respect to ancestry and country

HELIX consists of 1,171 European ancestry children and the rest from other ancestries, with Pakistani ancestry the second most common (102 children), according to self-report. We repeated the ExWAS in only European ancestry children **(Supplemental Experimental Procedures)**, and no major differences in effect sizes was observed **(Figures S3)**. The mean relative change in effect size between the two models was -23.9%, with a similar distribution for the pregnancy and postnatal exposomes.

We also investigated heterogeneity across cohorts by running the statistically significant exposure-omics associations by cohort **(Supplemental Experimental Procedures)**. Heterogeneity (I^2^) depended on period and molecular layer **(Figure S4)**. Around half of all associations presented I^2^ <0.5. While some associations seemed to be very consistent among cohorts (*e*.*g*. maternal cadmium and methylation) others were more cohort dependent (*e*.*g* child meteorological conditions and serotonin).

## Discussion

This is the first exposome study to systematically associate a wide range of environmental exposures during vulnerable early life periods with multi-omics signatures in childhood. We observed 1,170 unique associations between exposures and molecular features, 249 relating to pregnancy and 921 to childhood exposures. By partitioning these associations into clusters, this study reveals potential biological responses and routes of exposure. Our findings confirm persistent methylation changes associated with maternal tobacco smoking in pregnancy and principal routes of exposure to chemical pollutants through diet based on food-related biomarkers. Furthermore, we identify novel omics associations with indoor air quality, essential trace elements, endocrine disruptors and weather conditions. Our comprehensive resource of all associations (https://helixomics.isglobal.org/) is the first of its kind and will serve to guide future investigation on the biological imprints of the early life exposome.

Among the novel molecular signatures identified, some highlight plausible biological mechanisms to disease. For instance, copper, has been related to several health outcomes in HELIX children (lung function, blood pressure, BMI and ADHD) and here we show potential perturbed pathways that may mediate these associations: immune response, lipid storage and sequestering of metal ions. Furthermore, data generated in this study provide a resource for the development of epigenetic biomarkers of past exposures (Reese et al., 2017). For instance, the essential element molybdenum (Mo) during pregnancy is associated with methylation changes in a remarkable number of CpGs, which can be used to predict prenatal exposure. Also, our study demonstrates the ability of metabolomics to accurately reflect variance in dietary exposure. Diet is a complex, multidimensional exposure, and studying its contribution to health requires a multipronged approach, for which metabolomics is well positioned (Garcia-Perez et al., 2017; Lau et al., 2018; Posma et al., 2020, Stratakis, Siskos in preparation). Importantly, we illustrate in this study that many anthropogenic chemicals are also delivered in the body through the diet, adding to the complexity of metabolomic profiles in human biospecimens and creating an extensive network of nutrient–pollutant interactions that remains vastly unknown and poorly defined by conventional assessment methods (Cano-Sancho and Casas, 2020).

Our results indicate that the choice of molecular layer and biological matrix is key in the design of exposome studies. We found that the prenatal environment is mainly captured by the methylome with sustained changes until childhood, which suggests that the methylome acts as the main source of biological ‘memory’ for the *in utero* exposome. In contrast, recent exposures during childhood were associated with features across all omics layers. Evidence to date suggests that the metabolome in particular is strongly influenced by the immediate environment, and may thus be more sensitive for detecting associations in cross-sectional settings (Everson and Marsit, 2018). However, although to a lesser extent, the metabolome also captured persistent biological responses to the pregnancy exposome (e.g. to phthalates and parabens). On the other hand, our findings were clearly dependent on the biological matrix that was assessed. For instance, only a few of the exposure-metabolite associations were found both in serum and urine. Sampling one matrix and not another has clear implications in the biological mechanisms that can be detected, and thus on the exposure-omics associations that can be identified.

The main strengths of our study are: 1) the comprehensive assessment of environmental exposures in two critical developmental time periods (pregnancy and childhood), and including highly sensitive biomarkers for many chemical exposures and wide-ranging geospatial modelling of the outdoor and built environment; 2) the extensive multi-omics assessment of molecular phenotypes; and 3) the wide geographic coverage and relatively large sample size for which we were able to measure many exposures and omics features.

Our study also has some limitations. First, omics platforms have a coverage bias and biological interpretability issues. For instance, even genome-wide platforms such as the 450K methylation array cover a small fraction (1-2%) of all CpGs in the genome and their functional interpretation has been simplified by assuming they regulate the closest gene. The LC-MS/MS (Biocrates) method has low coverage and does not give specific fatty acid side-chain composition for lipids, but it is widely used in large cohort studies and provides reproducible measurements with unambiguous annotation, easily comparable to other studies (Floegel et al., 2013; Illig et al., 2010; Siskos et al., 2017; Varma et al., 2018; Wong et al., 2008). We note that there are additional molecular layers and omics technologies of interest for future exposome studies, which were not included in our study, such as the gut metagenome, adductomics, sensitive high-resolution mass spectrometers or single cell methods (Jiang et al., 2018; Petrick et al., 2020; Walker et al., 2019). Second, different exposures are measured with different types and levels of measurement error. For example, urine levels of non-persistent chemicals have a high intra-individual variability and are expected to suffer particularly from classical-type measurement error resulting in an attenuation bias (Casas et al., 2018). Exposures measured by models and questionnaires are expected to suffer from other types of measurement errors with less predictable effects (Nieuwenhuijsen et al., 2015). Moreover, the correlated nature of the exposome makes identification of driving exposures difficult. Mixture or multi-pollutant approaches aim to tackle this, however these are not yet suitable for high-dimensional datasets such as ours (Jain et al., 2018; Park et al., 2017). Third, our comparison with previous literature and functional enrichment analyses are limited by existing bias in public databases. Fourth, although the majority of epidemiological studies utilize biological samples that are most readily accessible for the measurement of omics profiles, these may not be the ideal target tissue for the relevant health outcomes. Biofluids such as plasma/serum and urine, solve this in part as they contain molecules from peripheral organs, but they clearly are limited in their ability to capture some other biological events. Finally, although our models were adjusted for confounders, causal links would need to be proven through interventions, Mendelian randomization analyses or cross-contextual studies in order to move to therapeutic and preventative strategies.

To conclude, this first comprehensive study of the multi-omics signatures of the early life exposome demonstrates that molecular phenotypes can reveal biological responses to, or sources of, environmental exposures at an early time point in life. This can help to improve our understanding of biological mechanisms and, ultimately, to detect and prevent environmental damage to health earlier in life, before clinical manifestation. Besides the main findings described here, the entire result catalogue is publicly available (https://helixomics.isglobal.org/), enabling exploration of the complete list of exposome-omics relationships. With the rich exposome and molecular information available, we provide a valuable resource to the scientific community for the development and validation of exposure and response biomarkers and to improve our understanding of disease aetiology.

## Data Availability

The summarized results (exposure, omics biomarker, effect, standard error, p-value) generated during this study are available at https://helixomics.isglobal.org/. The raw data supporting the current study are available from the corresponding author on request subject to ethical and legislative review.

## Code availability

The code to test the relationship between the pregnancy and childhood exposomes and molecular features is available through the R package *omicRexposome* (Hernandez-Ferrer et al., 2019).

## Acknowledgements

We would like to thank all the families for their generous contribution. The study has received funding from the European Community’s Seventh Framework Programme (FP7/2007-206) under grant agreement no 308333 (HELIX project) and the H2020-EU.3.1.2. - Preventing Disease Programme under grant agreement no 874583 (ATHLETE project). LM is funded by a Juan de la Cierva-Incorporación fellowship (IJC2018-035394-I) awarded by the Spanish Ministerio de Economía, Industria y Competitividad. BiB received core infrastructure funding from the Wellcome Trust (WT101597MA) and a joint grant from the UK Medical Research Council (MRC) and Economic and Social Science Research Council (ESRC) (MR/N024397/1). INMA data collections were supported by grants from the Instituto de Salud Carlos III, CIBERESP, and the Generalitat de Catalunya-CIRIT. KANC was funded by the grant of the Lithuanian Agency for Science Innovation and Technology (6-04-2014_31V-66). The Norwegian Mother, Father and Child Cohort Study is supported by the Norwegian Ministry of Health and Care Services and the Ministry of Education and Research. The Rhea project was financially supported by European projects (EU FP6-2003-Food-3-NewGeneris, EU FP6. STREP Hiwate, EU FP7 ENV.2007.1.2.2.2. Project No 211250 Escape, EU FP7-2008-ENV-1.2.1.4 Envirogenomarkers, EU FP7-HEALTH-2009-single stage CHICOS, EU FP7 ENV.2008.1.2.1.6. Proposal No 226285 ENRIECO, EU-FP7-HEALTH-2012 Proposal No 308333 HELIX), and the Greek Ministry of Health (Program of Prevention of obesity and neurodevelopmental disorders in preschool children, in Heraklion district, Crete, Greece: 2011-2014; “Rhea Plus”: Primary Prevention Program of Environmental Risk Factors for Reproductive Health, and Child Health: 2012-15). ISGlobal acknowledges support from the Spanish Ministry of Science and Innovation through the “Centro de Excelencia Severo Ochoa 2019-2023” Program (CEX2018-000806-S), and support from the Generalitat de Catalunya through the CERCA Program. MV-U and CR-A were supported by a FI fellowship from the Catalan Government (FI-DGR 2015 and #016FI_B 00272). MC received funding from Instituto Carlos III (Ministry of Economy and Competitiveness) (CD12/00563 and MS16/00128).

## Author contributions

LM, MB, JW, RS, JS, CT, MC, JRG, HK and MV designed the omics study in HELIX. The following authors participated in omics data acquisition and quality control: AC, IQ, MB, CR-A (DNA methylation), XE, MB, MV-U (transcriptomics), ES, EB (proteomics), CEL, APS, LM, HK, MC (metabolomics). JW, DM, RS, BH, JS, MCasas, KBG, EP, RG, LCh, CT and AKS are the PIs of the cohorts or participated in sample and exposure data acquisition. MCasas, CT, AKS, MN and IT measured the pregnancy and postnatal exposomes. CH-F, LM, MB, DT, SC, JU and JRG, performed statistical analyses and JRG functional enrichment analyses. The HELIX project was coordinated by MV. LM, MB and MV wrote the original draft of the paper and CH-F, APS, OR, JW, DM, SC, KBG, EP, CT, AKS, MN, JU, MC and HK contributed to reviewing and editing the manuscript. All authors read and approved the manuscript.

---

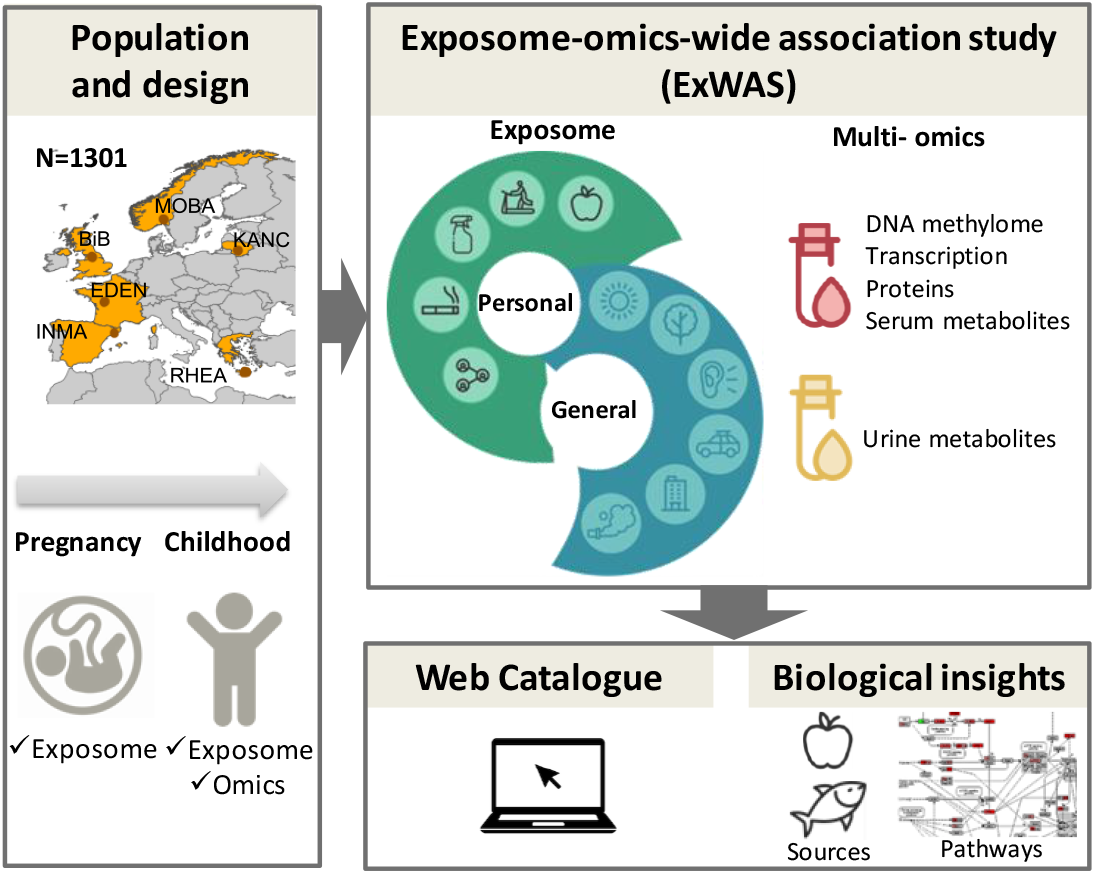

