## Supplementary methods for "Multi-omics signatures of the human early life exposome"

**Supplemental Experimental Procedures**

A summary of the data and the steps followed for the statistical analysis and biological interpretation can be seen in **Figure 1**.

***Population***

The HELIX study is a collaborative project across 6 established and ongoing longitudinal population-based birth cohort studies in Europe: the Born in Bradford (BiB) study in the UK (Wright et al., 2013), the Étude des Déterminants pré et postnatals du développement et de la santé de l’Enfant (EDEN) study in France (Heude et al., 2016), the INfancia y Medio Ambiente (INMA) cohort in Spain (Guxens et al., 2011), the Kaunus cohort (KANC) in Lithuania (Grazuleviciene et al., 2009), the Norwegian Mother and Child Cohort Study (MoBa) (Magnus et al., 2016), and the RHEA Mother Child Cohort study in Crete, Greece (Chatzi et al., 2017). The subcohort is a nested study within HELIX encompassing 1301 children from these cohorts (**Tables S1**). A follow-up examination of the children between ages 6 and 11 years was carried out in order to assess child health outcomes, to fully characterize the pregnancy and childhood exposome, and to measure several molecular phenotypes (Maitre et al., 2018). Local ethical committees approved the studies that were conducted according to the guidelines laid down in the Declaration of Helsinki. The ethical committees for each cohort were the following: BIB: Bradford Teaching Hospitals NHS Foundation Trust, EDEN: Agence nationale de sécurité du médicament et des produits de santé, INMA: Comité Ético de Inverticación Clínica Parc de Salut MAR, KANC: LIETUVOS BIOETIKOS KOMITETAS, MoBa: Regional komité for medisinsk og helsefaglig forskningsetikk, Rhea: Ethical committee of the general university hospital of Heraklion, Crete. Informed consent was obtained from a parent and/or legal guardian of all participants in the study.

| **Table S1. Characteristics of the 1301 HELIX children** | |  |
| --- | --- | --- |
| Variable | N or Mean | % or SD^a^ |
| Cohort |  |  |
| BiB | 205 | 15.8 |
| EDEN | 198 | 15.2 |
| INMA | 223 | 17.1 |
| KANC | 204 | 15.7 |
| MoBa | 272 | 20.9 |
| RHEA | 199 | 15.3 |
| Age (years) | 7.98 | 1.6 |
| Sex |  |  |
| Male | 711 | 54.7 |
| Female | 590 | 45.3 |
| Ethnic group |  |  |
| White European | 1171 | 90.0 |
| Pakistani or Asian | 102 | 7.8 |
| Other | 28 | 2.2 |
| Obesity status^b^ |  |  |
| Underweight/normal | 927 | 71.4 |
| Overweight | 245 | 18.8 |
| Obese | 129 | 9.9 |
| Maternal education |  |  |
| Primary school | 183 | 14.1 |
| Secondary school | 444 | 34.1 |
| University degree or higher | 674 | 51.8 |
| Number of children with exposome data |  |  |
| Pregnancy exposome | 1301^c^ | 100.0 |
| Childhood exposome | 1301^c^ | 100.0 |
| Number of children with omics data |  |  |
| 6 omics | 871 | 66.9 |
| 4 or 5 omics | 312 | 24.0 |
| 2 or 3 omics | 29 | 2.2 |
| only 1 omics | 65 | 5.0 |
| no omics data | 24 | 1.8 |
| ^a^SD: standard deviation |  |  |
| ^b^According to WHO reference categories |  |  |
| ^c^Missings were imputed for the totality of the population | |  |

***Biological samples***

During the clinical examination, two spot urine samples (one before bedtime and one first morning void) were brought by the participants to the research centre in cool packs and stored at 4°C until processing. Urine samples of the night before the visit and the first morning void on the day of the visit were combined to provide Two urine samples, representing last night-time and first morning voids, were collected on the evening and morning before the clinical examination and were subsequently pooled to generate a more representative sample of the last 24 h for metabolomic analysis (n = 1107). Either the night-time void (n = 37) or morning void (n = 48) sample was analysed in cases where a pooled sample was missing.(Maitre et al., 2018).

Eighteen mL of blood were collected at the end of the clinical examination of the child to ensure an approximate 3 hours (median = 3.5 hours, SD = 1.1 hour) fasting time since the last meal. Blood samples were collected using a ‘butterfly’ vacuum clip and local anaesthetic and processed into a variety of sample matrices, including serum, plasma, whole blood for RNA extraction (Tempus tubes - Life Technologies, USA), red cells, and buffy coat for DNA extraction. These samples were frozen at -80°C under optimized and standardized procedures until analysis.

***Exposome measures in pregnancy and childhood***

Two main windows of exposure were considered, one cross-sectional including the exposome data of children at the same time as of omics sampling (childhood), and a prenatal window including the pregnancy period or measures of long term maternal exposures (e.g. persistent pollutants). A total of 91 prenatal and 116 childhood exposures were investigated in the study, including the outdoor exposome (air pollution, built environment, noise, green and blue space, and meteorological data), the individual exposome (cotinine, metals, POPs, PFAS, phthalates, phenols, and organophosphates) as well as lifestyle factors (exposure to tobacco smoking, diet and physical activity) (**Table S2**). They were measured in diverse ways as described below.

The outdoor exposome was assessed through GIS information and existing land use regression models adjusted for data from regulatory monitors and remote sensing data (Maitre et al., 2018). In this particular study we analysed whole pregnancy and childhood levels of air pollution at the home address (the year average before follow-up), while first pregnancy trimester and childhood monthly levels of meteorological variables. Built environment was calculated in a 300 m buffer.

Biomarkers of chemical exposures were measured in samples collected from the children at age 6-11 years and in samples previously collected from mothers during pregnancy. Biomarkers include: organochlorine compounds (OCs) and brominated compounds (PBDEs) in serum, perfluoroalkyl substances (PFASs) in plasma, metals and essential minerals in whole blood, and non-persistent chemicals (phthalates, phenols, organophosphate pesticides (OPs), and cotinine) in urine samples. Concentrations of OCs and PBDEs were adjusted for to total lipid percentage and expressed in ng/g of lipids. Urinary concentrations were adjusted for creatinine and expressed in μg/g of creatinine. Further details can be found elsewhere (Haug et al., 2018).

Lifestyle factors were assessed through standardized questionnaires: including the KIDMED questionnaires to assess Mediterranean diet (Serra-Majem et al., 2004), physical activity of the child, sleeping patterns of child, socioeconomic status (family affluence scale (Boyce et al., 2006), and subjective wealth), social capital of the family (Kritsotakis et al., 2008), exposure to environmental tobacco smoke, water consumption habits, cooking and heating methods at the home, cleaning products, bedroom location, noise perception, child's use of mobile phones and other devices, use of green spaces, commuting behaviour, holidays and sun exposure, and puberty development of the child. Concentrations of drinking water disinfection by products (DBPs) during pregnancy were estimated from water company concentration and habits obtained from questionnaire data. Data was not sufficiently complete to estimate child exposure to DBPs. Indoor air concentrations of NO_2_, PM_2.5_, PM_abs_, benzene, and TEX (toluene, ethylbenzene, xylene) were estimated by combining measurements in the homes of a subgroup of children during the two seasons with questionnaire data.

Exposures were either continuous variables or categorical variables with two or more levels. Continuous exposure variables were transformed to achieve linearity or categorized, when needed. Missing data were imputed using a chained equations method (White et al., 2011) implemented in the mice R package (Buuren and Groothuis-Oudshoorn, 2011). Twenty imputed datasets were created, although we only used the first imputation in this study. The correlation among exposures, within each exposure window or overall has been described in detail elsewhere (Tamayo-Uria et al., 2019).

**Table S2. Exposure list measured in the pregnancy and the childhood exposomes**

| **Pregnancy exposome** | | | **Childhood exposome** | | |  |
| --- | --- | --- | --- | --- | --- | --- |
| Group | Subgroup | Label | Group | Subgroup | Label | Correlation |
| Air Pollution | NO2 | NO2 (preg) | Air Pollution | NO2 | NO2 (year) | 0.641 |
| Air Pollution | PM10 | PM10 (preg) | Air Pollution | PM10 | PM10 (year) | 0.827 |
| Air Pollution | PM2.5 | PM2.5 (preg) | Air Pollution | PM2.5 | PM2.5 (year) | 0.644 |
| Air Pollution | PMAbsorbance | PMabsorbance (preg) | Air Pollution | PMAbsorbance | PMabsorbance (year) | 0.656 |
| Built Environment | Access | Accessibility (bus lines 300m) | Built Environment | Access | Accessibility (bus lines 300m) | 0.463 |
| Built Environment | Access | Accessibility (bus stops 300m) | Built Environment | Access | Accessibility (bus stops 300m) | 0.733 |
| Built Environment | Built density | Built density (300m) | Built Environment | Built density | Built density (300m) | 0.721 |
| Built Environment | Facility | Facility density (300m) | Built Environment | Facility | Facility density (300m) | 0.542 |
| Built Environment | Land use | Land use (300m) | Built Environment | Land use | Land use (300m) | 0.460 |
| Built Environment | Population | Population density | Built Environment | Population | Population density | 0.691 |
| Built Environment | Connectivity | Connectivity (300m) | Built Environment | Connectivity | Connectivity density (300 m) | 0.668 |
| Built Environment | Walkability | Walkability | Built Environment | Walkability | Walkability index | 0.419 |
| Meteorological | Humidity | Humidity (t1) | Meteorological | Humidity | Humidity (month) | 0.073 |
| Meteorological | Temperature | Temperature (t1) | Meteorological | Temperature | Temperature (month) | 0.343 |
|  |  |  | Meteorological | UV | UV - Vit.D (month) |  |
| Natural Spaces | Blue | Blue spaces (300 m) | Natural Spaces | Blue | Blue spaces (300 m) | 0.449 |
| Natural Spaces | Green | Green spaces (300 m) | Natural Spaces | Green | Green spaces (300 m) | 0.525 |
| Natural Spaces | NDVI | NDVI (100 m) | Natural Spaces | NDVI | NDVI (100 m) | 0.711 |
| Noise | Noise | Traffic noise (24h) | Noise | Noise | Traffic noise (24h) | 0.401 |
|  |  |  | Noise | Noise | Traffic noise (night) |  |
| Traffic | Traffic | Inverse distance to nearest road | Traffic | Traffic | Inverse distance to nearest road | 0.313 |
| Traffic | Traffic | Road traffic load (100 m) | Traffic | Traffic | Road traffic load (100 m) | 0.503 |
| Traffic | Traffic | Traffic density on nearest road | Traffic | Traffic | Traffic density on nearest road (home) | 0.438 |
|  |  |  | Traffic | Traffic | Traffic load of major roads (100 m) |  |
| Essential minerals | Co | Cobalt | Essential minerals | Co | Cobalt | 0.088 |
| Essential minerals | Cu | Copper | Essential minerals | Cu | Copper | 0.153 |
| Essential minerals | K | K | Essential minerals | K | K | -0.197 |
| Essential minerals | Mg | Mg | Essential minerals | Mg | Mg | 0.030 |
| Essential minerals | Mn | Manganese | Essential minerals | Mn | Manganese | 0.193 |
| Essential minerals | Mo | Molybdenum | Essential minerals | Mo | Molybdenum | 0.01 |
| Essential minerals | Na | Na | Essential minerals | Na | Na | -0.01 |
| Essential minerals | Se | Se | Essential minerals | Se | Se | 0.232 |
| Essential minerals | Zn | Zn | Essential minerals | Zn | Zn | 0.103 |
|  |  |  | Indoor air | BTEX | Indoor benzene | - |
|  |  |  | Indoor air | BTEX | Indoor BTEX | - |
|  |  |  | Indoor air | NO2 | Indoor NO2 | - |
|  |  |  | Indoor air | PM | Indoor PM2.5 | - |
|  |  |  | Indoor air | PM | Indoor PMabsorbance | - |
| Lifestyle | Diet | Cereals intake | Lifestyle | Diet | Cereals intake | 0.178 |
| Lifestyle | Diet | Fastfood intake | Lifestyle | Diet | Fastfood intake | 0.063 |
| Lifestyle | Diet | Fish and seafood intake | Lifestyle | Diet | Fish and seafood intake | 0.249 |
| Lifestyle | Diet | Fruits intake | Lifestyle | Diet | Fruits intake | 0.096 |
| Lifestyle | Diet | Meat intake | Lifestyle | Diet | Meat intake | 0.099 |
| Lifestyle | Diet | Vegetables intake | Lifestyle | Diet | Vegetables intake | -0.042 |
| Lifestyle | Diet | Dairy intake | Lifestyle | Diet | Dairy products intake | 0.077 |
| Lifestyle | Physical activity | Moderate physical activity (t3) | Lifestyle | Physical activity | Moderate and vigorous PA | 0.024 |
|  |  |  | Lifestyle | Diet | Bakery products intake | - |
|  |  |  | Lifestyle | Diet | Bread intake | - |
|  |  |  | Lifestyle | Diet | Breakfast cereals intake | - |
|  |  |  | Lifestyle | Diet | Breastfeeding duration (days) | - |
|  |  |  | Lifestyle | Diet | Caffeinated drinks | - |
|  |  |  | Lifestyle | Allergens | Cat at home | - |
|  |  |  | Lifestyle | Allergens | Dog at home | - |
|  |  |  | Lifestyle | Diet | KIDMED score | - |
|  |  |  | Lifestyle | Diet | Organic food intake | - |
|  |  |  | Lifestyle | Allergens | Other pets at home | - |
|  |  |  | Lifestyle | Diet | Potatoes intake | - |
|  |  |  | Lifestyle | Diet | Processed meat intake | - |
|  |  |  | Lifestyle | Diet | Ready made food intake | - |
|  |  |  | Lifestyle | Physical activity | Sedentary behaviour | - |
|  |  |  | Lifestyle | Sleep | Sleep duration | - |
|  |  |  | Lifestyle | Diet | Soda intake | - |
|  |  |  | Lifestyle | Diet | Sweets intake | - |
|  |  |  | Lifestyle | Diet | Total fat intake | - |
|  |  |  | Lifestyle | Diet | Yogurt intake | - |
| Metals | As | Arsenic | Metals | As | Arsenic | 0.206 |
| Metals | Cd | Cadmium | Metals | Cd | Cadmium | -0.053 |
| Metals | Cs | Cesium | Metals | Cs | Cesium | 0.462 |
| Metals | Hg | Mercury | Metals | Hg | Mercury | 0.467 |
| Metals | Pb | Lead | Metals | Pb | Lead | 0.216 |
| Metals | Tl | Thallium | Metals | Tl | Thallium | -0.006 |
| OCs | DDE | DDE | OCs | DDE | DDE | 0.632 |
| OCs | DDT | DDT | OCs | DDT | DDT | 0.345 |
| OCs | HCB | HCB | OCs | HCB | HCB | 0.016 |
| OCs | PCBs | PCB 118 | OCs | PCBs | PCB 118 | 0.258 |
| OCs | PCBs | PCB 138 | OCs | PCBs | PCB 138 | 0.344 |
| OCs | PCBs | PCB 153 | OCs | PCBs | PCB 153 | 0.314 |
| OCs | PCBs | PCB 170 | OCs | PCBs | PCB 170 | 0.283 |
| OCs | PCBs | PCB 180 | OCs | PCBs | PCB 180 | 0.38 |
| OP Pesticides | DEP | DEP | OP Pesticides | DEP | DEP | 0.016 |
| OP Pesticides | DETP | DETP | OP Pesticides | DETP | DETP | - |
| OP Pesticides | DMP | DMP | OP Pesticides | DMP | DMP | 0.079 |
| OP Pesticides | DMTP | DMTP | OP Pesticides | DMTP | DMTP | 0.025 |
|  |  |  | OP Pesticides | DMDTP | DMDTP | - |
| PBDEs | PBDE153 | PBDE 153 | PBDEs | PBDE153 | PBDE 153 | 0.085 |
| PBDEs | PBDE47 | PBDE 47 | PBDEs | PBDE47 | PBDE 47 | -0.029 |
| PFASs | PFHXS | PFHXS | PFASs | PFHXS | PFHXS | 0.486 |
| PFASs | PFNA | PFNA | PFASs | PFNA | PFNA | 0.152 |
| PFASs | PFOA | PFOA | PFASs | PFOA | PFOA | 0.199 |
| PFASs | PFOS | PFOS | PFASs | PFOS | PFOS | 0.457 |
| PFASs | PFUNDA | PFUNDA | PFASs | PFUNDA | PFUNDA | 0.222 |
| Phenols | BPA | BPA | Phenols | BPA | BPA | 0.058 |
| Phenols | BUPA | BUPA | Phenols | BUPA | BUPA | 0.029 |
| Phenols | ETPA | ETPA | Phenols | ETPA | ETPA | 0.068 |
| Phenols | MEPA | MEPA | Phenols | MEPA | MEPA | 0.045 |
| Phenols | OXBE | OXBE | Phenols | OXBE | OXBE | 0.19 |
| Phenols | PRPA | PRPA | Phenols | PRPA | PRPA | 0.031 |
| Phenols | TRCS | TRCS | Phenols | TRCS | TRCS | 0.09 |
| Phthalates | MBZP | MBzP | Phthalates | MBZP | MBzP | 0.157 |
| Phthalates | MECPP | MECPP | Phthalates | MECPP | MECPP | -0.032 |
| Phthalates | MEHHP | MEHHP | Phthalates | MEHHP | MEHHP | -0.037 |
| Phthalates | MEHP | MEHP | Phthalates | MEHP | MEHP | -0.033 |
| Phthalates | MEOHP | MEOHP | Phthalates | MEOHP | MEOHP | -0.06 |
| Phthalates | MEP | MEP | Phthalates | MEP | MEP | 0.112 |
| Phthalates | MIBP | MiBP | Phthalates | MIBP | MiBP | 0.105 |
| Phthalates | MNBP | MnBP | Phthalates | MNBP | MnBP | 0.065 |
| Phthalates | OHMiNP | OH-MiNP | Phthalates | OHMiNP | OH-MiNP | 0.031 |
| Phthalates | OXOMINP | oxo-MiNP | Phthalates | OXOMINP | oxo-MiNP | 0.024 |
|  |  |  | Social and economic capital | Social capital | Contact with family and friends | - |
|  |  |  | Social and economic capital | Economic capital | Family affluence score | - |
|  |  |  | Social and economic capital | Social capital | House crowding | - |
|  |  |  | Social and economic capital | Maternal stress | Maternal stress | - |
|  |  |  | Social and economic capital | Social capital | Social participation | - |
| Tobacco Smoke | Cotinine | Cotinine | Tobacco Smoke | Cotinine | Cotinine | 0.410 |
| Tobacco Smoke | Tobacco Smoke | Maternal smoking (active and ETS) | Tobacco Smoke | Tobacco Smoke | Parental smoking | 0.36 |
|  |  |  | Tobacco Smoke | Tobacco Smoke | ETS | - |
| Built Environment | Facility | Facility richness (300m) |  |  |  | - |
| Meteorological | Pressure | Pressure (t1) |  |  |  | - |
| Water DBPs | Water DBPs | Water Brominated THMs |  |  |  | - |
| Water DBPs | Water DBPs | Water Chloroform |  |  |  | - |
| Water DBPs | Water DBPs | Water THMs |  |  |  | - |
| Lifestyle | Physical activity | Vigorous physical activity (t3) |  |  |  | - |
| Lifestyle | Diet | Legumes intake |  |  |  | - |
| Lifestyle | Folic acid consumption | Folic acid supplementation |  |  |  | - |
| Lifestyle | Prenatal Alcohol | Alcohol intake |  |  |  | - |

***Child molecular phenotypes***

Several child molecular phenotypes were measured as described in **Table S3**, and described below.

| **Table S3. Number of children and number of molecular features per omics, before and after quality control** | | | | | |
| --- | --- | --- | --- | --- | --- |
| Omics | Method | N features initial^a^ | N features final | N samples initial | N samples final |
| Blood DNA methylation | 450K, Illumina | 485512 | 386518 | 1180 | 1173 |
| Blood gene expression | HTA v2.0, Affymetrix | 70523 | 58254 | 1024 | 1007 |
| Blood miRNA expression | SurePrint Human miRNA rel 21, Agilent | 2570 | 1117 | 947 | 941 |
| Plasma proteins | 3 kits, Luminex | 43 | 36 | 1190 | 1170 |
| Serum metabololites | AbsoluteIDQ p180 kit, Biocrates | 188 | 177 | 1199 | 1198 |
| Urinary metabololites | ^1^H NMR | 36K | 44 | 1198 | 1198 |
| ^a^Number of initial features in the arrays, including control probes, in the commercial kits or in the untargetted ^1^H NMR assay | | | | | |

***Blood DNA methylation***

DNA was obtained from children’s peripheral blood (buffy coat) collected in EDTA tubes. DNA was extracted using the Chemagen kit (Perkin Elmer, USA) in batches of 12 samples within each cohort. DNA concentration was determined in a Nanodrop 1000 UV-Vis Spectrophotometer (Thermo Fisher Scientific, USA) and also with Quant-iT^TM^ PicoGreen^®^ dsDNA Assay Kit (Life Technologies, USA). DNA extraction was repeated in around 8% of the blood samples as the DNA quantity or quality of the first extraction was low. Less than 1.5% of the samples were finally excluded due to low quality.

DNA methylation was assessed with the Infinium HumanMethylatio450 beadchip (Illumina, USA) at the University of Santiago de Compostela – Spanish National Genotyping Center (CeGen-USC) (Spain). 700 ng of DNA were bisulfite-converted using the EZ 96-DNA kit (Zymo Research, USA) following the manufacturer’s standard protocol. All samples of the study were randomized considering sex and cohort. In addition, each plate contained a HapMap control sample and 24 HELIX inter-plate duplicates were included.

After an initial inspection of the quality of the methylation data with the MethylAid package (van Iterson et al., 2014), probes with a call rate <95% based on a detection p-value of 1E-16 and samples with a call rate <98% were removed (Lehne et al., 2015). Samples with discordant sex were eliminated from the study as well as duplicates with inconsistent genotypes and samples with inconsistent genotypes respect to existing genome-wide genotyping array data. Methylation data was normalized using the functional normalization method with prior background correction with Noob (Fortin et al., 2014). Then, some probes were filtered out: control probes, probes to detect single nucleotide polymorphisms (SNPs), probes to detect methylation in non-CpG sites, probes located in sexual chromosomes, cross hybridizing probes (Chen et al., 2013), probes containing a SNP at any position of the sequence with a minor allele frequency (MAF) >5% and probes with a SNP at the CpG site or at the single base extension (SBE) at any MAF in the combined population from 1000 Genomes Project. Batch effects and blood cell composition were corrected by calculating 134 surrogate variables while protecting cohort, sex and age with the surrogate variable analysis (SVA) method (Leek and Storey, 2007), and residualizing them on the methylation data. CpGs were annotated with the IlluminaHumanMethylation450kanno.ilmn12.hg19 R package (Hansen. K. D., 2012).

***Blood gene expression***

RNA was extracted from whole blood samples collected in Tempus tubes (Thermo Fisher Scientific, USA) using MagMAX for Stabilized Blood Tubes RNA Isolation Kit. The quality of RNA was evaluated with a 2100 Bioanalyzer (Agilent Technologies, USA) and the concentration with a NanoDrop 1000 UV-Vis Spectrophotometer. Samples classified as good RNA quality (78.67%) had a similar RNA pattern at visual inspection in the Bioanalyzer, a RNA Integrity Number (RIN) >5 and a concentration >10 ng/ul. Mean values (standard deviation, SD) for the RIN, concentration (ng/ul), Nanodrop 260/280 ratio and Nanodrop 260/230 ratio were: 7.05 (0.72), 109.07 (57.63), 2.15 (0.16) and 0.61 (0.41).

Gene expression was assessed using the GeneChip^®^ Human Transcriptome Array 2.0 (HTA 2.0) from Affymetrix (USA) at the University of Santiago de Compostela (USC) (Spain). Briefly, RNA samples were concentrated or evaporated in order to reach the required RNA input concentration (200 ng of total RNA). Amplified and biotinylated sense-strand DNA targets were generated from total RNA. Microarrays were hybridized according to the Affymetrix recommendations using the Affymetrix labeling and hybridization kits. All samples were randomized within each batch considering sex and cohort. Two different types of control RNA samples (HeLa and FirstChoice^®^ Human Brain Reference RNA (Thermo Fisher Scientific, USA)) were included in each batch, but they were hybridized only in the first batches.

Raw data were extracted with the Affymetrix AGCC software and normalized with the GCCN (SST-RMA) algorithm at the gene level ([http://tools.thermofisher.com/content/sfs/brochures/ sst_gccn_whitepaper.pdf](http://tools.thermofisher.com/content/sfs/brochures/%20sst_gccn_whitepaper.pdf)). Gene expression values were log2 transformed. Annotation of transcripts clusters (TCs) to genes was done with the Affymetrix ExpressionConsole software using the HTA-2_0 Transcript Cluster Annotations Release na36 (hg19). A transcript cluster is defined as a group of one or more probes covering a region of the genome reflecting all the exonic transcription evidence known for the region and corresponding to a known or putative gene. Four samples with discordant sex were detected with the MassiR R package (Buckberry et al., 2014) and excluded. Control probes, and TCs in sexual chromosomes and without chromosome information were filtered out. Batch effects and blood cell composition were corrected using 71 surrogate variables calculated with the SVA method (Leek and Storey, 2007). In order to determine TC call rate, 10 constitutive or best probes based on probe scoring and cross-hybridation potential were selected per TC. Probe Detection Above Background (DABG) p-values were computed based on the rank order against the background probe set intensities. Probe level p-values were combined into a TC level p-value using the Fisher equation. TCs with a DABG p-value <0.05 were defined as detected. Three samples with low call rate (<40%) as well as TCs with a call rate <1% were excluded from the dataset.

***Blood miRNA expression***

RNA was extracted from whole blood as detailed in the previous section. MiRNA expression was quantified using the SurePrint Human miRNA Microarray rel.21 (Agilent Technologies, USA), which evaluates >2000 miRNA from miRbase v21 (Kozomara and Griffiths-Jones, 2011), at the Genomics Core facility at the Centre for Genomic Regulation (CRG) (Spain). After sample randomization by sex and cohort, 24 samples were processed per batch and hybridized in 3 different slides (8 samples per slide). A control sample, a total RNA mixture from 9 human tissues or cell lines (universal miRNA reference kit (Agilent Technologies, USA)), was included in 2/3 of the batches with a total of 39 control samples. HELIX RNA samples were concentrated or evaporated using a SpeedVac in order to reach the required RNA input (100 ng of total RNA). Fluorescently-labeled miRNA, obtained with the miRNA Complete Labeling and Hybridization kit, was hybridized onto the microarrays according to Agilent recommendations. Raw data were extracted with the Agilent Feature extraction software. Seventeen samples did not reach the laboratory quality control parameters and were processed again. Six of them were finally removed due to overall low quality.

After testing different normalization methods using the 39 control RNA samples, miRNA expression levels were normalized with the Least Variant Set (LVS) method (Suo et al., 2010) with background correction with the Normexp method implemented in the limma package (Ritchie et al., 2015). LVS normalization method builds upon the fact that the data-driven housekeeping miRNAs that are the least variant across samples might be a good reference set for normalization. For the identification of housekeeping miRNAs, a random set of 50 HELIX samples was used. Normalization using the selected miRNA reference set was done by applying the variance stabilization and calibration for microarray data (vsn) method(Huber et al., 2002). Normalized miRNA levels were log2 transformed and annotated using a combination of information from Agilent annotation (“Annotation_7056”) and miRbase v21 (GRCh38 and mapped back to hg19) released in January 2017 (“annotation_miRBase_GRCh38_coordinates-gff3”). Then control probes, miRNAs in sexual chromosomes and unannotated miRNAs were excluded from the database. Batch effects and blood cell composition was corrected as described above with a total of 35 surrogate variables. Finally, miRNAs with a call rate <1% were also filtered out. A miRNA was considered as detected if its expression was different from the background or the standard error of its different probes was smaller than 3 times the expression signal. After exclusion of control samples and control probes, the average number of detected miRNAs was 420.43 (minimum: 71; maximum: 940).

***Plasma proteins***

Plasma protein levels were assessed using the antibody-based multiplexed platform from Luminex. Three kits targeting 43 unique candidate proteins were selected (Thermo Fisher Scientifics, USA): Cytokines 30-plex (Catalog Number (CN): LHC6003M), Apoliprotein 5-plex (CN: LHP0001M) and Adipokine 15-plex (CN: LHC0017M).

All samples were randomized and blocked by cohort prior measurement. For quantification, an 8-point calibration curve per plate was performed with protein standards provided in the Luminex kit and following procedures described by the vendor. Commercial heat inactivated, sterile-filtered plasma from human male AB plasma (Sigma-Aldrich, USA) was used as constant samples to control for intra- and inter-plate variability. Four control samples were added per plate. All samples, including controls, were diluted ½ for the 30-plex kit, ¼ for the 15-plex kit and 1/2500 for the 5-plex kit.

Raw intensities obtained with the xMAP and Luminex system for each plasma sample were converted to ng/ml (5-plex kit) and to pg/ml (15 and 30-plex kits) using the calculated standard curves of each plate and accounting for the dilutions made prior measurement. The percentages of coefficients of variation (CV%) for each protein by plate ranged from 3% to 36%. The limit of detection (LOD) and the lower and upper limit of quantification (LOQ1 and LOQ2, respectively) were estimated by plate, and then averaged. Only proteins with >30% of measurements in the linear range of quantification were kept in the database and the others were removed. Seven proteins were measured twice (in two different multiplex kits). We kept the measure with higher quality. The 36 proteins that passed the quality control criteria mentioned above were log2 transformed (Vives-Usano et al., 2020). Then, the plate batch effect was corrected by subtracting the plate specific average for each protein minus the overall average of all plates for that protein. After that, values below the LOQ1 and above the LOQ2 were imputed using a truncated normal distribution implemented in the truncdist R package (Nadarajah and Kotz, 2006). Twenty samples were excluded due to having ten or more proteins out of the linear range of quantification.

***Serum metabolites***

The AbsoluteIDQTM p180 kit was chosen for serum analysis as it is a standardised, targeted LC-MS/MS assay, widely used for large-scale epidemiology studies and its inter-laboratory reproducibility has been demonstrated by several independent laboratories (Siskos et al., 2017).Serum samples were quantified using the AbsoluteIDQTM p180 kit following the manufacturer’s protocol (User Manual UM_p180_AB_SCIEX_9, Biocrates Life Sciences AG) using LC-MS/MS; an Agilent HPLC 1100 liquid chromatography coupled to a SCIEX QTRAP 6500 triple quadrupole mass spectrometer. A full description of the HELIX metabolomics methods and data can be found elsewhere (Lau et al., 2018).

Briefly, the kit allows for the targeted analysis of 188 metabolites in the classes of amino acids, biogenic amines, acylcarnitines, glycerophospholipids, sphingolipids and sum of hexoses, covering a wide range of analytes and metabolic pathways in one targeted assay. The kit consists of a single sample processing procedure, with two separate analytical runs, a combination of liquid chromatography (LC) and flow injection analysis (FIA) coupled to tandem mass spectrometry (MS/MS). Isotopically labelled and chemically homologous internal standards were used for quantification. The AbsoluteIDQ p180 data of serum samples were acquired in 18 batches. Every analytical batch, in a 96-well plate format, included up to 76 randomised cohort samples. Also in every analytical batch, three sets of quality control samples were included, the NIST SRM 1950 plasma reference material (in 4 replicates), a commercial available serum QC material (CQC in 2 replicates, SeraLab, S-123-M-27485) and the QCs provided by the manufacturer in three concentration levels. The NIST SRM 1950 reference was used as the main quality control sample for the LC-MS/MS analysis. Coefficients of variation (CVs) for each metabolite were calculated based on the NIST SRM 1950 and also the limits of detection (LODs) were also used to assess the analytical performance of individual metabolites. Metabolite exclusion was based on a metabolite variable meeting two conditions: (1) CV of over 30% and (2) over 30% of the data are below LOD. Eleven out of the 188 serum metabolites detected were excluded as a result, leaving 177 serum metabolites to be used for further statistical analysis. The mean coefficient of variation across the 177 LC-MS/MS detected serum metabolites was 16%. We also excluded one HELIX sample, which was hemolyzed. Concentration levels were log2 transformed.

***Urinary metabolites***

Two urine samples, representing last night-time and first morning voids, were collected on the evening and morning before the clinical examination, kept in a fridge and transported in a temperature-controlled environment, and aliquoted and frozen within 3 h of arrival at the clinics. They were subsequently pooled to generate a more representative sample of the last 24 h for metabolomic analysis.

Urinary metabolic profiles were acquired using ^1^H NMR spectroscopy according to (Lau et al., 2018). In brief one-dimensional 600 MHz ^1^H NMR spectra of urine samples from each cohort were acquired on the same Bruker Avance III spectrometer operating at 14.1 Tesla within a period of 1 month. The spectrometer was equipped with a Bruker SampleJet system, and a 5-mm broad-band inverse configuration probe maintained at 300K. Prior to analysis, cohort samples were randomised. Deuterated 3-(trimethylsilyl)-[2,2,3,3-d4]-propionic acid sodium salt (TSP) was used as internal reference. Aliquots of the study pooled quality control (QC) sample were used to monitor analytical performance throughout the run and were analysed at an interval of every 23 samples (i.e. 4 QC samples per well plate). The ^1^H NMR spectra were acquired using a standard one-dimensional solvent suppression pulse sequence. 44 metabolites were identified and quantified as described (Lau et al., 2018). The urinary NMR showed excellent analytical performance, the mean coefficient of variation across the 44 NMR detected urinary metabolites was 11%. Data was normalized using the median fold change normalization method (Dieterle et al., 2006), which takes into account the distribution of relative levels of all 44 metabolites compared to the reference sample in determining the most probable dilution factor. An offset of ½ of the minimal value was applied and then concentration levels were expressed as log2.

***Statistical analysis (exposome-omics-wide association study - ExWAS)***

To test the relationship between the pregnancy and childhood exposomes and molecular features, we fitted linear regressions between each exposure variable and each molecular feature adjusting for covariates, using the limma R package (Ritchie et al., 2015) implemented in omicRexposome (Hernandez-Ferrer et al., 2019). The MultiDataSet (Hernandez-Ferrer et al., 2017) and rexposome (Hernandez-Ferrer et al., 2019) R packages were used to manage the omics and exposure data.

Main covariates for all omics were: cohort, child’s sex, child’s age, child sex and age z-score BMI calculated according to WHO reference curves (de Onis et al., 2007; 2015), child’s ethnicity defined in three categories (European ancestry; Pakistani or Asian; and other), and self-reported maternal education categorized in low (primary school), medium (secondary school) and high (university degree or higher). Ethnic origin (African, Asian, white European, Mexican, and other) was asked to the families as part of the spirometry protocol. This information was combined with existing data (parent’s ethnic origin and/or parent’s country of birth) in each of the cohorts. Missing data for covariates was imputed as described above for the exposome.

In addition, models for each omics were adjusted for specific covariates: (i) plasma protein models were adjusted for time to last meal and hour of blood collection, (ii) serum metabolite models for time to last meal, hour of blood collection, and technical batch, (iii) urinary metabolite models for sample type (bedtime, morning or pool), and technical batch. Blood methylation and transcriptomics data were corrected by surrogate variables as described above, which captured both batch effects and blood cell type composition.

In all omics, except for methylation, the effect size is reported as a log2 fold change (log2FC) of the molecular phenotype levels between categories of discrete exposure variables or for interquartile range (IQR) of continuous exposure variables. For DNA methylation, the effect size is reported as a difference in methylation levels between categories of discrete exposure variables or for IQR of continuous exposure variables.

P-values were corrected within each omics layer by the number of molecular features of a given omics platform. For methylation, gene expression and miRNAs we used the False Discovery Rate (FDR) - Benjamini Hochberg (BH) (Benjamini and Hochberg, 1995). For other omics we calculated the effective number of tests (ENT) (Li et al., 2012), and divided the nominal p-values (0.05) by that number. We also calculated a more stringent threshold correcting for all tests performed (all molecular features across platforms and the full exposome), giving a p-value of 1E-09. Associations reaching this p-value are flagged in the tables as “BN”. The lambda inflation factor distribution for DNA methylation and gene expression ranged from 0.9 to 1.11.

Finally, a set of sensitivity analyses were conducted. First, models were run again without adjustment for child zBMI. The difference in the effect size among main models and zBMI unadjusted models was calculated as (effect size main model – effect size alternative model) / effect size main model *100. Second, analyses were restricted to children of European ancestry (90%). Third, top hit associations were run by cohort and combined through fixed- and random-effects inverse variance weighted meta-analyses using the meta R package (Schwarzer, 2007), and forest-plots were visually inspected. I^2^ was used to evaluate heterogeneity in the results across cohorts.

Significant associations were represented in a variety of ways: Miami-plots by family of exposure, scatter-plots of the effect size of two different models, heatmaps, and forest-plots by cohort. These plots were created using ggplot2 (2018), qqman (Stephen Turner and Stephen Turner, 2017), calibrate (Jan Graffelman and Jan Graffelman, 2015) and pheatmap R packages, among others.

***Network analyses***

Networks visualization was carried out using Cytoscape 3.6.1 (http://cytoscape.org). The two networks, one for pregnancy and another for childhood exposures, were automatically arranged using the Cytoscape force-directed layout which aims to highlight the underlying topology of the graph (Shannon et al., 2003). Clustering of the childhood network was done based on Community Clustering (GLay) in order to find densely connected regions in the network (Newman and Girvan, 2003; Su et al., 2010). Node attributes were calculated using the network analyzer tool and presented in Tables S2A (pregnancy) and S2B (childhood). The NetworkAnalyzer plugin and a comprehensive online documentation with a tutorial are available at http://med.bioinf.mpi-inf.mpg.de/networkanalyzer/.

***Comparison with the EWAS catalogue and EWAS atlas***

We evaluated the overlap between significant CpGs per exposure or per cluster, with CpGs associated with traits or exposure retrieved from the EWAS Catalog (<http://ewascatalog.org/>, release 23/11/2018, 542,319 entries from 160 publications, covering 165 traits and 58 tissues) and the EWAS Atlas (<http://bigd.big.ac.cn/ewas/index>, release 13/05/2019, 590,007 entries, 519 publications, 370 traits and 133 tissues).

***Comparison with Exposome Explorer – dietary biomarker database***

The overlap between metabolites in urine and serum samples associated with exposures in HELIX samples with dietary intake based on the Exposome Explorer database (<http://exposome-explorer.iarc.fr/>, release 2019-09-06) was checked (Neveu et al., 2017). “Biomarker” and “Concentrations” datasets were downloaded and “Metabolomic associations”, linking metabolites with dietary intakes based on currated literature, were manually compiled from the web page. Our urine and serum metabolite datasets were merged with the ExposomeExplorer database based on HMDB identifiers. Tripartite plots were drawn based on the presence of associations between metabolites-exposure in HELIX samples and metabolites-dietary intake based on ExposomeExplorer database.

***Functional enrichment analyses***

Functional enrichment analyses were restricted to molecular layers with features which could be easily annotated at the gene level: DNA methylation, gene and miRNA transcription, and proteins. Enrichment was investigated using a variety of databases following the same workflow. We annotated molecular features to genes within each pregnancy and childhood cluster and performed functional enrichment analyses. Second, for pregnancy and childhood exposures with at least one significant association with molecular features after multiple-testing correction, we retrieved all molecular features associated at p-value <1E-03. After that, we annotated these features to genes as described in the omics data acquisition sections, and obtained a unique list of “dysregulated” genes by combining genes detected in any of the molecular layers. The gene universe was the list of genes in the methylation study, annotated with the *IlluminaHumanMethylation450kanno.ilmn12.hg19* (N=21,235).

ClusterProfiler R package (Yu, 2018) was used to perform over-representation analyses to assess enrichment for gene-sets (Gene Ontology (GO) Biological Processes terms, KEGG, Molecular Signatures Database - C2 curated gene-sets), diseases (DisGeNET) and transcription factor and miRNA binding motifs (Molecular Signatures Database - C3 motifs and transcription factors motifs). Multiple-testing was corrected with the FDR - BN method within each exposure and only gene-sets with >3 genes are reported. For visualization in plots, GO terms were pruned based on their similarity using the REVIGO tool (<http://revigo.irb.hr/>) with the following parameters: Select a semantic similarity measure to use = SimRel, and Allow similarity = Small.

***Expression quantitative trait methylation (eQTMs) and miRNA gene target prediction***

We used miRwalk v3 to explore miRNA targeted genes (Sticht et al., 2018). We applied the following filters: humans, only 3’UTR region, score 0.95 (default), experimentally validated interaction described in miRTarBase (release 7.0).

To test associations between DNA methylation levels and gene expression levels in cis (cis-eQTMs), we paired each transcript cluster (TC) to all CpGs closer than 500 kb from its TSS, either upstream or downstream. For each CpG-TC pair we fitted a linear regression model between gene expression and methylation levels adjusted for age, sex and cohort. Methylation slide batch effects were controlled using ComBat, while for gene expression we eliminated the effect of surrogate variables protecting covariates in the model (age, sex and cohort) as well as blood cell type proportions. The analysis was restricted to children of European ancestry and autosomal chromosomes. To ensure that CpGs paired to a higher number of TCs do not have higher chances of being part of an eQTM, multiple-testing was controlled at the CpG level. More details on the analyses and the multiple-testing correction can be found at <https://www.biorxiv.org/content/10.1101/2020.11.05.368076v1>.
