## Supplementary material for "Multi-omics signatures of the human early life exposome": Figure S2

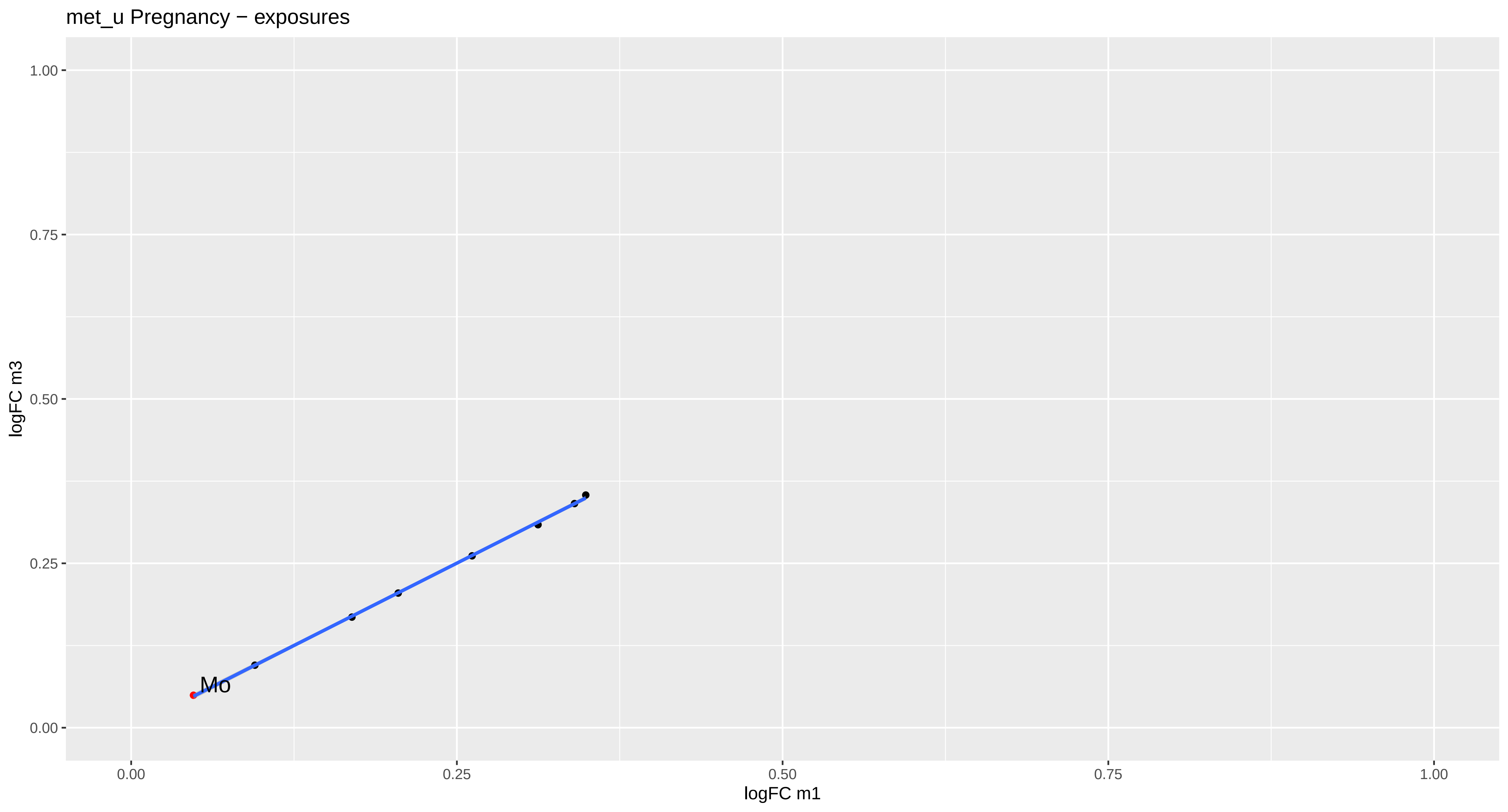
met_u Pregnancy − exposures
logFC m3
1.00
0.75
0.50
0.25
0.00
Mo
0.00
0.25
0.50
0.75
1.00
logFC m1

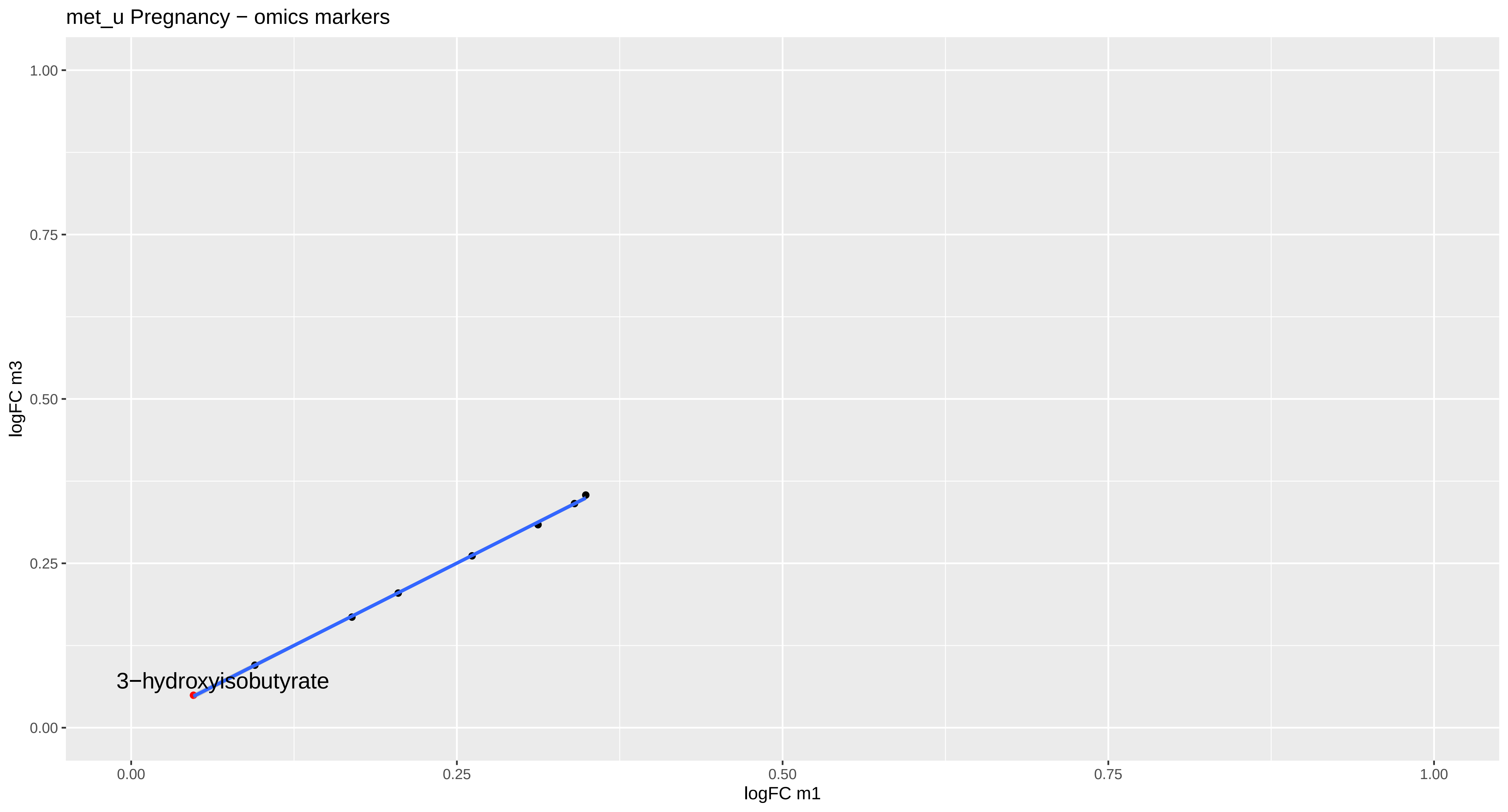
met_u Pregnancy − omics markers
logFC m3
1.00
0.75
0.50
0.25
0.00
3−hydroxyisobutyrate
0.00
0.25
0.50
0.75
1.00
logFC m1

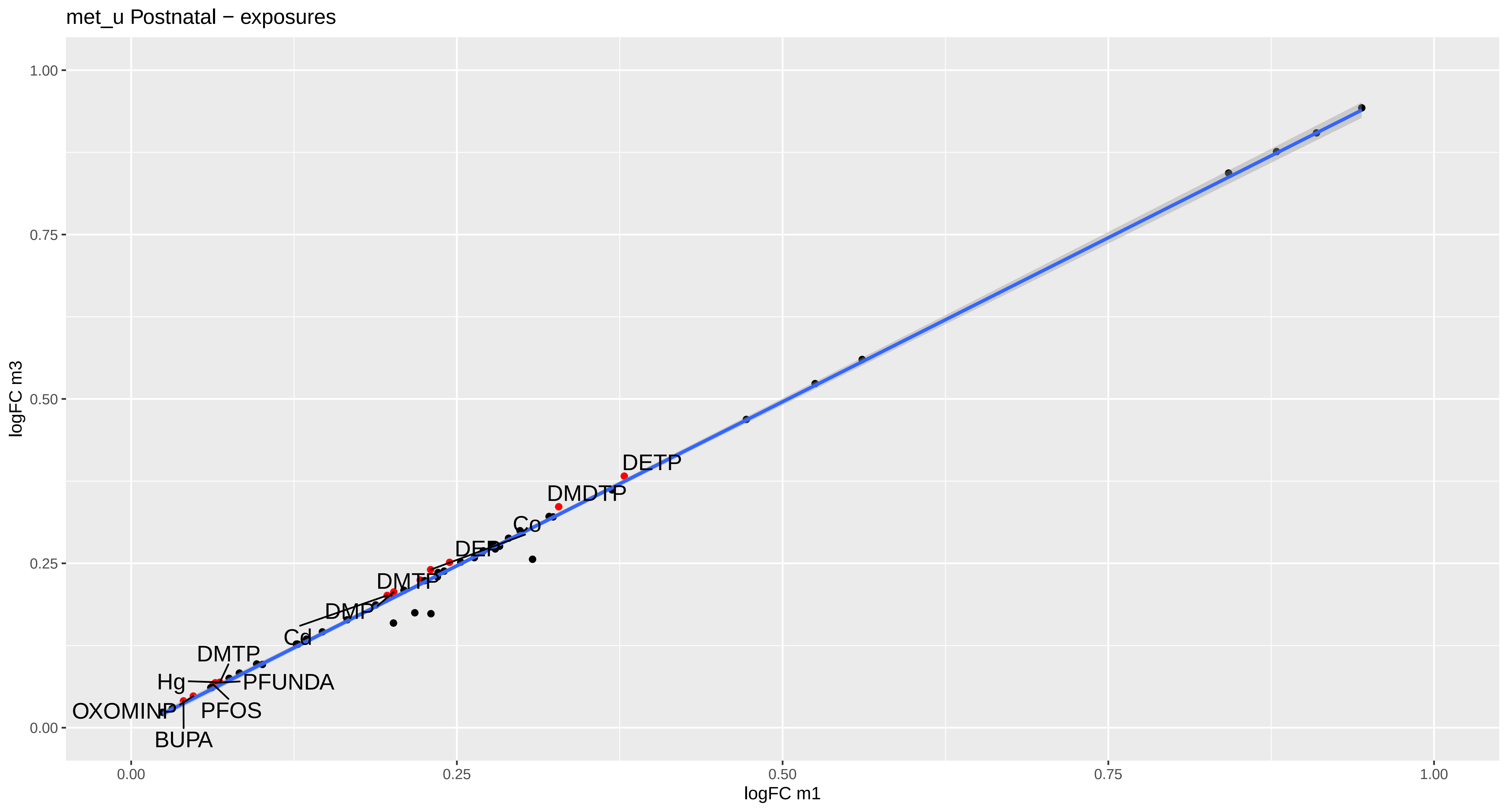

met_u Postnatal − exposures
logFC m3
logFC m1
1.00
0.75
0.50
0.25
0.00
DETP
DMDTP
Co
DEP
DMTP
DMP
Cd
DMTP
Hg
PFUNDA
PFOS
OXOMINP
BUPA

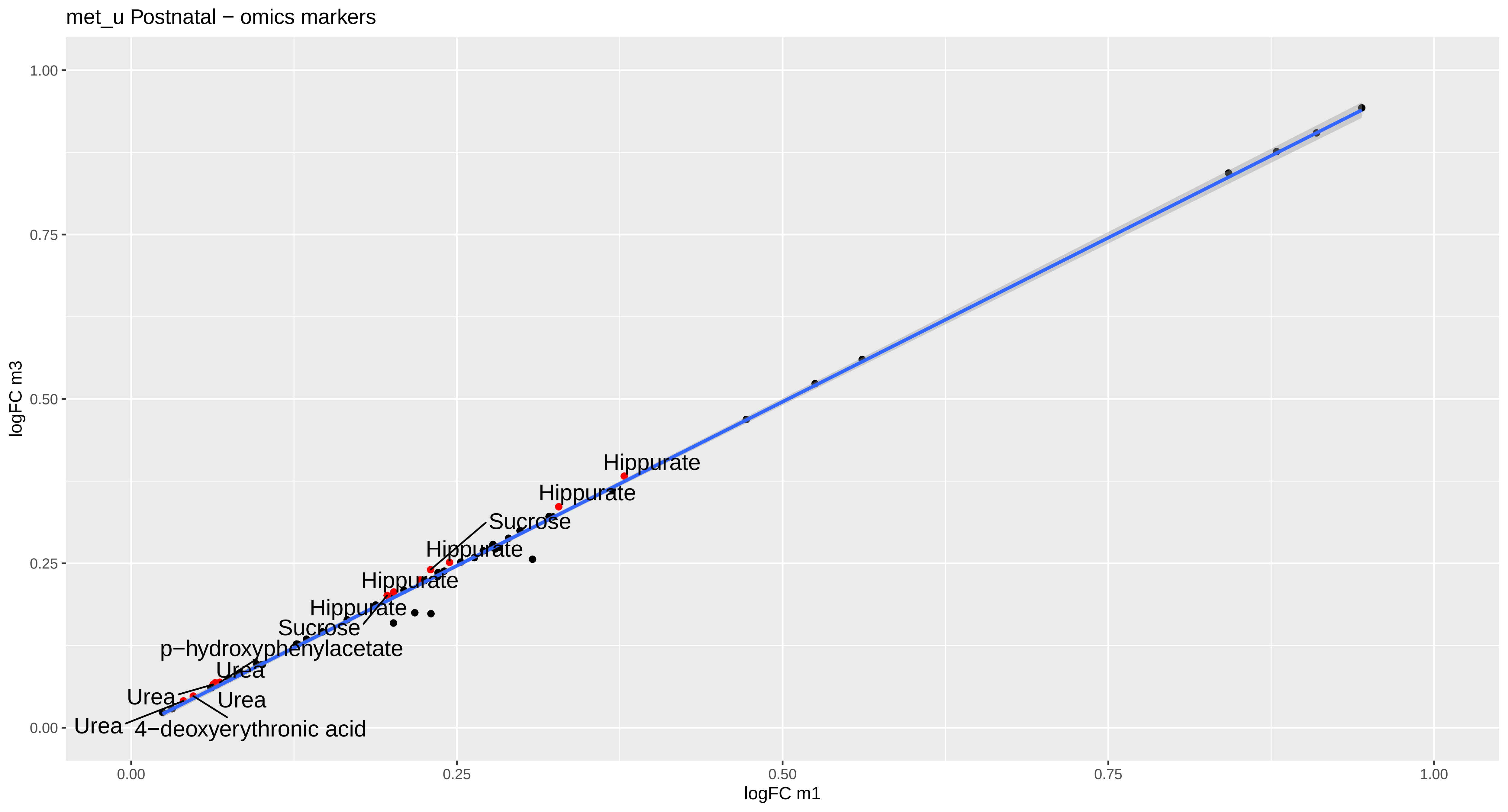

met_u Postnatal − omics markers
logFC m3
1.00
0.75
0.50
0.25
0.00
logFC m1
0.00
0.25
0.50
0.75
1.00
Hippurate
Hippurate
Sucrose
Hippurate
Hippurate
Hippurate
Sucrose
p−hydroxyphenylacetate
Urea
Urea
Urea
Urea
4−deoxyerythronic acid

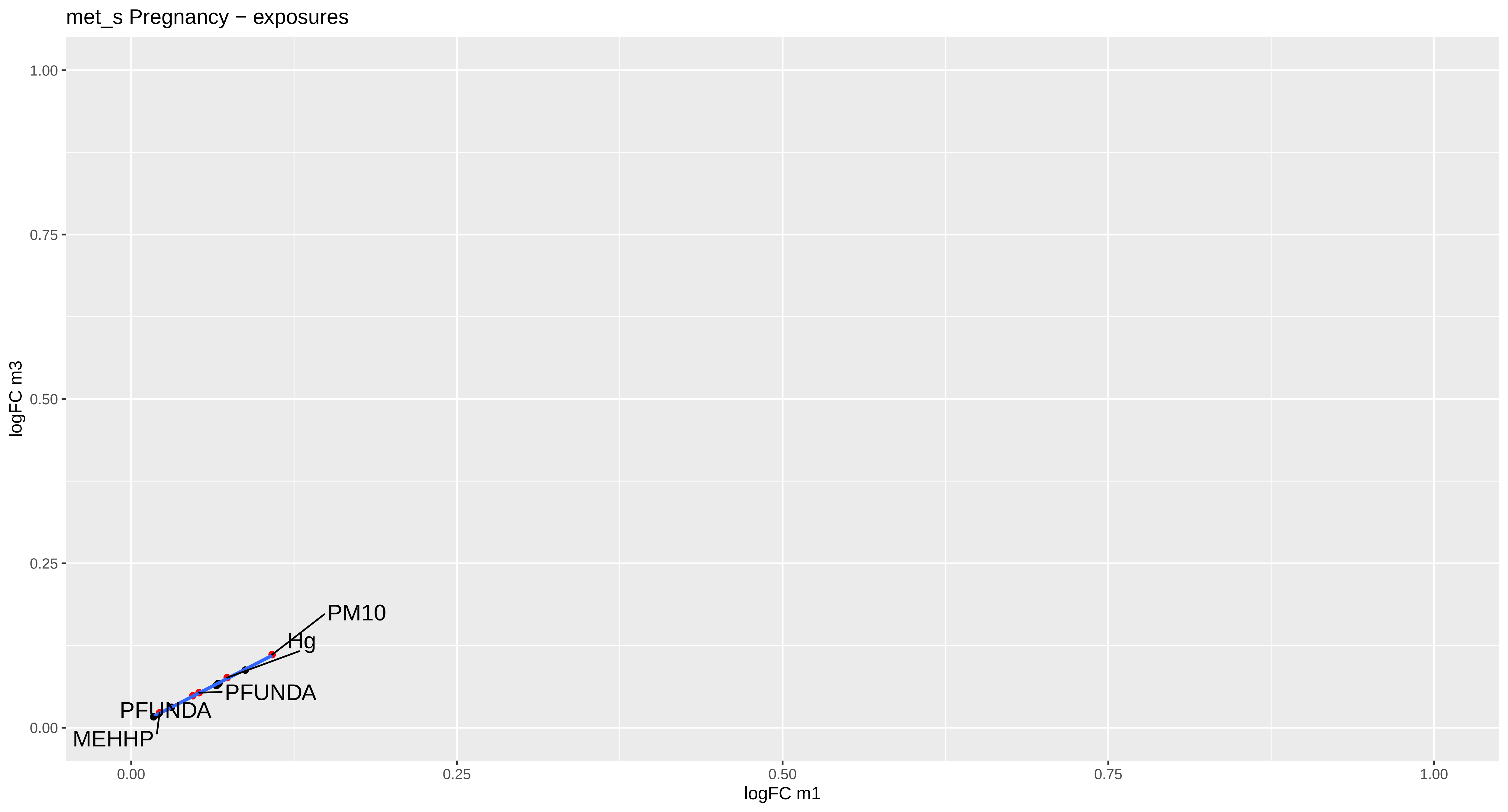

met_s Pregnancy − exposures
logFC m3
logFC m1
1.00
0.75
0.50
0.25
0.00
0.00
0.25
0.50
0.75
1.00
PM10
Hg
PFUNDA
PFUNDA
MEHHP

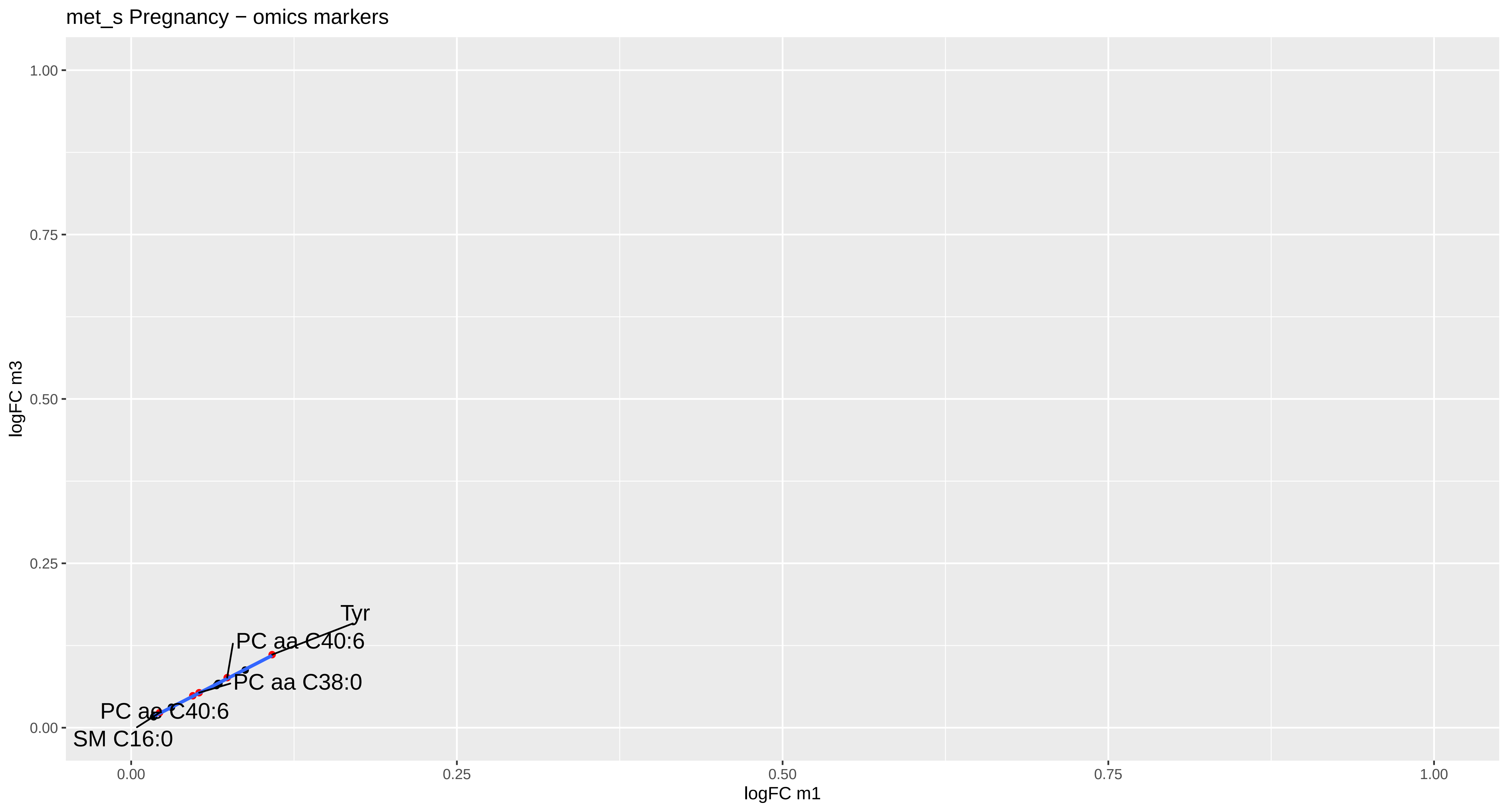
met_s Pregnancy − omics markers
1.00
0.75
0.50
0.25
0.00
logFC m3
Tyr
PC aa C40:6
PC aa C38:0
PC ae C40:6
SM C16:0
0.00
0.25
0.50
0.75
1.00
logFC m1

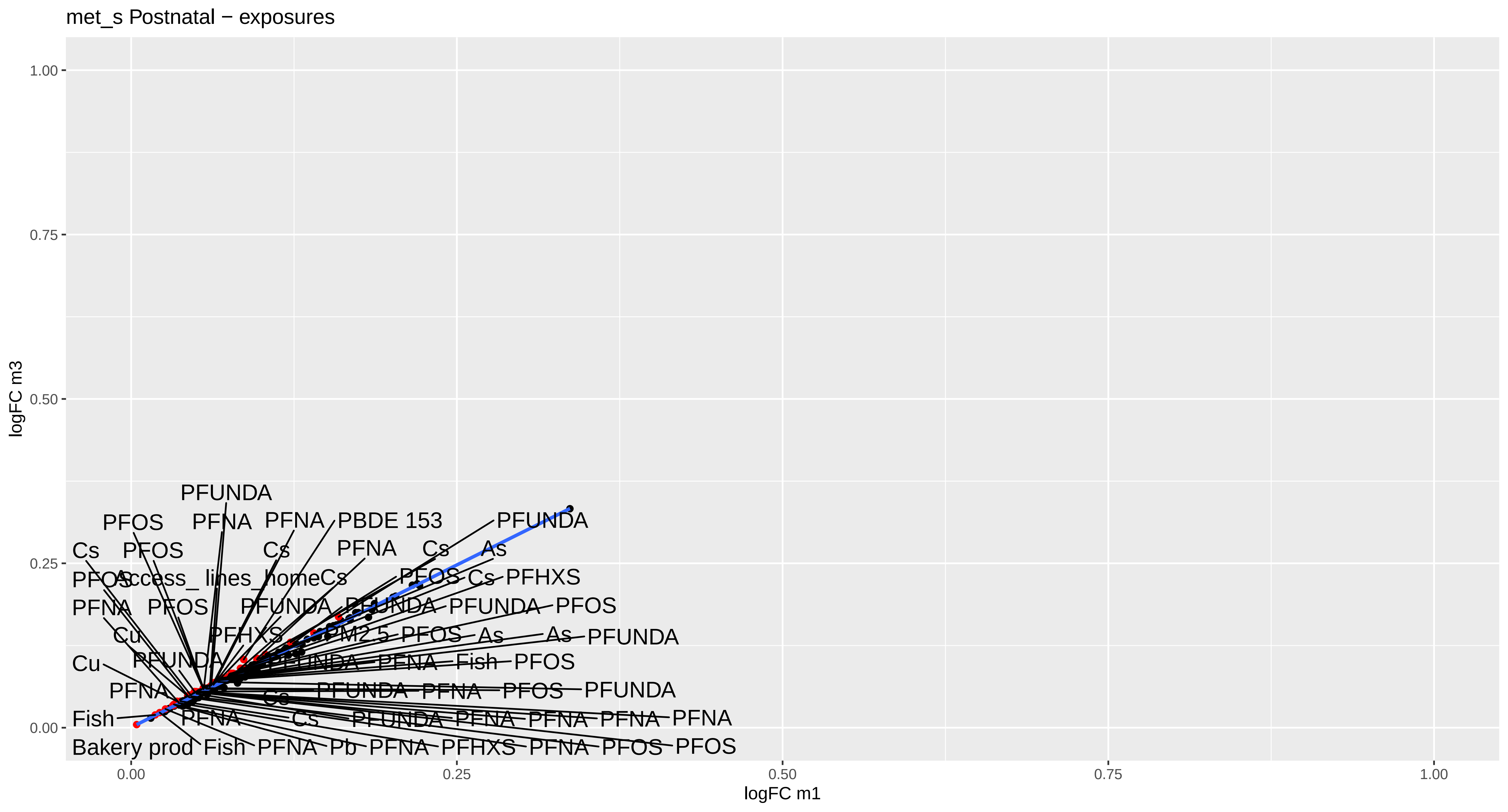

met_s Postnatal − exposures
logFC m3
logFC m1
0.00
0.25
0.50
0.75
1.00
PFUNDA
PFOS
PFNA
PFNA
PBDE 153
PFUNDA
Cs
PFOS
Cs
PFNA
Cs
As
Access_ lines_homeCs
PFOS
Cs
PFHXS
PFNA
PFOS
PFUNDA
PFUNDA
PFUNDA
PFOS
Cu
PFHXS
PM2.5
PFOS
As
As
PFUNDA
Cu
PFUNDA
PFUNDA
PFNA
Fish
PFOS
PFNA
Cs
PFUNDA
PFNA
PFOS
PFUNDA
Fish
PFNA
Cs
PFUNDA
PFNA
PFNA
PFNA
PFNA
Bakery prod
Fish
PFNA
Pb
PFNA
PFHXS
PFNA
PFOS
PFOS

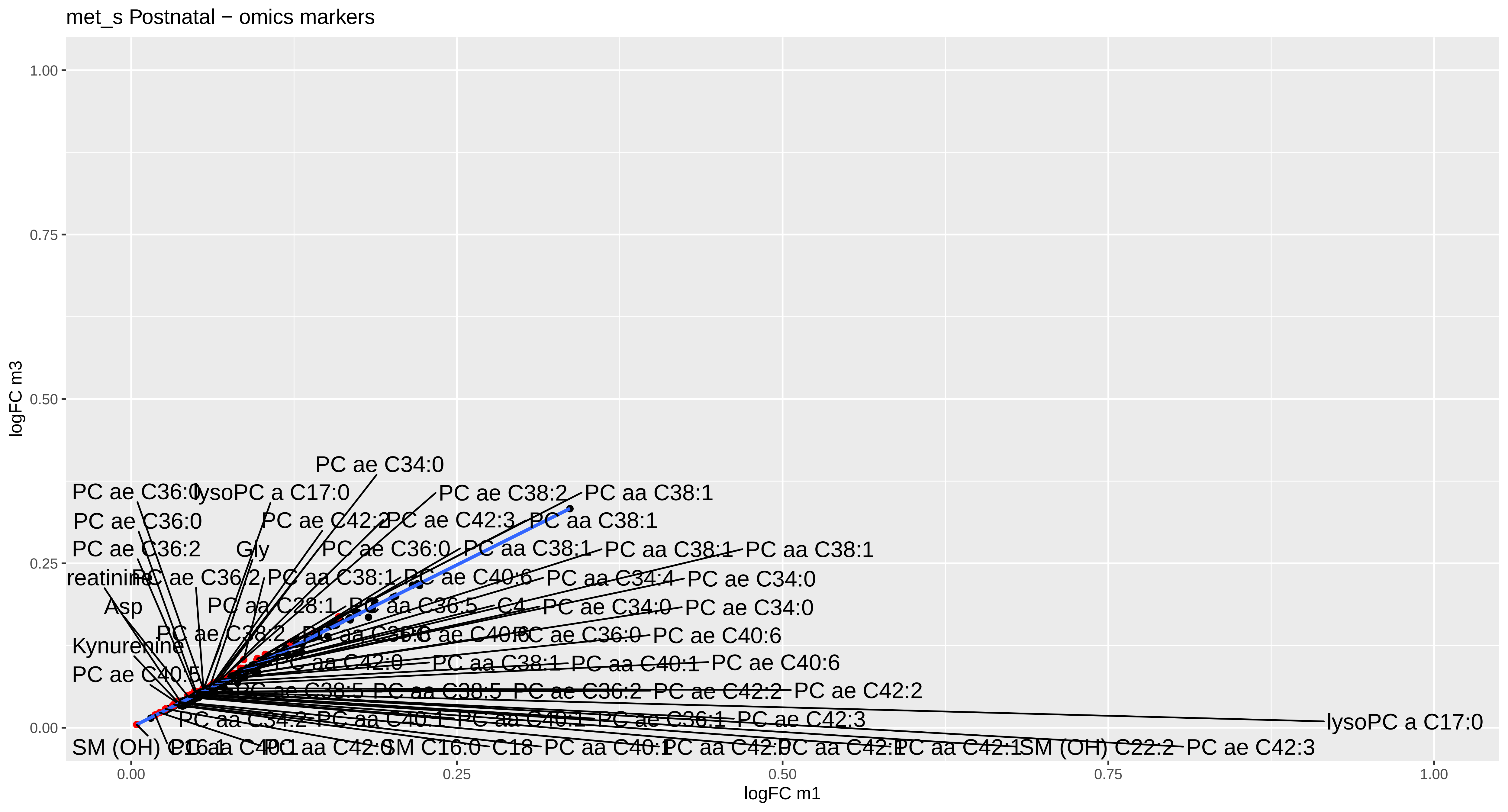
met_s Postnatal − omics markers
logFC m3
logFC m1
1.00
0.75
0.50
0.25
0.00
0.00
0.25
0.50
0.75
1.00
PC ae C34:0
PC ae C36:0
lysoPC a C17:0
PC ae C38:2
PC aa C38:1
PC ae C36:0
PC ae C42:2
PC ae C42:3
PC aa C38:1
PC ae C36:2
Gly
PC ae C36:0
PC aa C38:1
PC aa C38:1
PC aa C38:1
reatinine
PC ae C36:2
PC aa C38:1
PC ae C40:6
PC aa C34:4
PC ae C34:0
Asp
PC aa C28:1
PC aa C36:5
C4
PC ae C34:0
PC ae C34:0
PC ae C38:2
PC ae C40:6
PC ae C36:0
PC ae C40:6
Kynurenine
PC aa C42:0
PC aa C38:1
PC aa C40:1
PC ae C40:6
PC ae C40:5
PC ae C38:5
PC aa C38:5
PC ae C36:2
PC ae C42:2
PC ae C42:2
PC aa C34:2
PC aa C40:1
PC aa C40:1
PC ae C36:1
PC ae C42:3
lysoPC a C17:0
SM (OH) C16:1
PC aa C42:0
SM C16:0
C18
PC aa C40:1
PC aa C42:0
PC aa C42:1
PC aa C42:1
SM (OH) C22:2
PC ae C42:3

prote Pregnancy − exposures

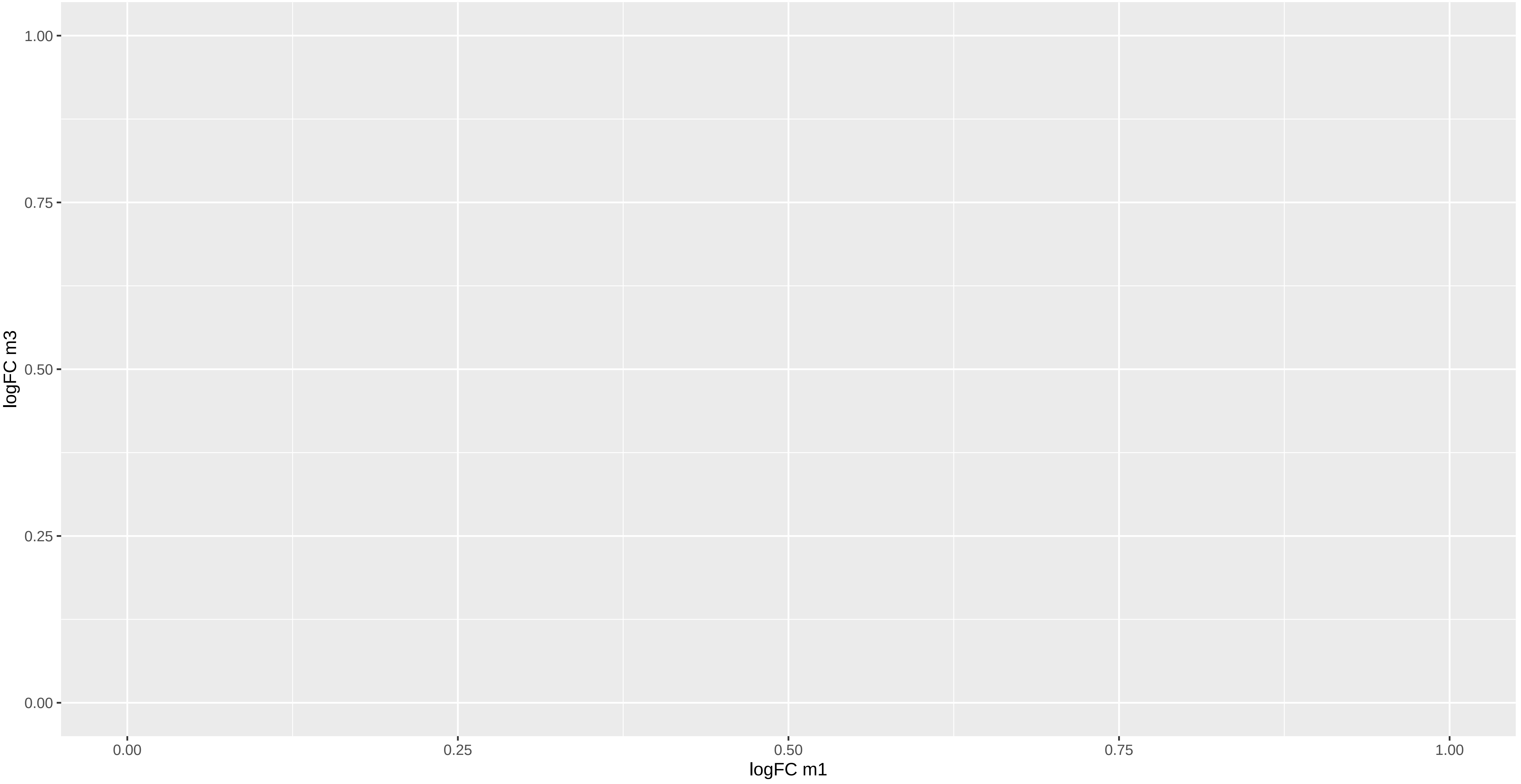

prote Pregnancy − omics markers

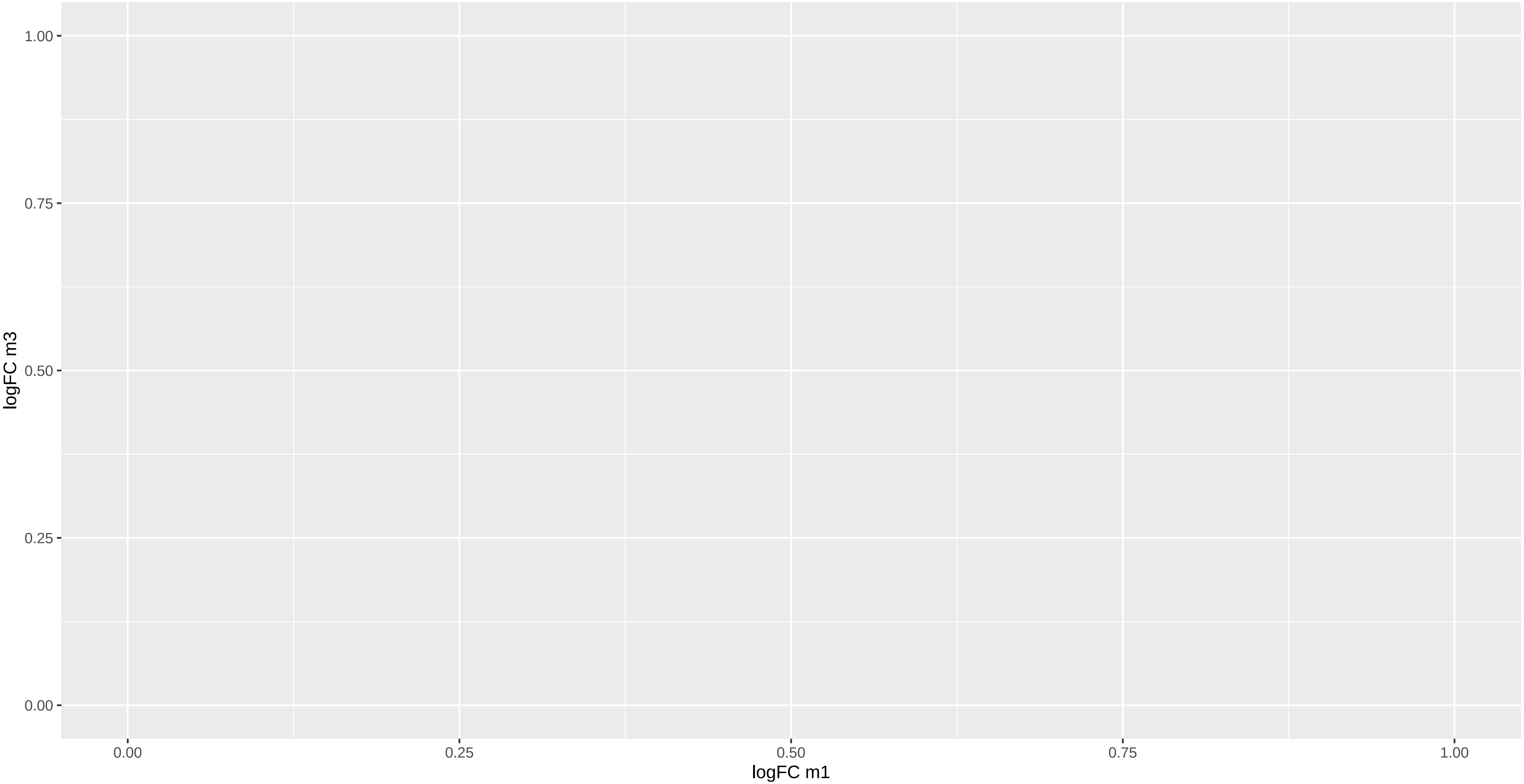

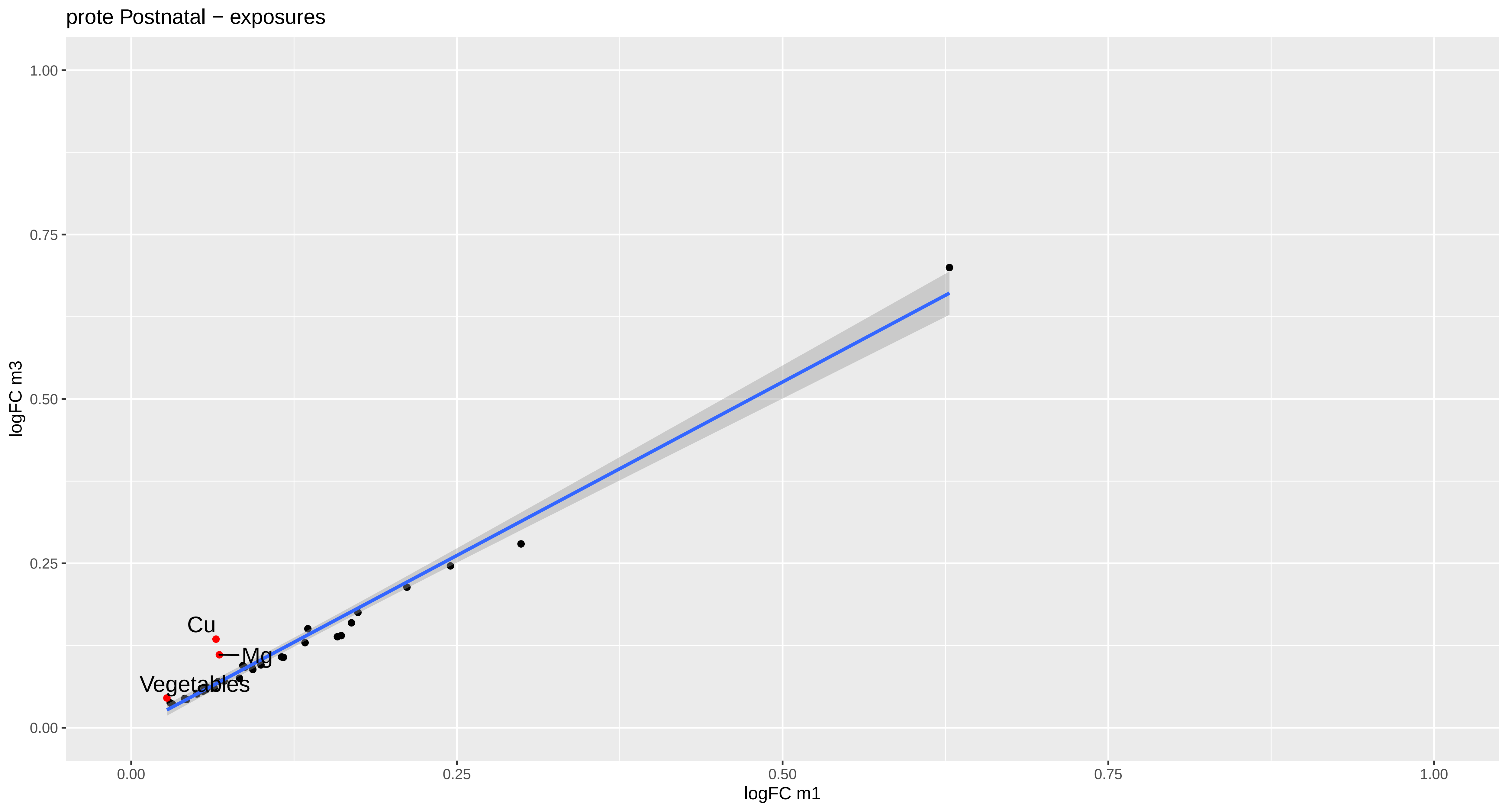

prote Postnatal − exposures
logFC m3
1.00
0.75
0.50
0.25
0.00
logFC m1
0.00
0.25
0.50
0.75
1.00
Cu
Mg
Vegetables

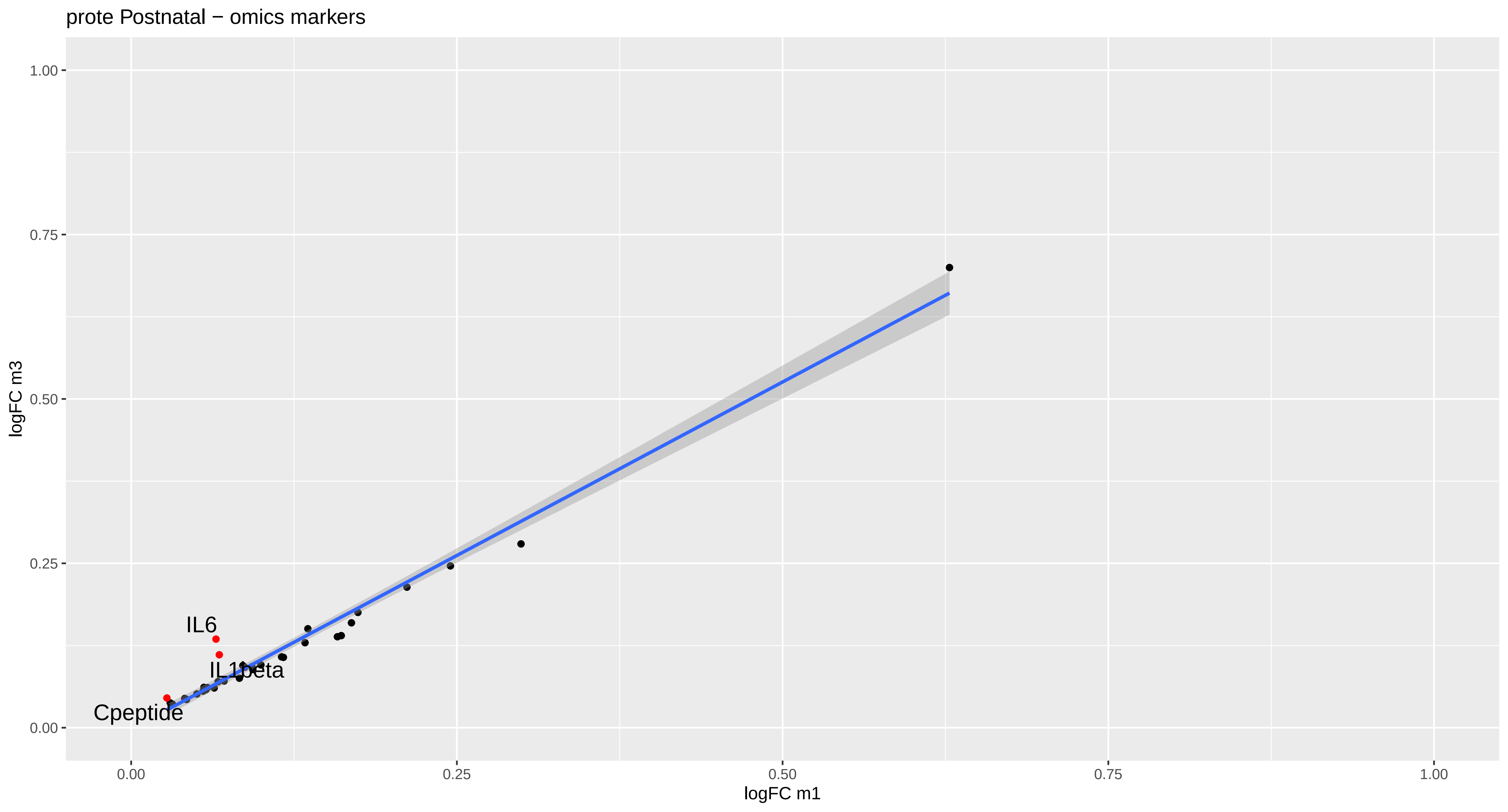

prote Postnatal − omics markers
logFC m3
logFC m1
1.00
0.75
0.50
0.25
0.00
0.00
0.25
0.50
0.75
1.00
IL6
IL1beta
Cpeptide

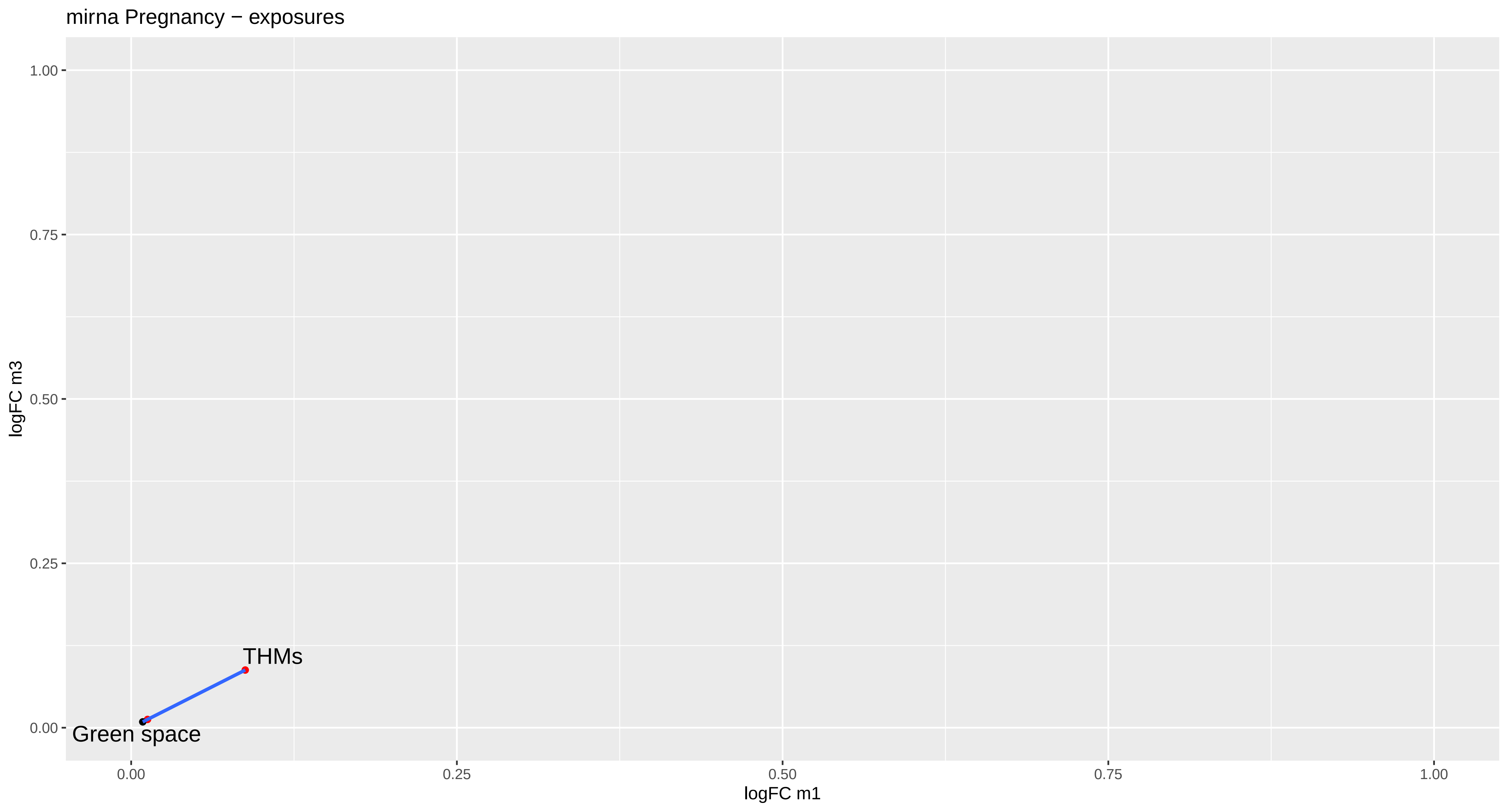
mirna Pregnancy − exposures
logFC m3
1.00
0.75
0.50
0.25
0.00
THMs
Green space
0.00
0.25
0.50
0.75
1.00
logFC m1

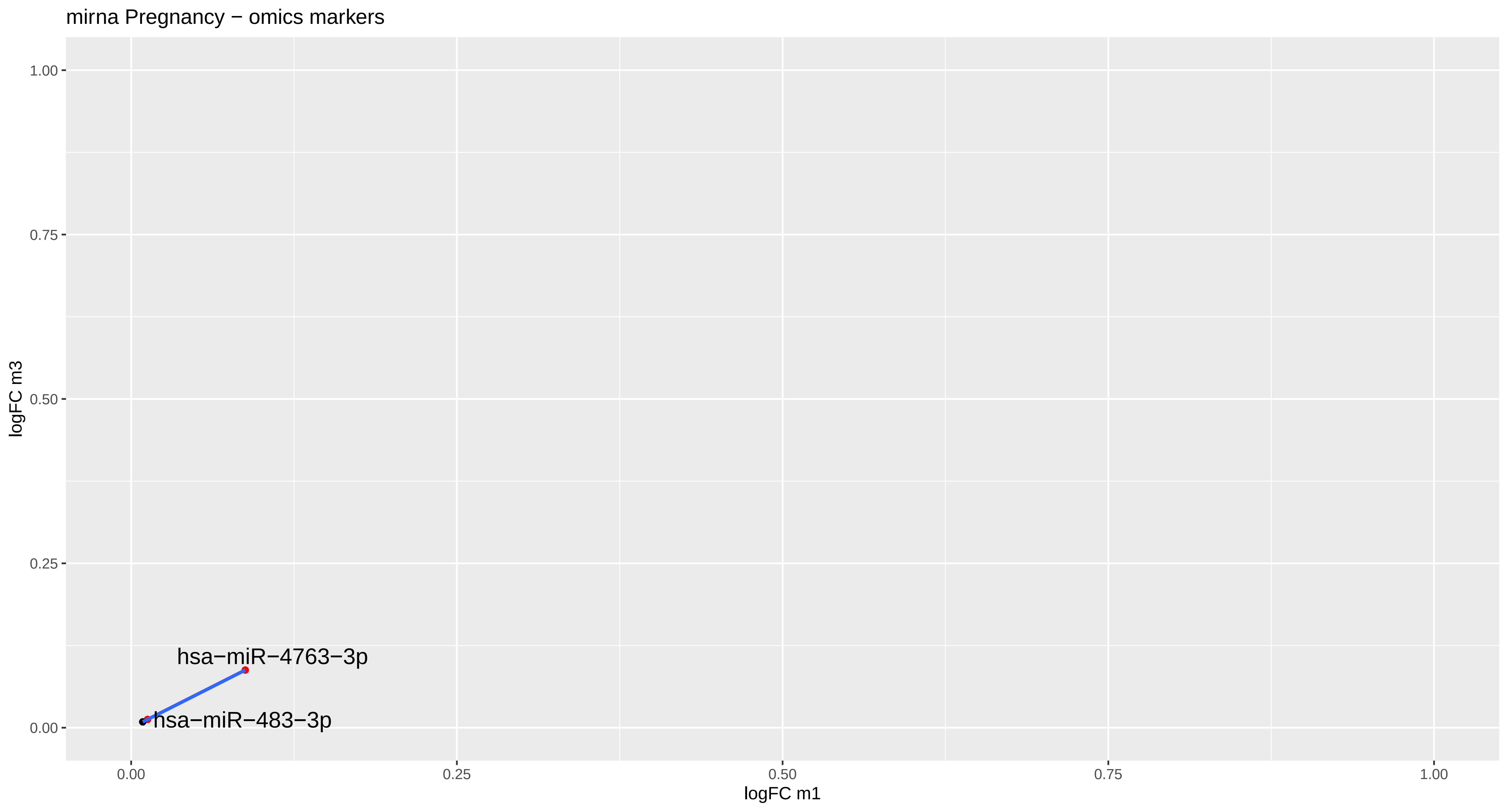
mirna Pregnancy − omics markers
logFC m3
1.00
0.75
0.50
0.25
0.00
hsa−miR−4763−3p
hsa−miR−483−3p
0.00
0.25
0.50
0.75
1.00
logFC m1

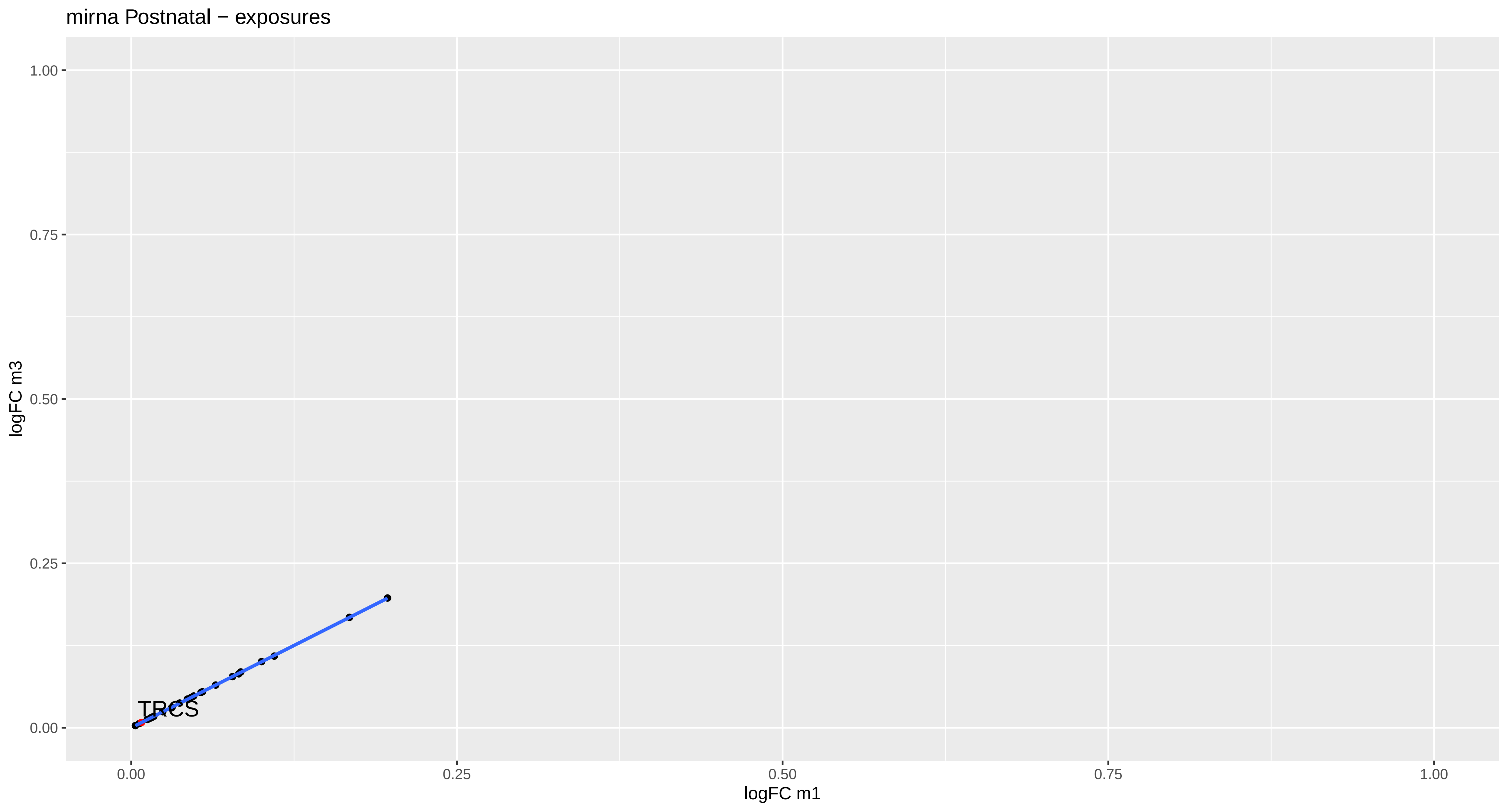

mirna Postnatal − exposures
logFC m3
1.00
0.75
0.50
0.25
0.00
0.00
0.25
0.50
0.75
1.00
logFC m1
TRCS

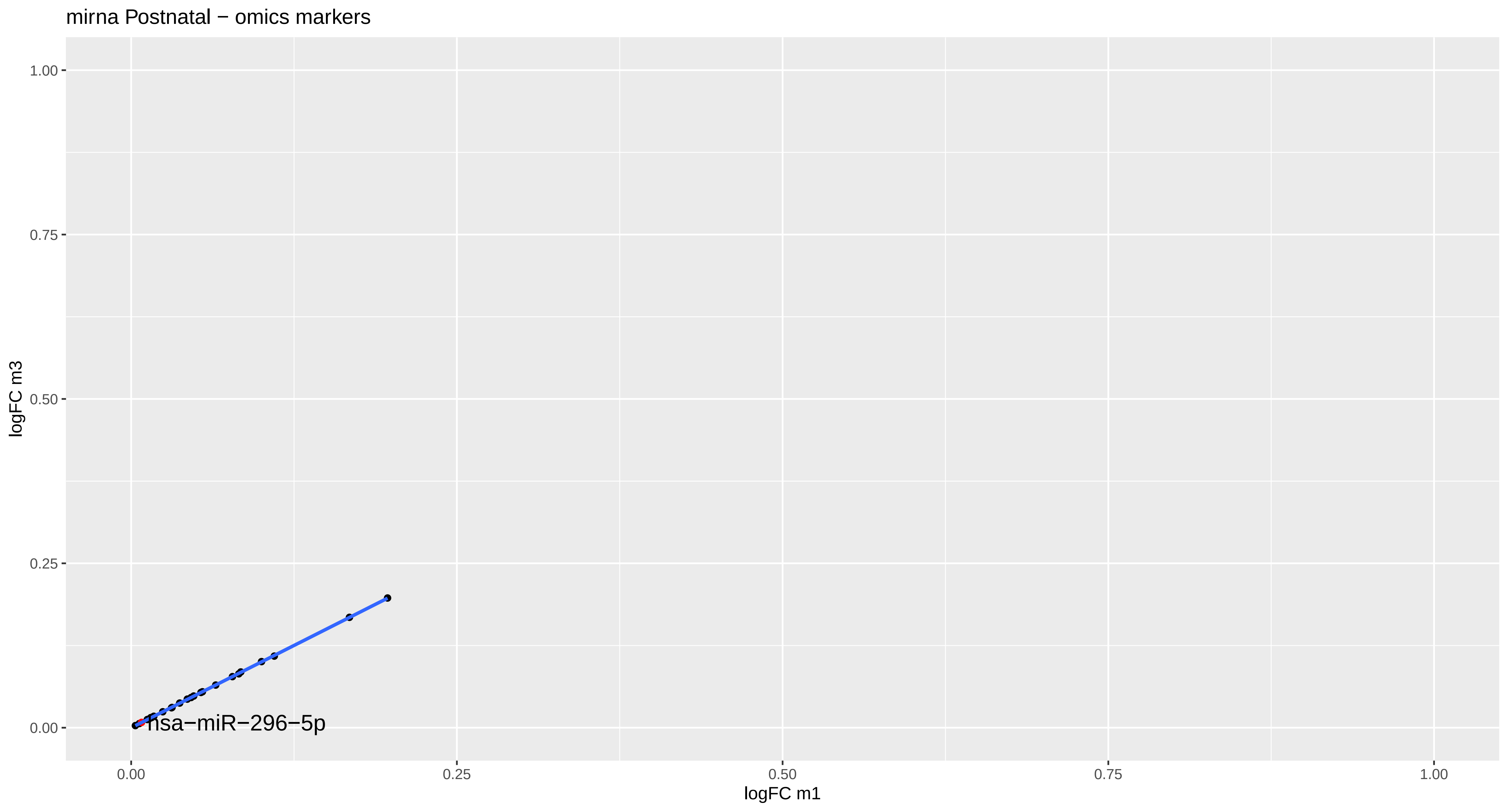
mirna Postnatal − omics markers
1.00
0.75
0.50
0.25
0.00
logFC m3
hsa−miR−296−5p
0.00
0.25
0.50
0.75
1.00
logFC m1

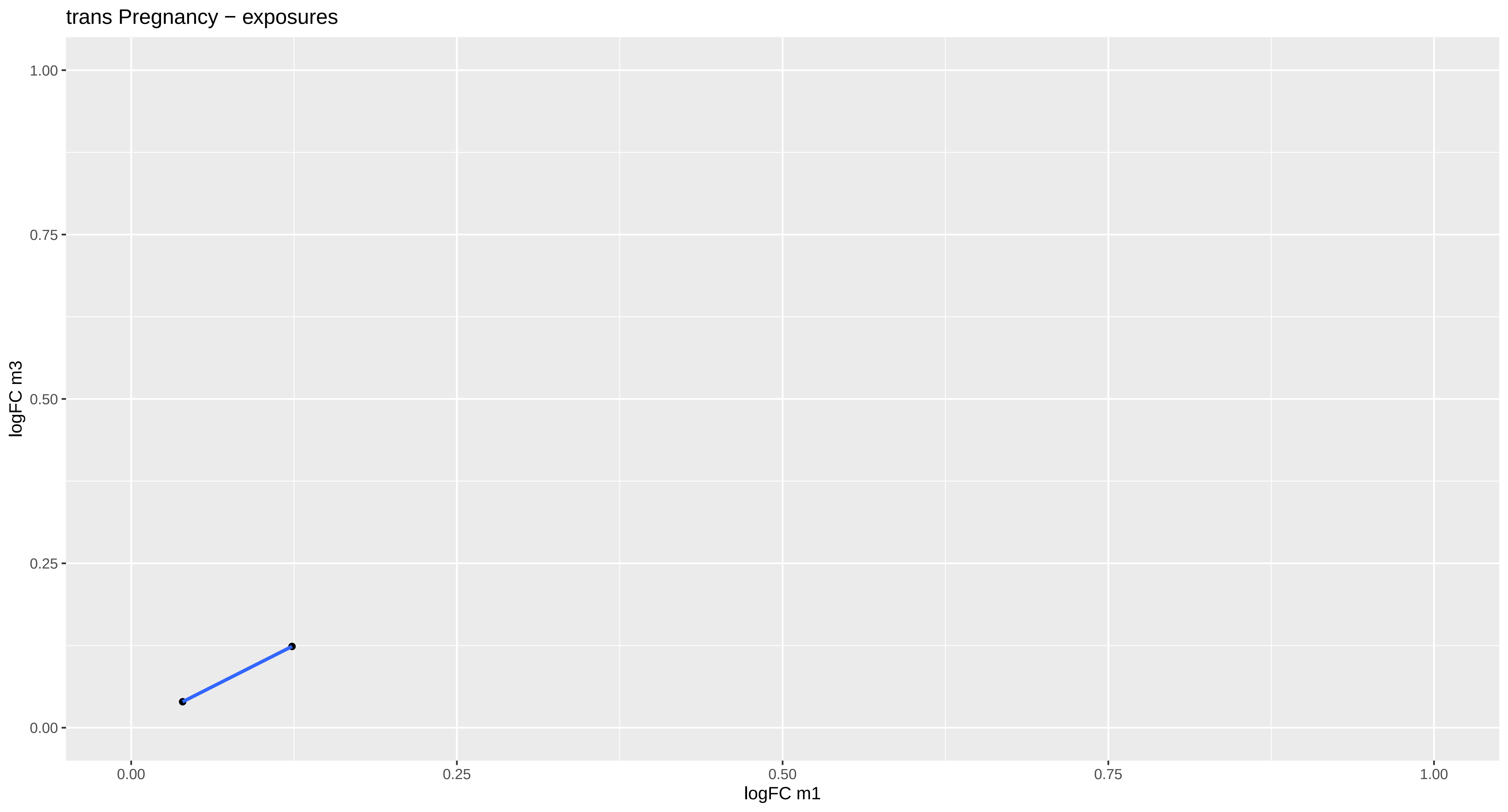
trans Pregnancy − exposures
logFC m3
1.00
0.75
0.50
0.25
0.00
0.00
0.25
0.50
0.75
1.00
logFC m1

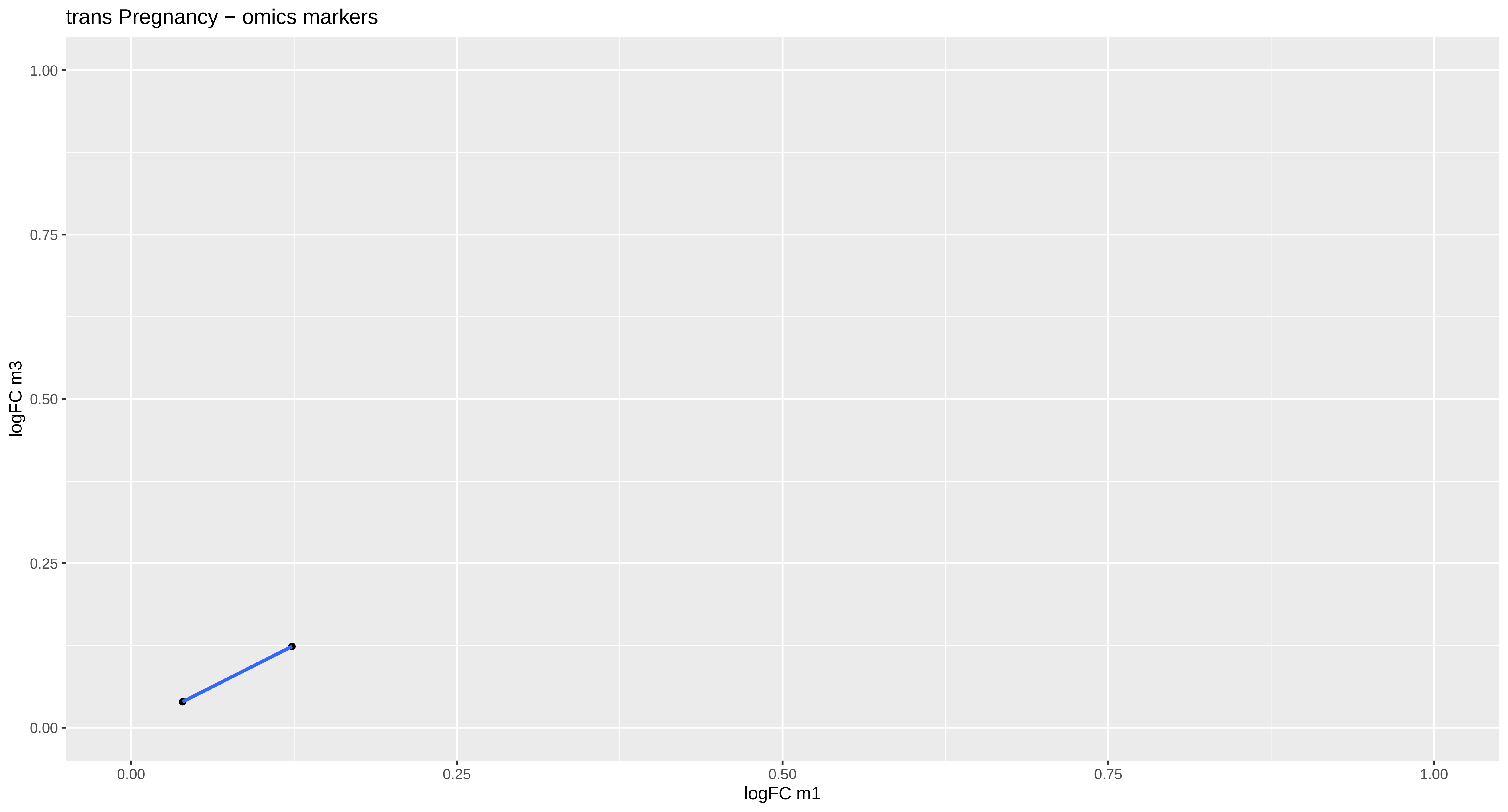
trans Pregnancy − omics markers
logFC m3
logFC m1
1.00
0.75
0.50
0.25
0.00
0.00
0.25
0.50
0.75
1.00

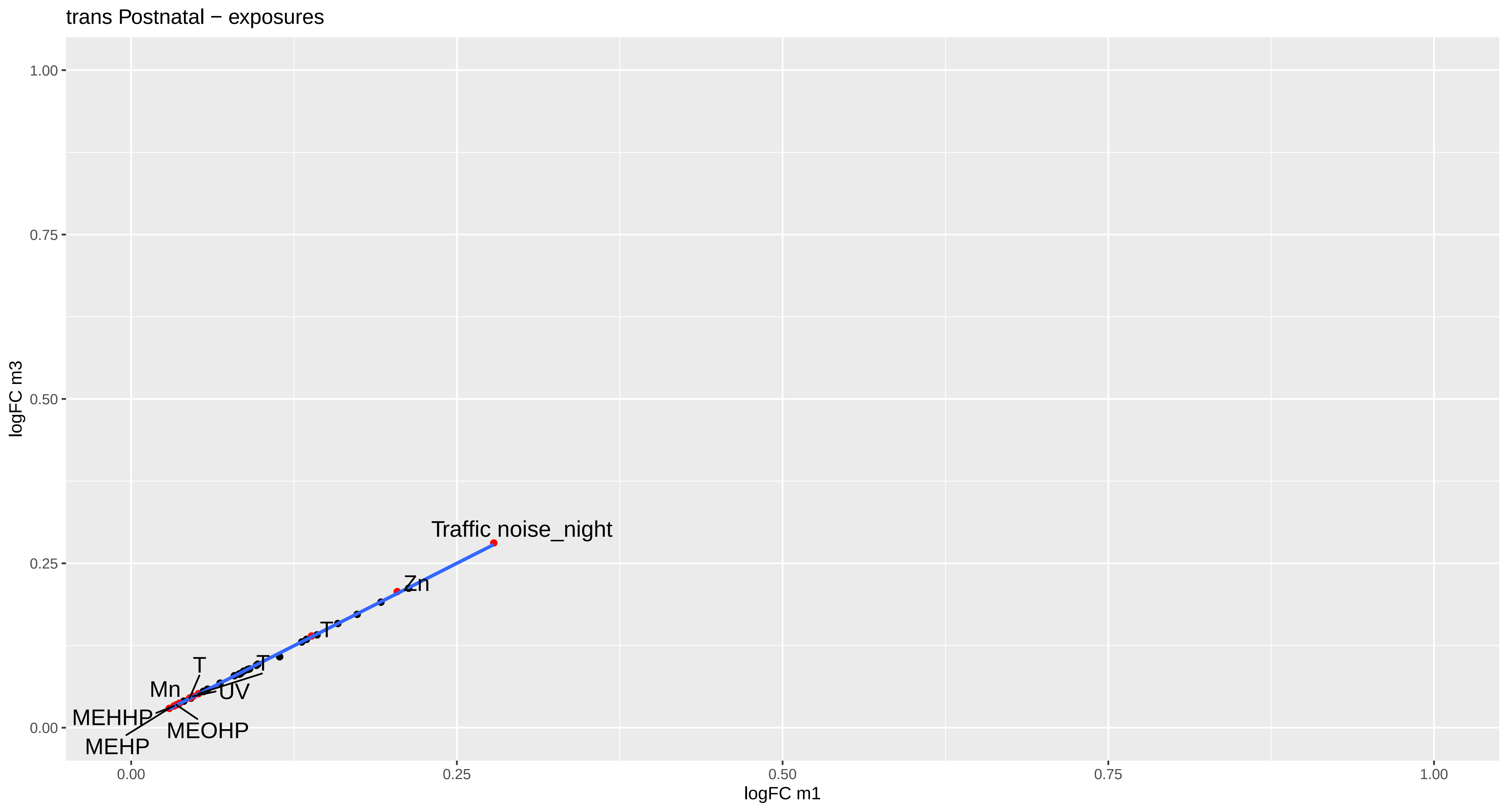

trans Postnatal − exposures
logFC m3
logFC m1
1.00
0.75
0.50
0.25
0.00
0.00
0.25
0.50
0.75
1.00
Traffic noise_night
Zn
T
T
T
Mn
UV
MEHHP
MEOHP
MEHP

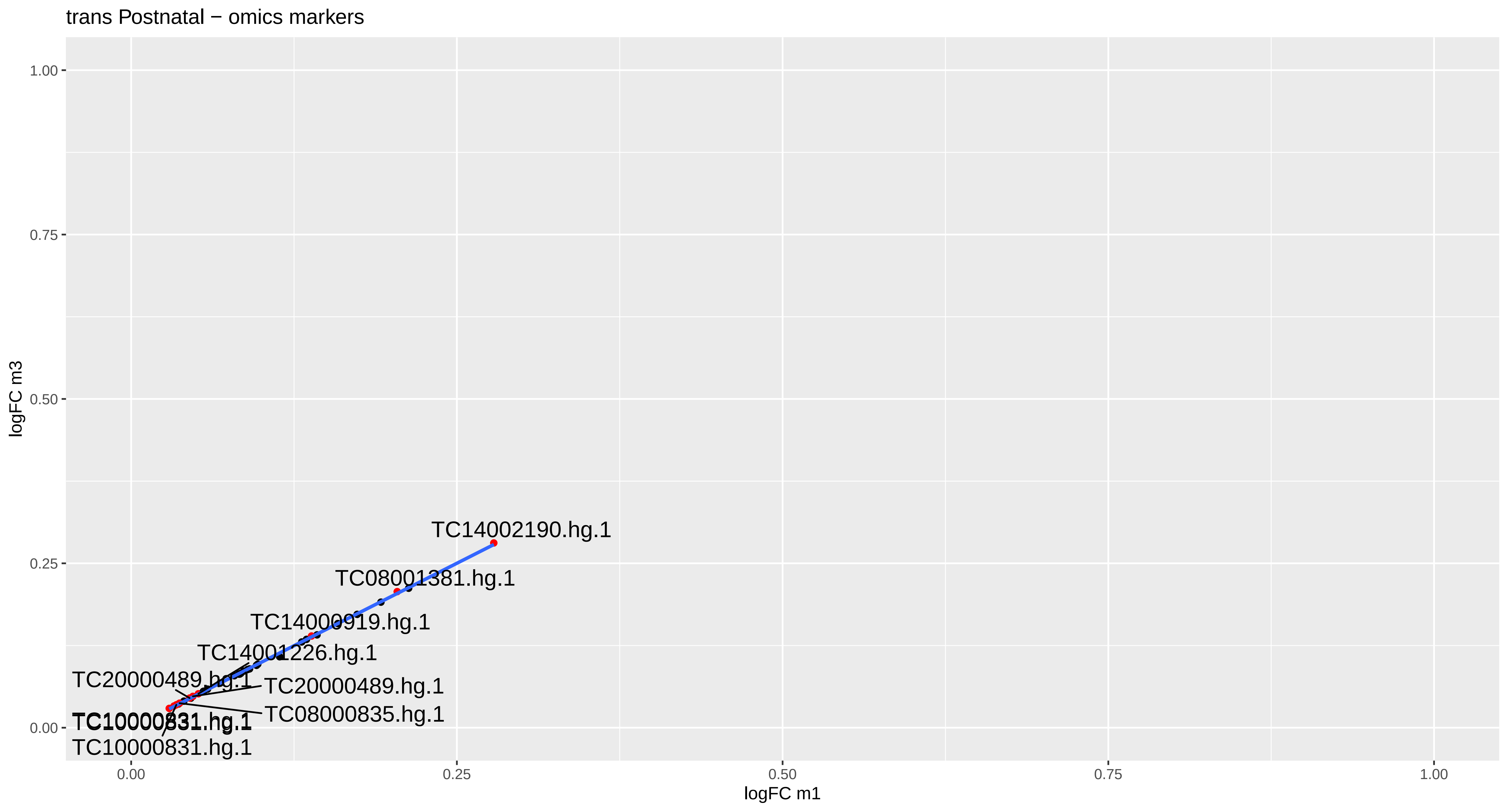

trans Postnatal − omics markers
logFC m3
logFC m1
1.00
0.75
0.50
0.25
0.00
0.00
0.25
0.50
0.75
1.00
TC14002190.hg.1
TC08001381.hg.1
TC14000919.hg.1
TC14001226.hg.1
TC20000489.hg.1
TC20000489.hg.1
TC08000835.hg.1
TC10000831.hg.1
TC10000831.hg.1

methy Pregnancy − exposures

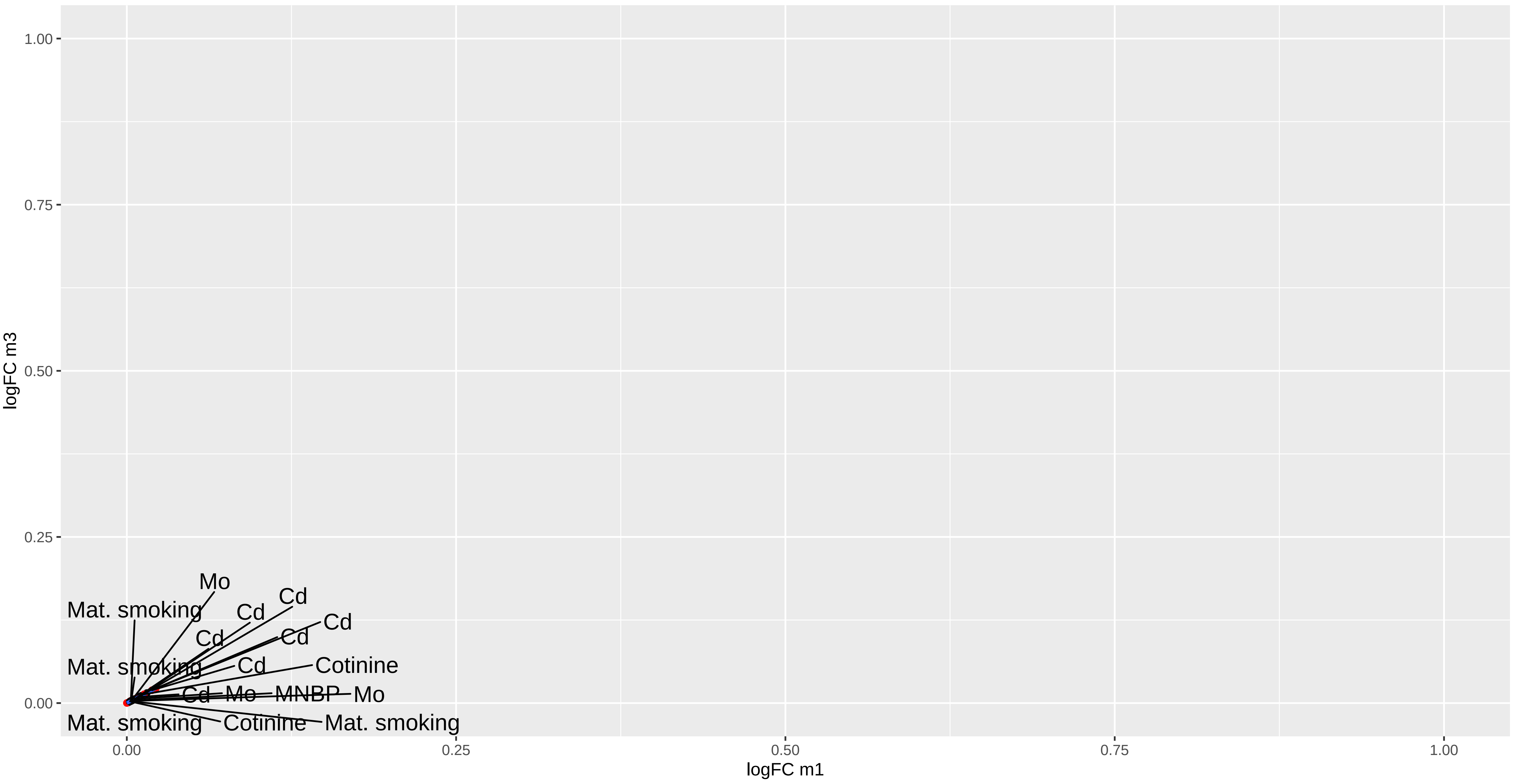

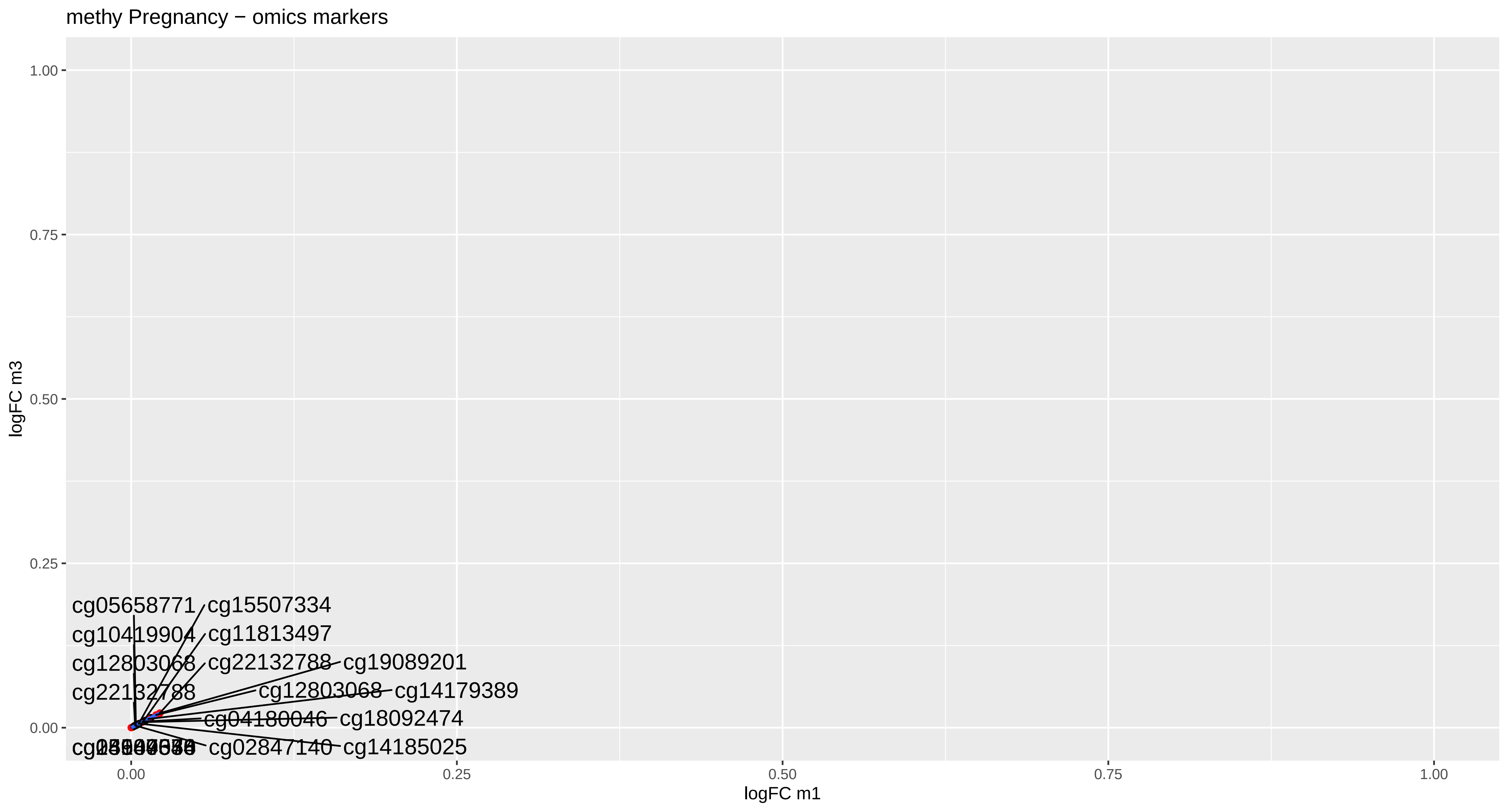
methy Pregnancy − omics markers
logFC m3
logFC m1
1.00
0.75
0.50
0.25
0.00
0.00
0.25
0.50
0.75
1.00
cg05658771
cg15507334
cg10419904
cg11813497
cg12803068
cg22132788
cg19089201
cg22132788
cg12803068
cg14179389
cg04180046
cg18092474
cg02847140
cg14185025

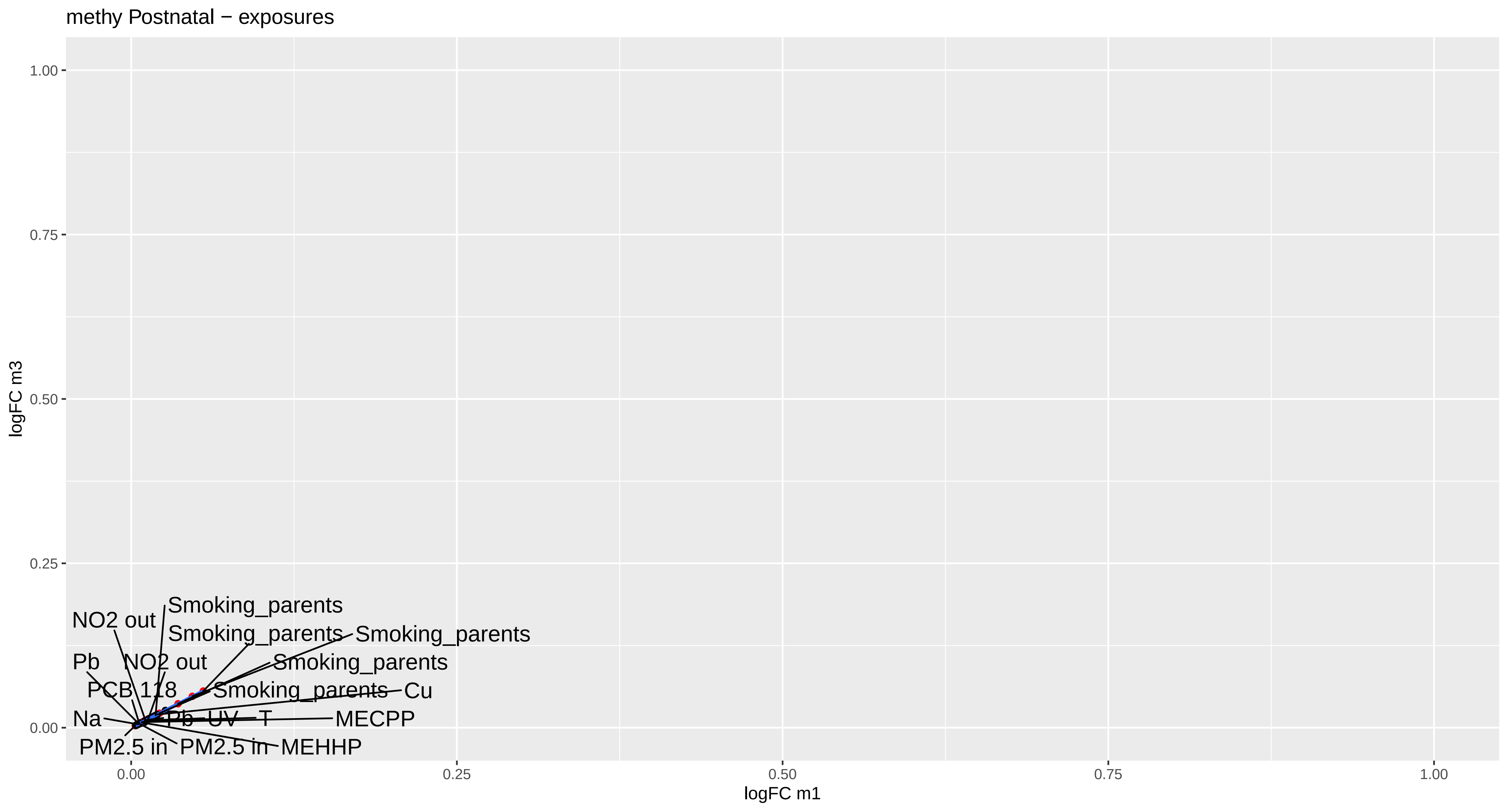

methy Postnatal − exposures
logFC m3
logFC m1
1.00
0.75
0.50
0.25
0.00
0.00
0.25
0.50
0.75
1.00
Smoking_parents
NO2 out
Smoking_parents
Smoking_parents
Pb
NO2 out
Smoking_parents
PCB 118
Smoking_parents
Cu
Na
Pb
UV
T
MECPP
PM2.5 in
PM2.5 in
MEHHP

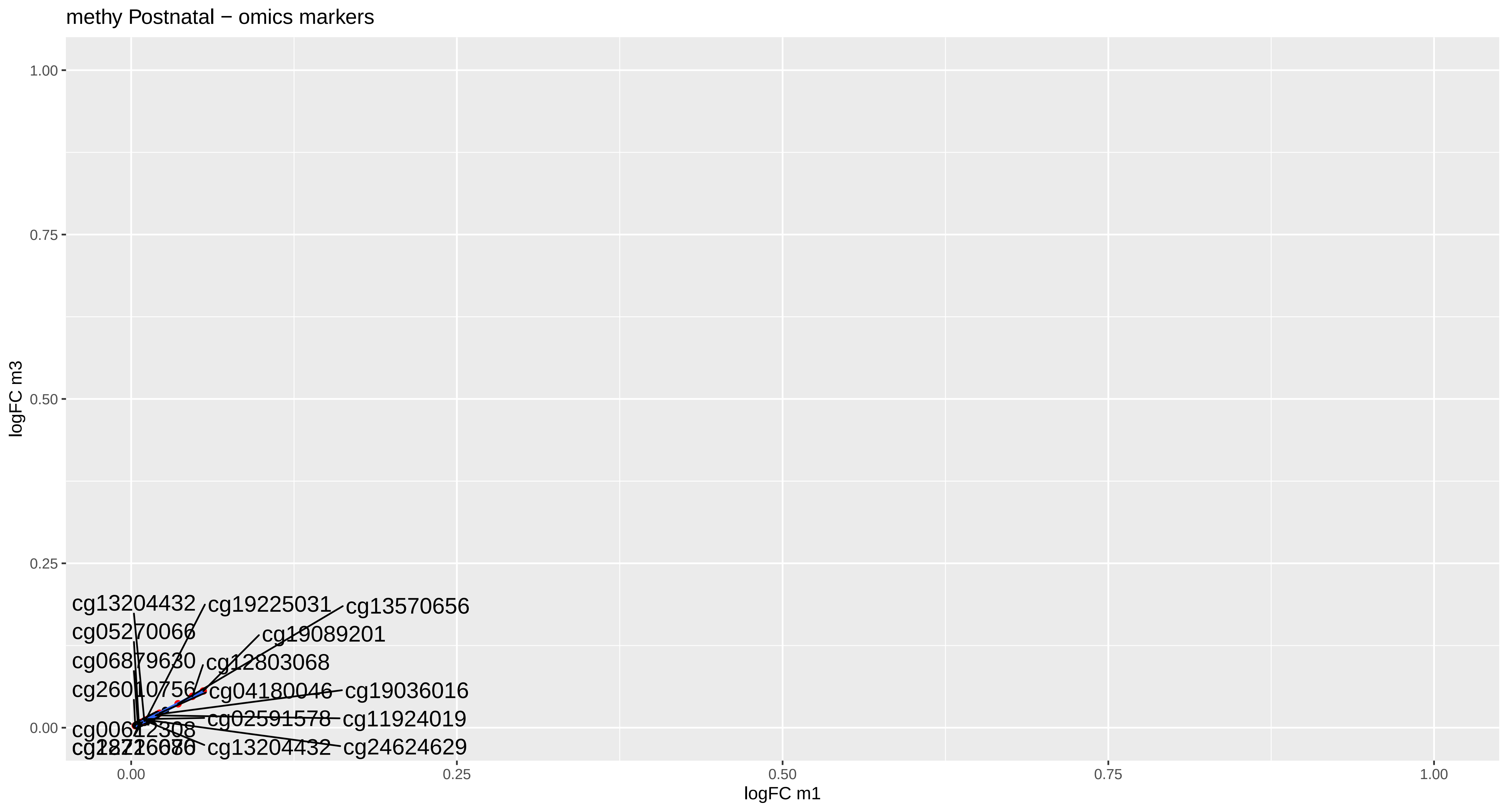
methy Postnatal − omics markers
logFC m3
logFC m1
1.00
0.75
0.50
0.25
0.00
0.00
0.25
0.50
0.75
1.00
cg13204432
cg19225031
cg13570656
cg05270066
cg19089201
cg06879630
cg12803068
cg26010756
cg04180046
cg19036016
cg02591578
cg11924019
cg13204432
cg24624629
