## Supplementary material for "Multi-omics signatures of the human early life exposome": Figure S3

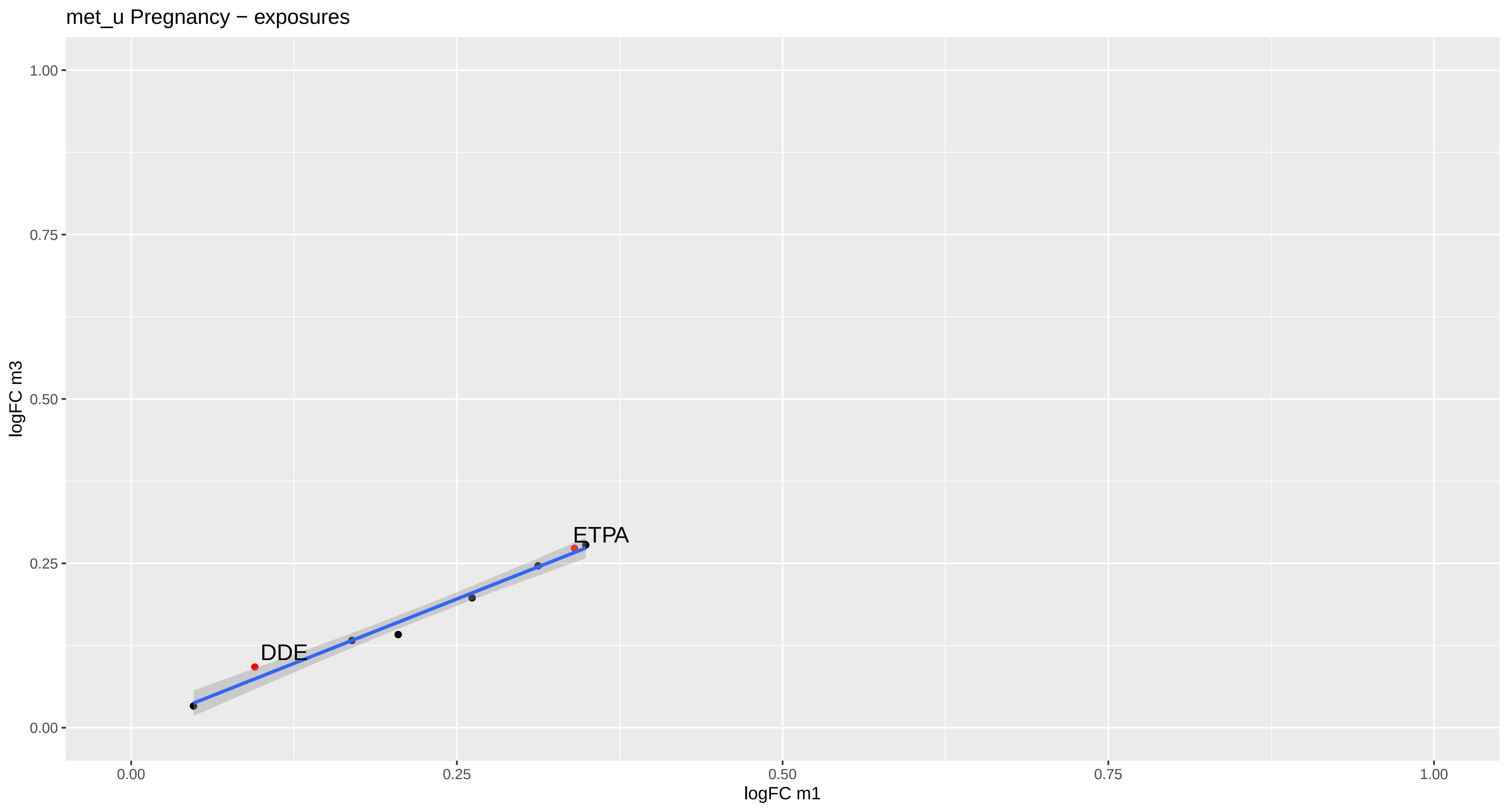
met_u Pregnancy − exposures
logFC m3
logFC m1
1.00
0.75
0.50
0.25
0.00
0.00
0.25
0.50
0.75
1.00
DDE
ETPA

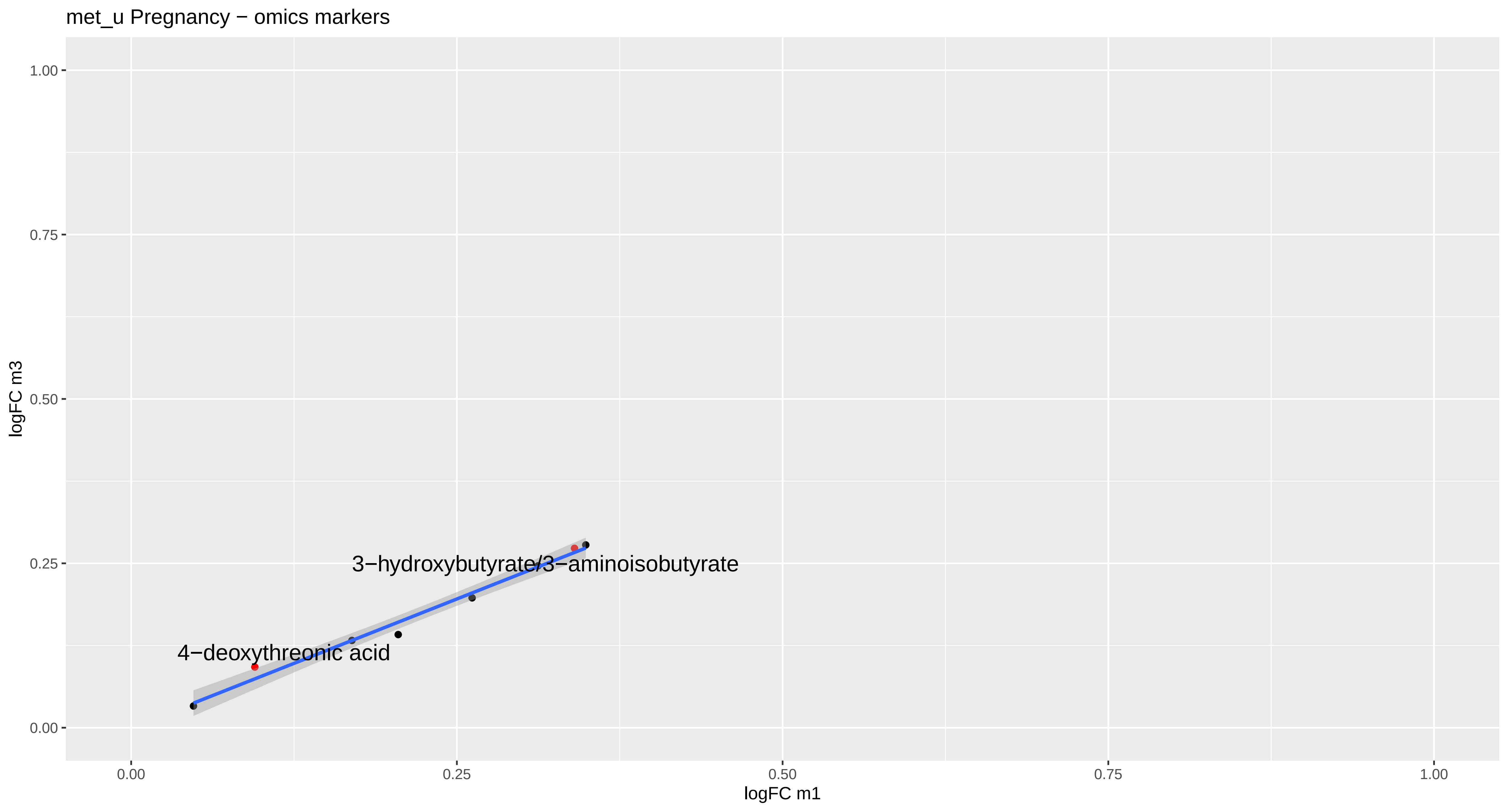

met_u Pregnancy − omics markers
1.00
0.75
0.50
0.25
0.00
logFC m3
3−hydroxybutyrate/3−aminoisobutyrate
4−deoxythreonic acid
0.00
0.25
0.50
0.75
1.00
logFC m1

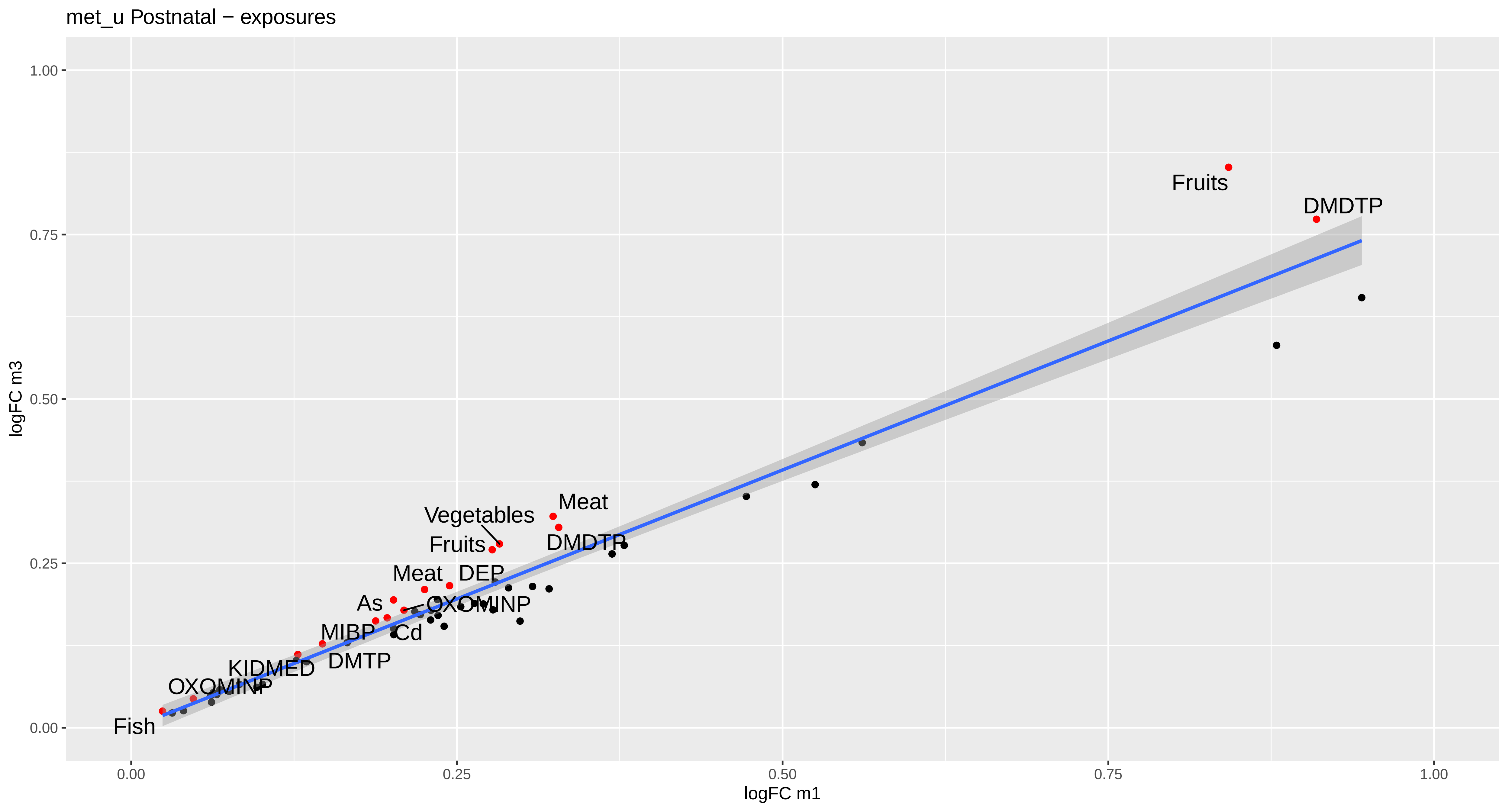
met_u Postnatal − exposures
logFC m3
logFC m1
1.00
0.75
0.50
0.25
0.00
0.00
0.25
0.50
0.75
1.00
Fruits
DMDTP
Meat
Vegetables
Fruits
DMDTP
Meat
DEP
As
OXOMINP
MIBP
Cd
DMTP
KIDMED
OXOMINP
Fish

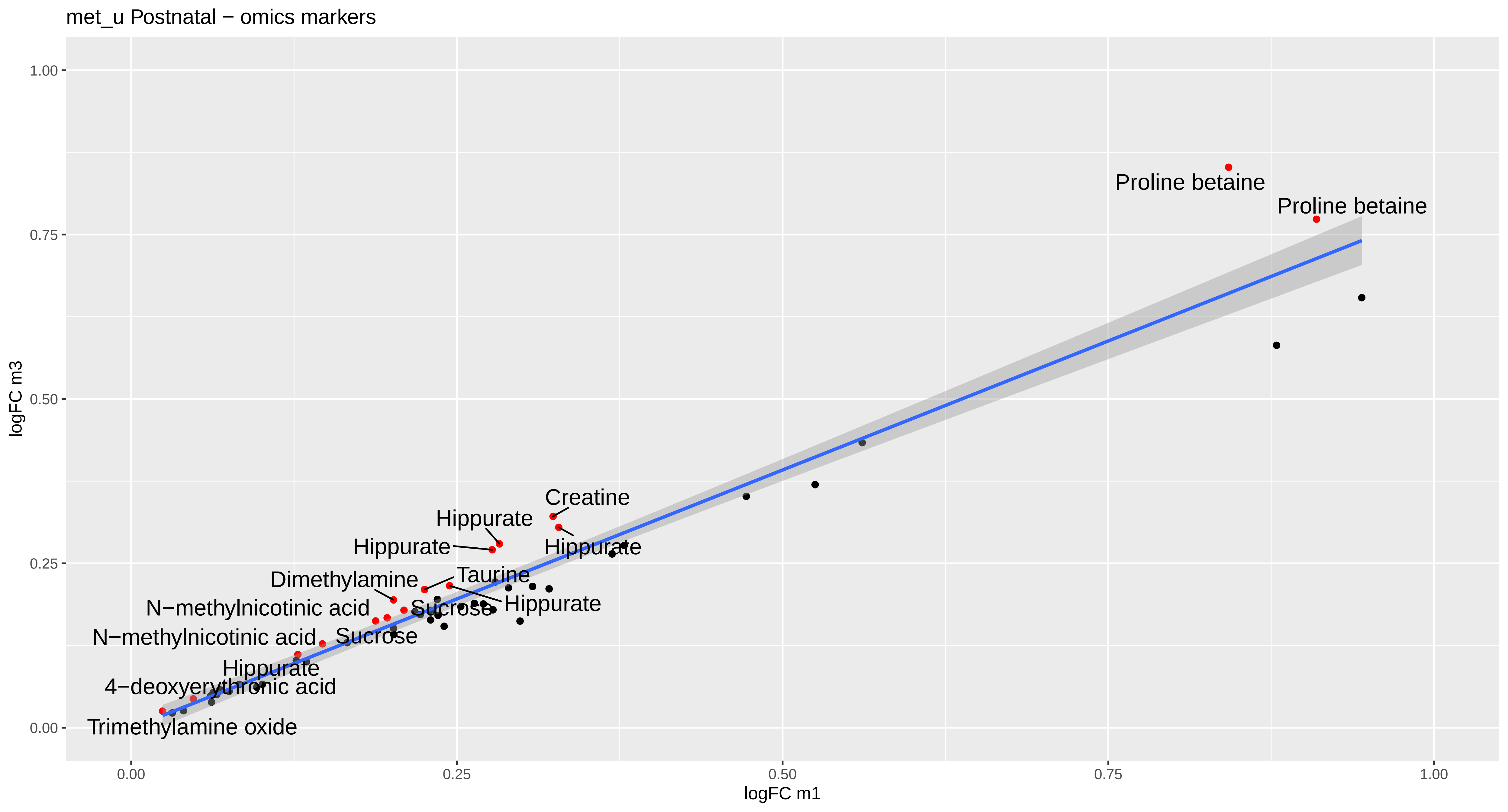
met_u Postnatal − omics markers
logFC m3
logFC m1
0.00
0.25
0.50
0.75
1.00
Proline betaine
Proline betaine
Creatine
Hippurate
Hippurate
Hippurate
Dimethylamine
Taurine
Hippurate
N−methylnicotinic acid
Sucrose
N−methylnicotinic acid
Sucrose
Hippurate
4−deoxyerythronic acid
Trimethylamine oxide

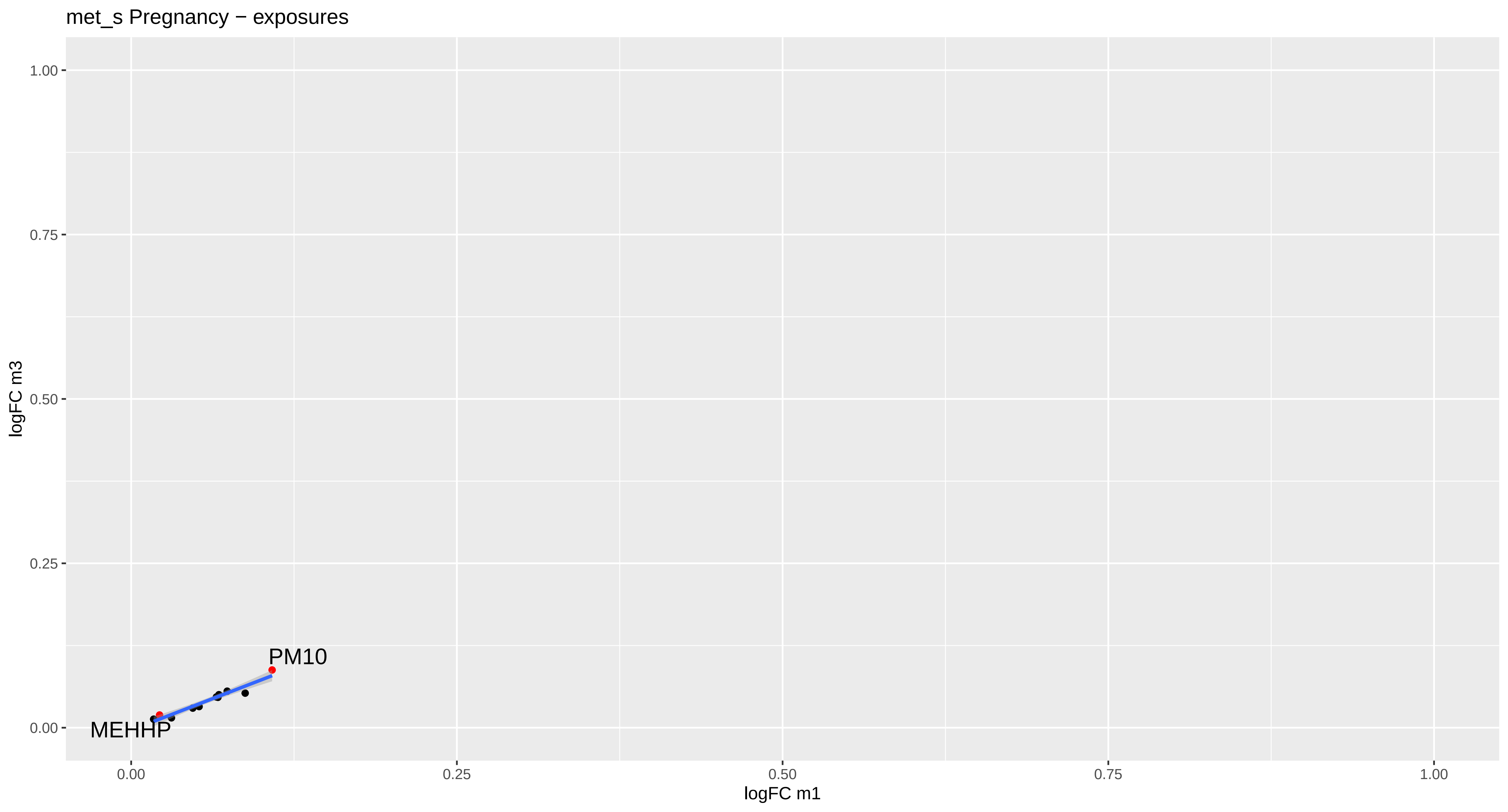
met_s Pregnancy − exposures
1.00
0.75
0.50
0.25
0.00
logFC m3
PM10
MEHHP
0.00
0.25
0.50
0.75
1.00
logFC m1

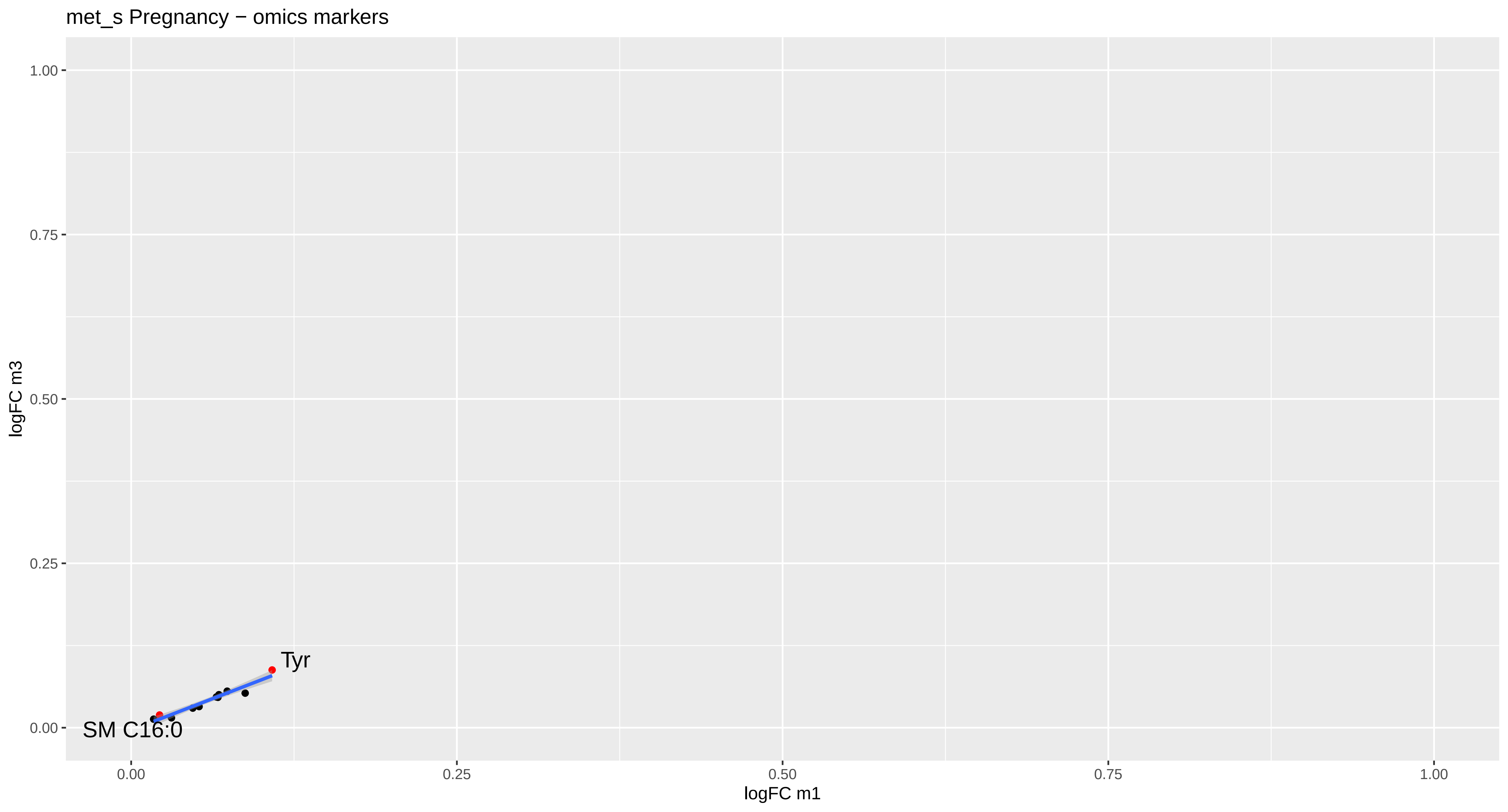

met_s Pregnancy − omics markers
logFC m3
1.00
0.75
0.50
0.25
0.00
Tyr
SM C16:0
0.00
0.25
0.50
0.75
1.00
logFC m1

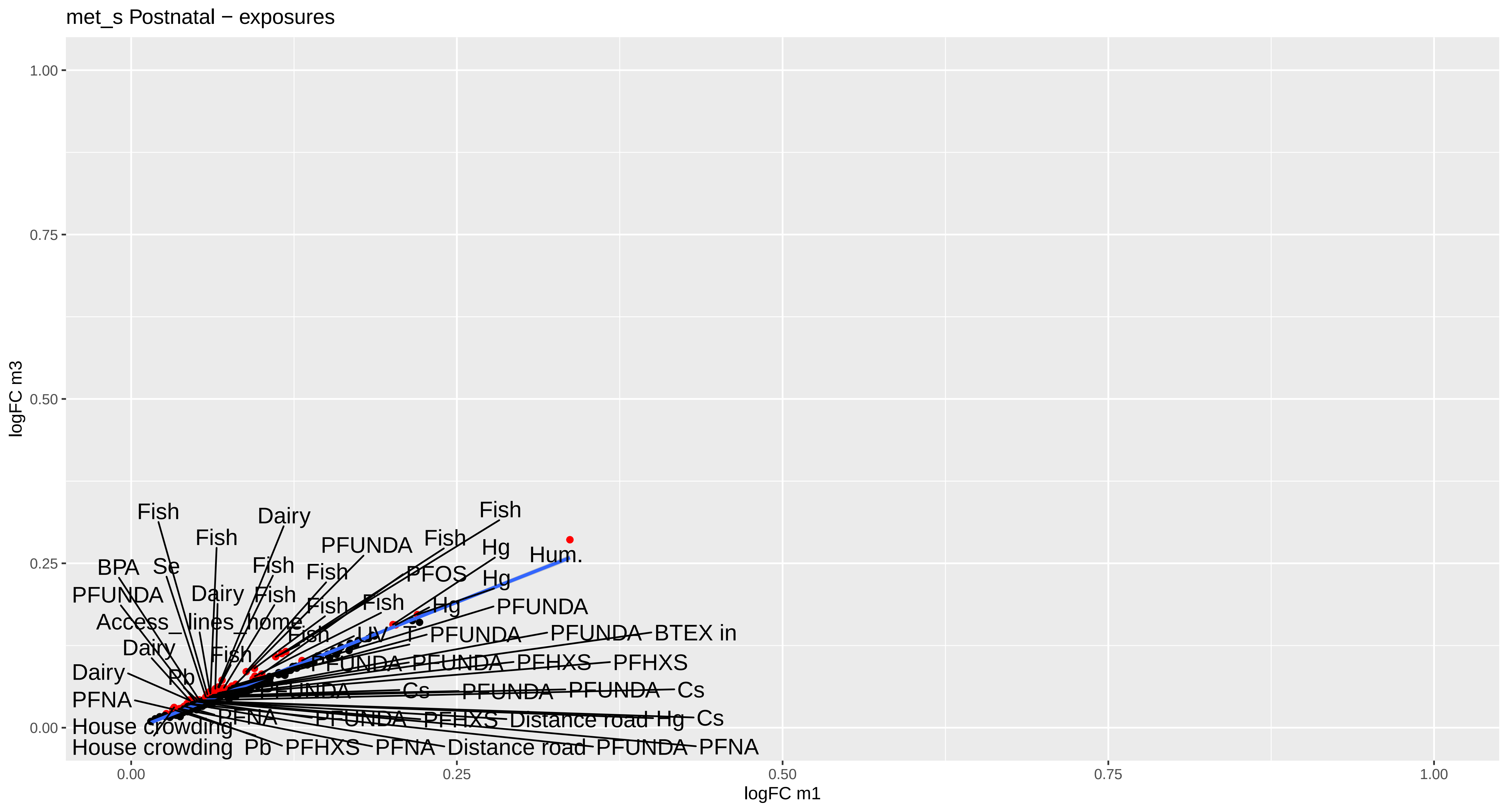
met_s Postnatal − exposures
logFC m3
logFC m1
1.00
0.75
0.50
0.25
0.00
0.00
0.25
0.50
0.75
1.00
Fish
Dairy
Fish
PFUNDA
Fish
Fish
Hg
Hum.
BPA
Se
Fish
Fish
PFOS
Hg
PFUNDA
Dairy
Fish
Fish
Fish
Hg
PFUNDA
Access_lines_home
Fish
UV
T
PFUNDA
PFUNDA
BTEX in
Dairy
Fish
PFUNDA
PFUNDA
PFHXS
PFHXS
Dairy
Pb
PFUNDA
Cs
PFUNDA
PFUNDA
Cs
PFNA
PFNA
PFUNDA
PFHXS
Distance road
Hg
Cs
House crowding
House crowding
Pb
PFHXS
PFNA
Distance road
PFUNDA
PFNA

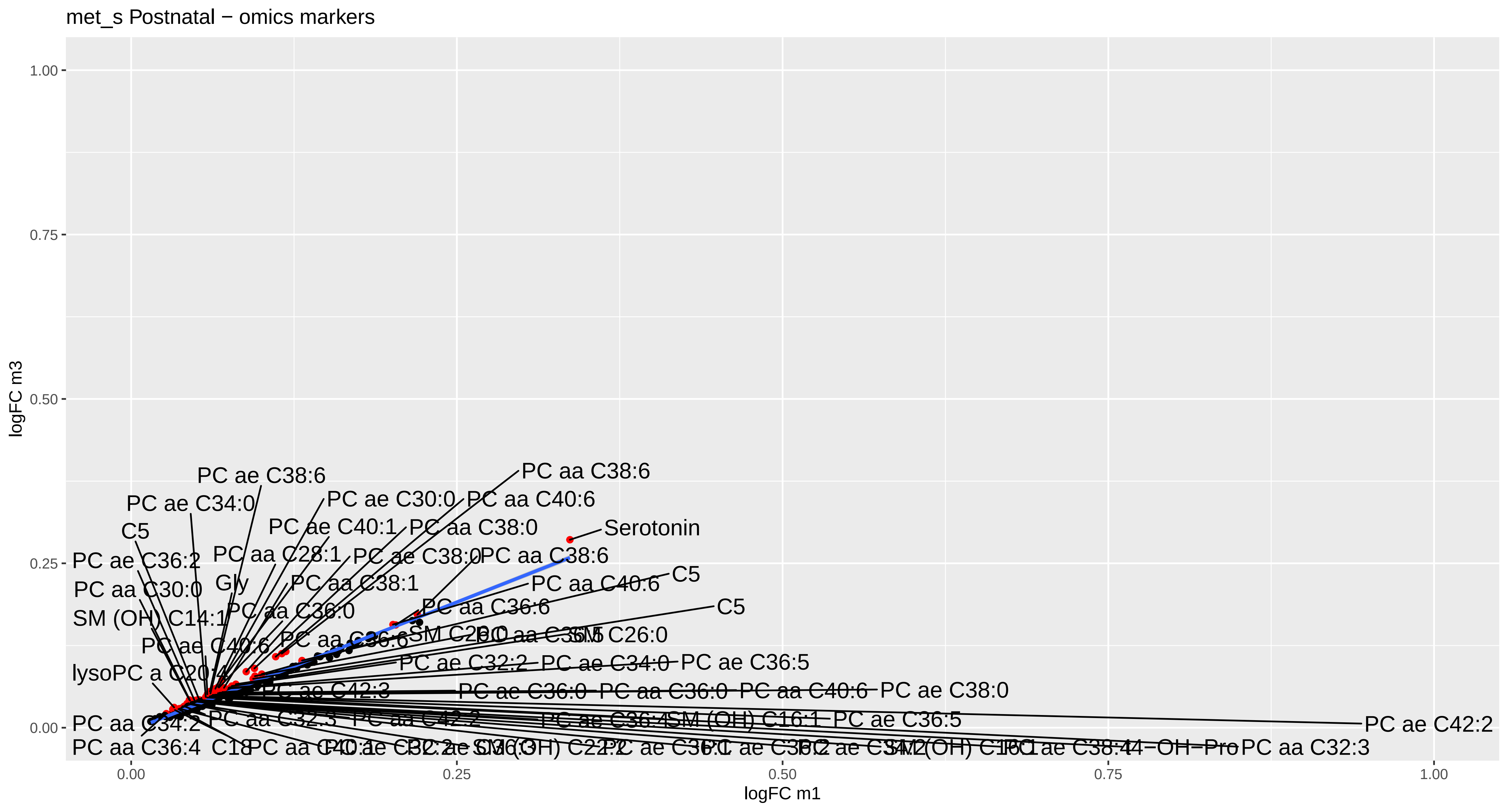
met_s Postnatal − omics markers
logFC m3
logFC m1
0.00
0.25
0.50
0.75
1.00
PC aa C38:6
PC ae C38:6
PC ae C34:0
PC ae C30:0
PC aa C40:6
C5
PC ae C40:1
PC aa C38:0
Serotonin
PC ae C36:2
PC aa C28:1
PC ae C38:0
PC aa C30:0
Gly
PC aa C38:1
SM (OH) C14:1
PC aa C36:0
PC aa C36:6
PC ae C40:6
SM C26:0
lysoPC a C20:4
PC ae C32:2
PC ae C34:0
PC ae C36:5
PC ae C42:3
PC ae C36:0
PC aa C36:0
PC aa C40:6
PC ae C38:0
PC aa C34:2
PC aa C32:3
PC aa C42:2
SM (OH) C16:1
PC ae C42:2
PC aa C36:4
C18
PC aa C32:3

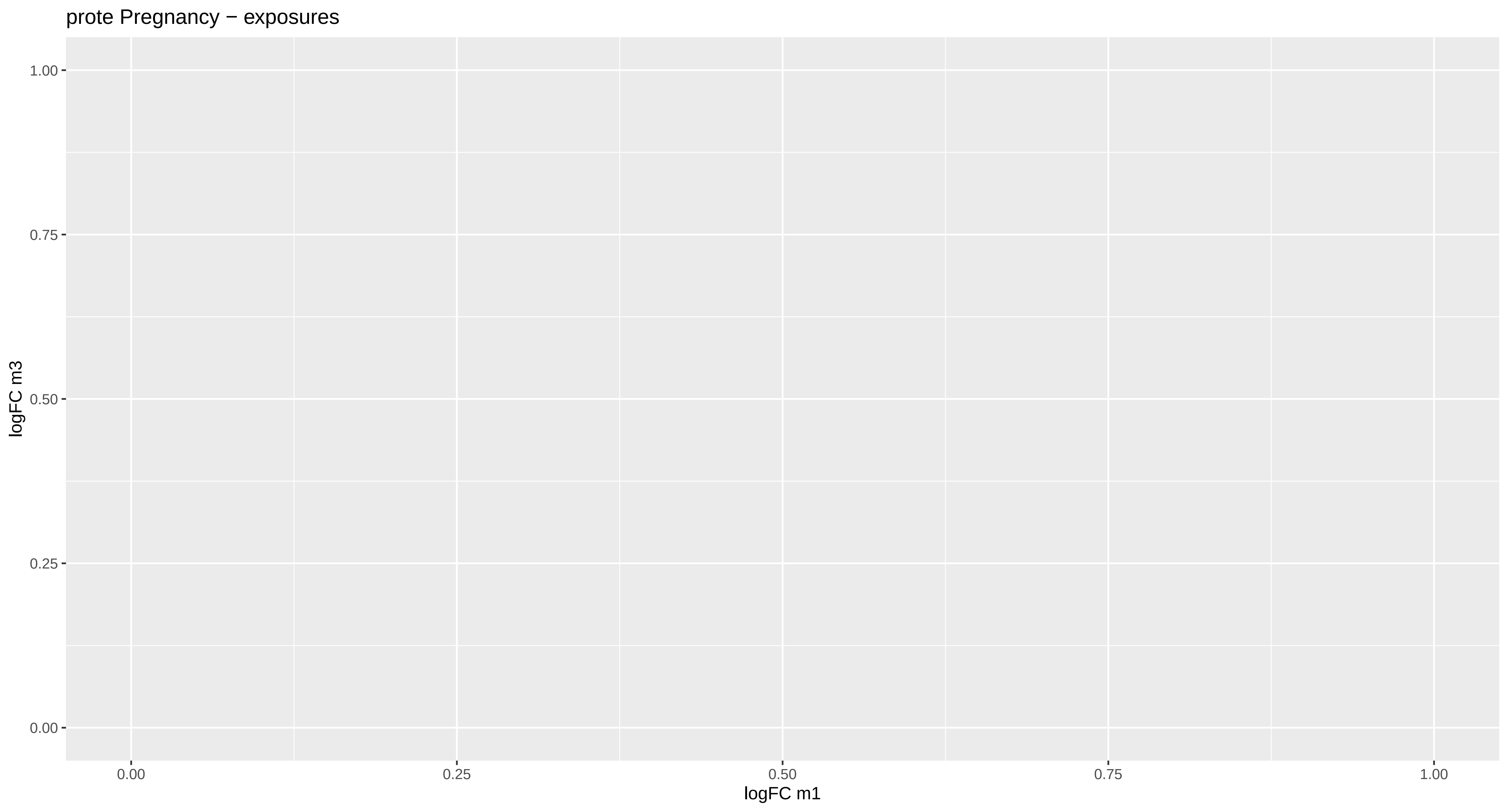
prote Pregnancy − exposures
logFC m3
1.00
0.75
0.50
0.25
0.00
0.00
0.25
0.50
0.75
1.00
logFC m1

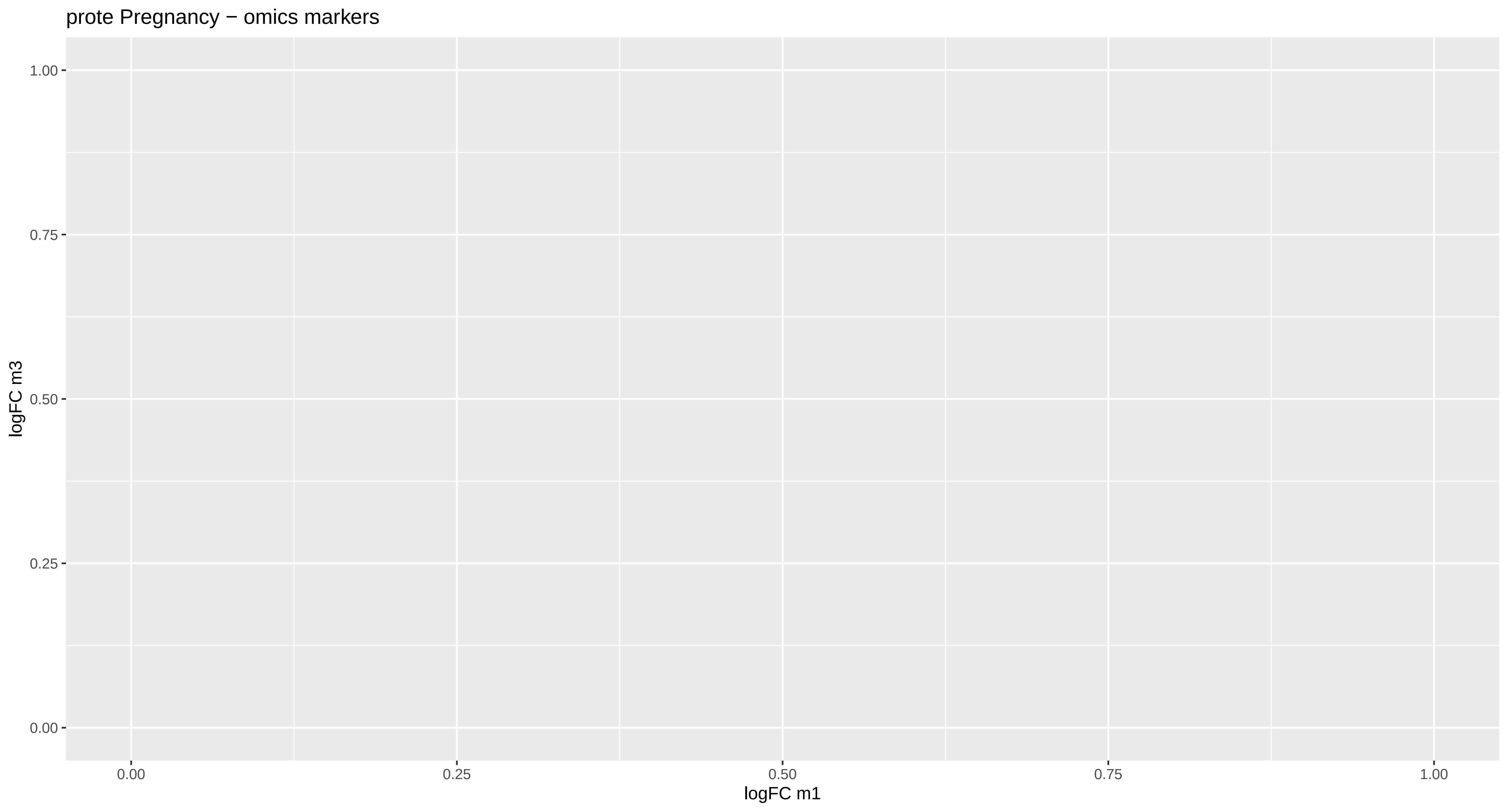
prote Pregnancy − omics markers
logFC m3
1.00
0.75
0.50
0.25
0.00
0.00
0.25
0.50
0.75
1.00
logFC m1

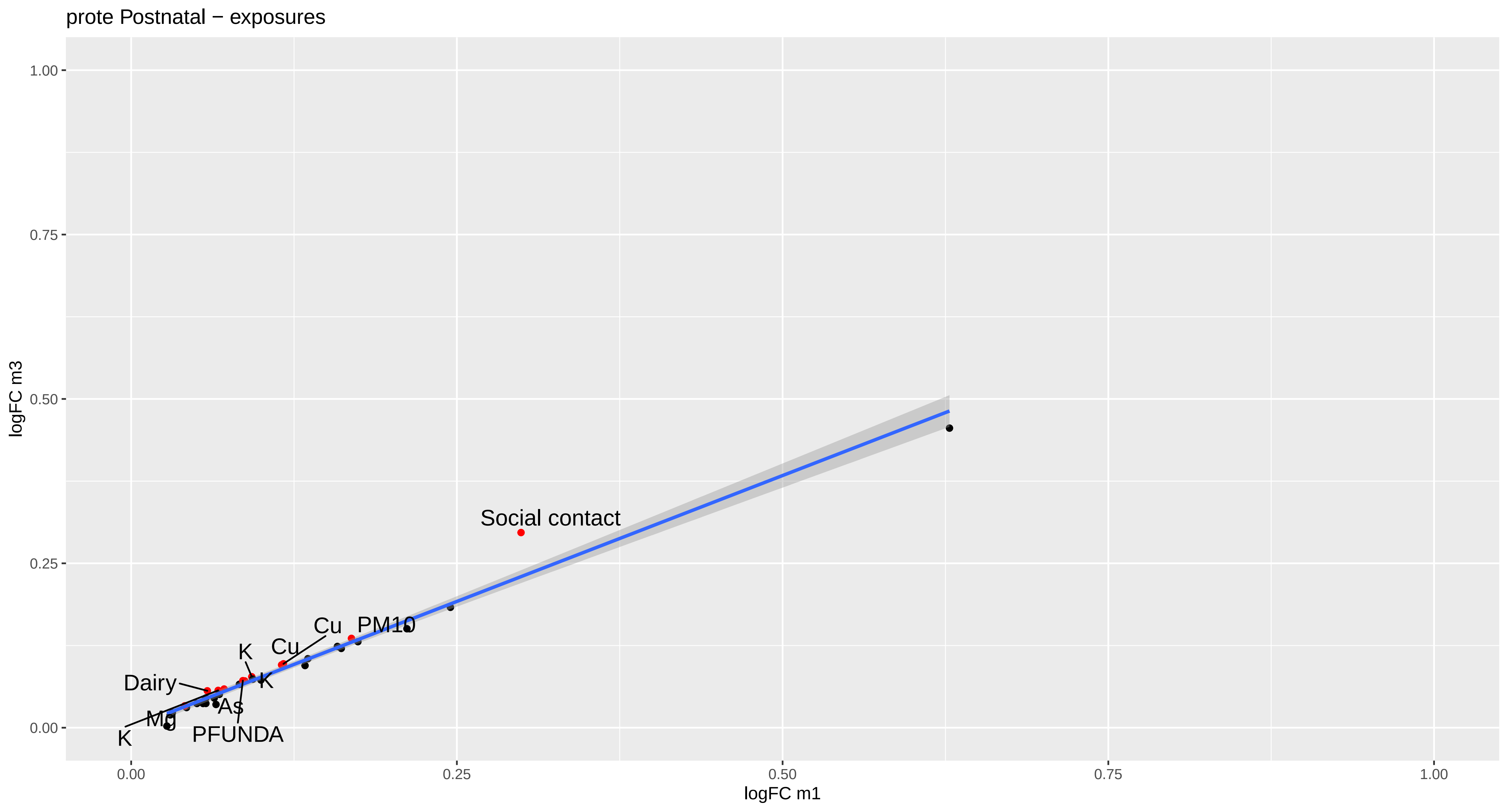
prote Postnatal − exposures
logFC m3
logFC m1
1.00
0.75
0.50
0.25
0.00
0.00
0.25
0.50
0.75
1.00
Social contact
PM10
Cu
Cu
K
K
Dairy
As
Mg
K
PFUNDA

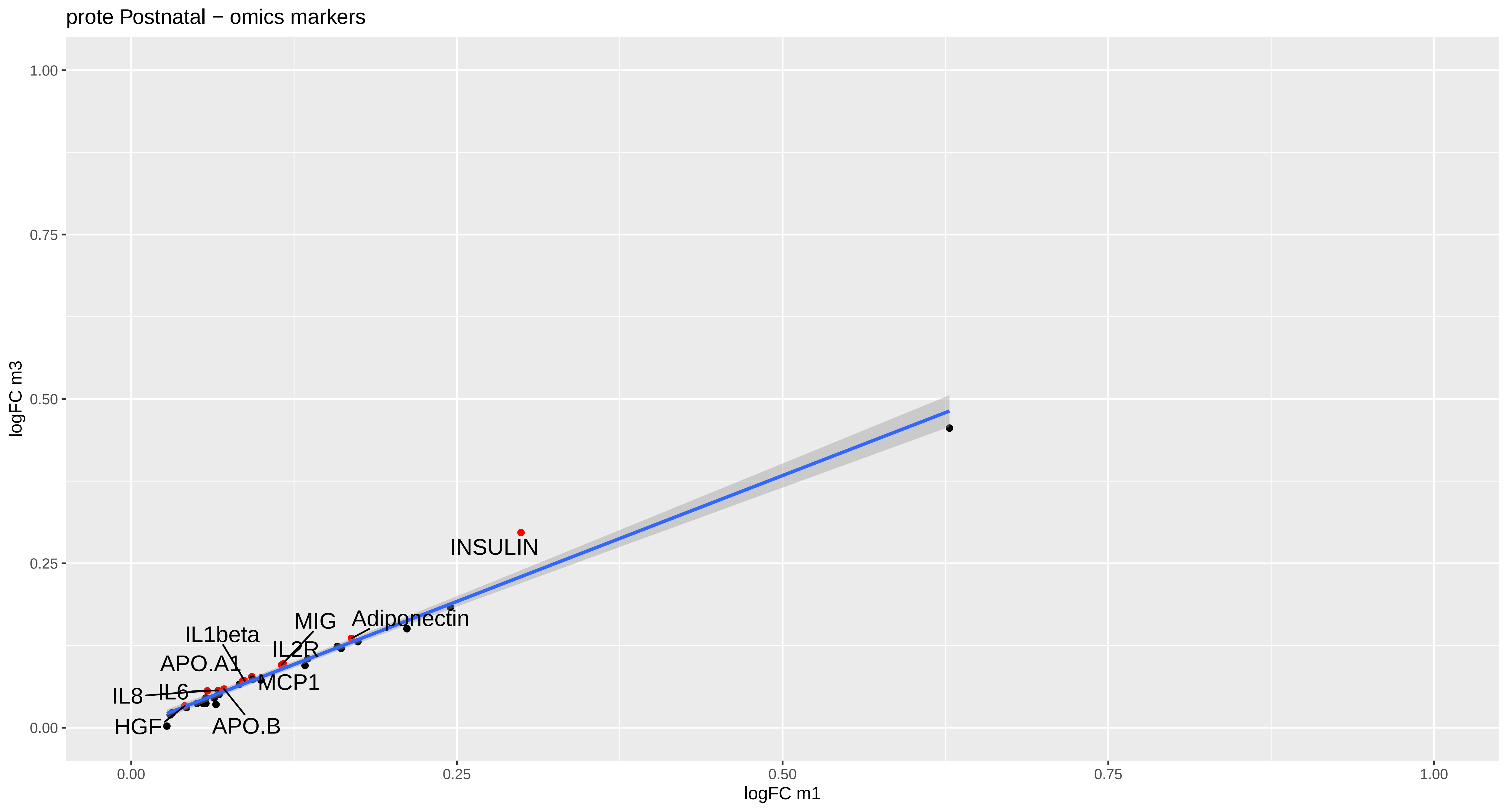

prote Postnatal − omics markers
logFC m3
logFC m1
1.00
0.75
0.50
0.25
0.00
0.00
0.25
0.50
0.75
1.00
INSULIN
MIG
Adiponectin
IL1beta
IL2R
APO.A1
MCP1
IL8
IL6
HGF
APO.B

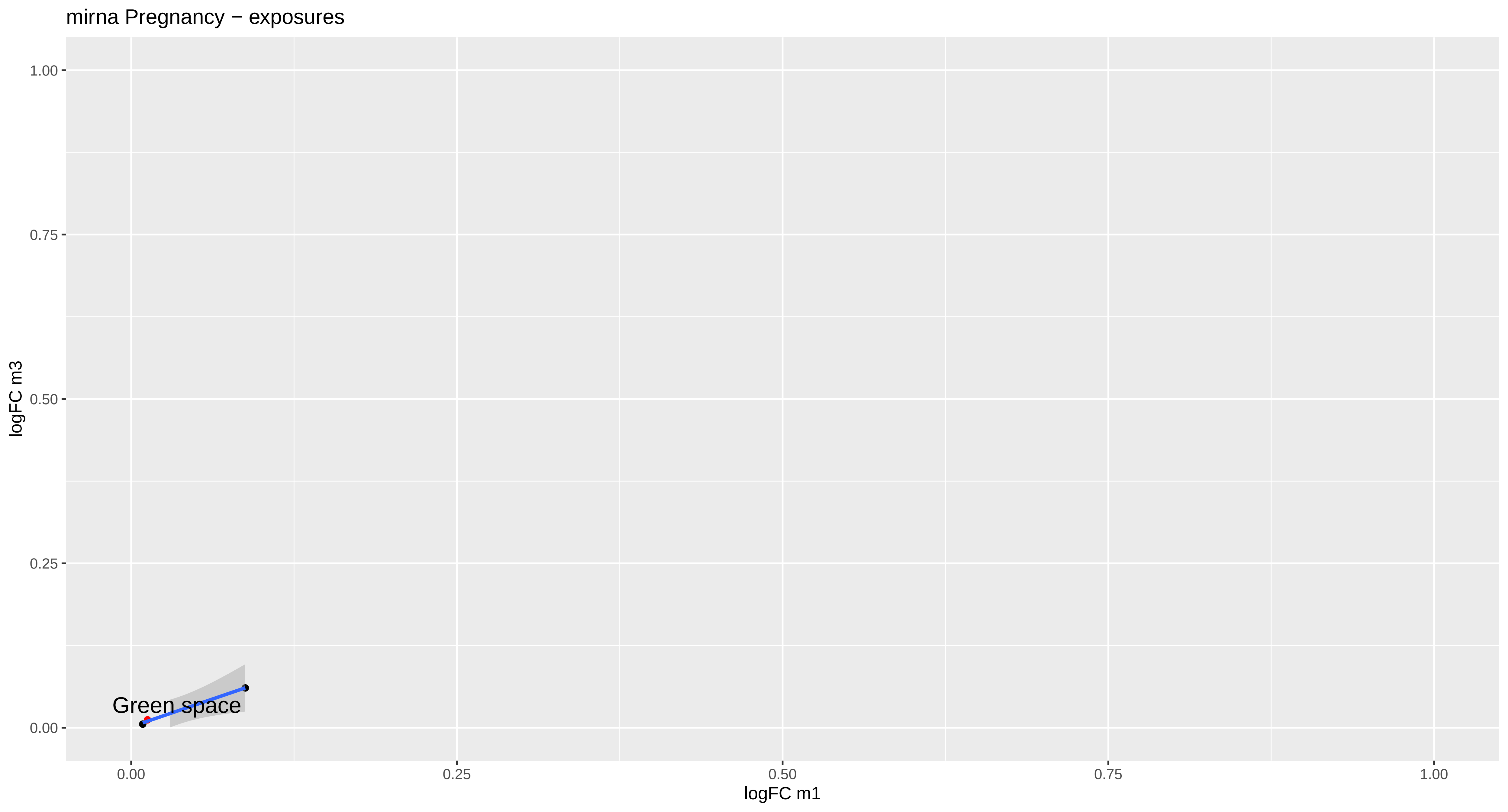

mirna Pregnancy − exposures
1.00
0.75
0.50
0.25
0.00
logFC m3
Green space
0.00
0.25
0.50
0.75
1.00
logFC m1

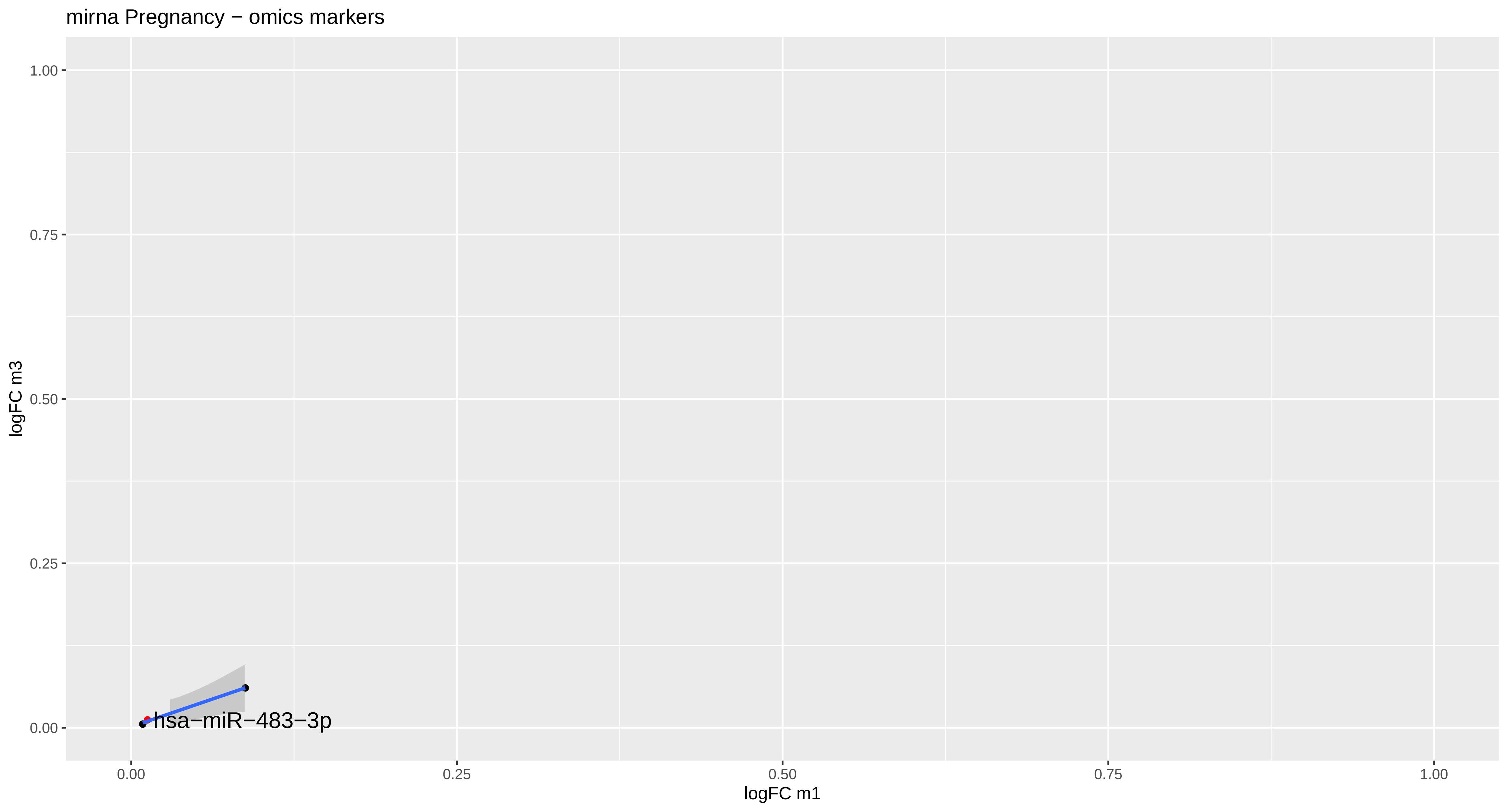
mirna Pregnancy − omics markers
logFC m3
1.00
0.75
0.50
0.25
0.00
hsa−miR−483−3p
0.00
0.25
0.50
0.75
1.00
logFC m1

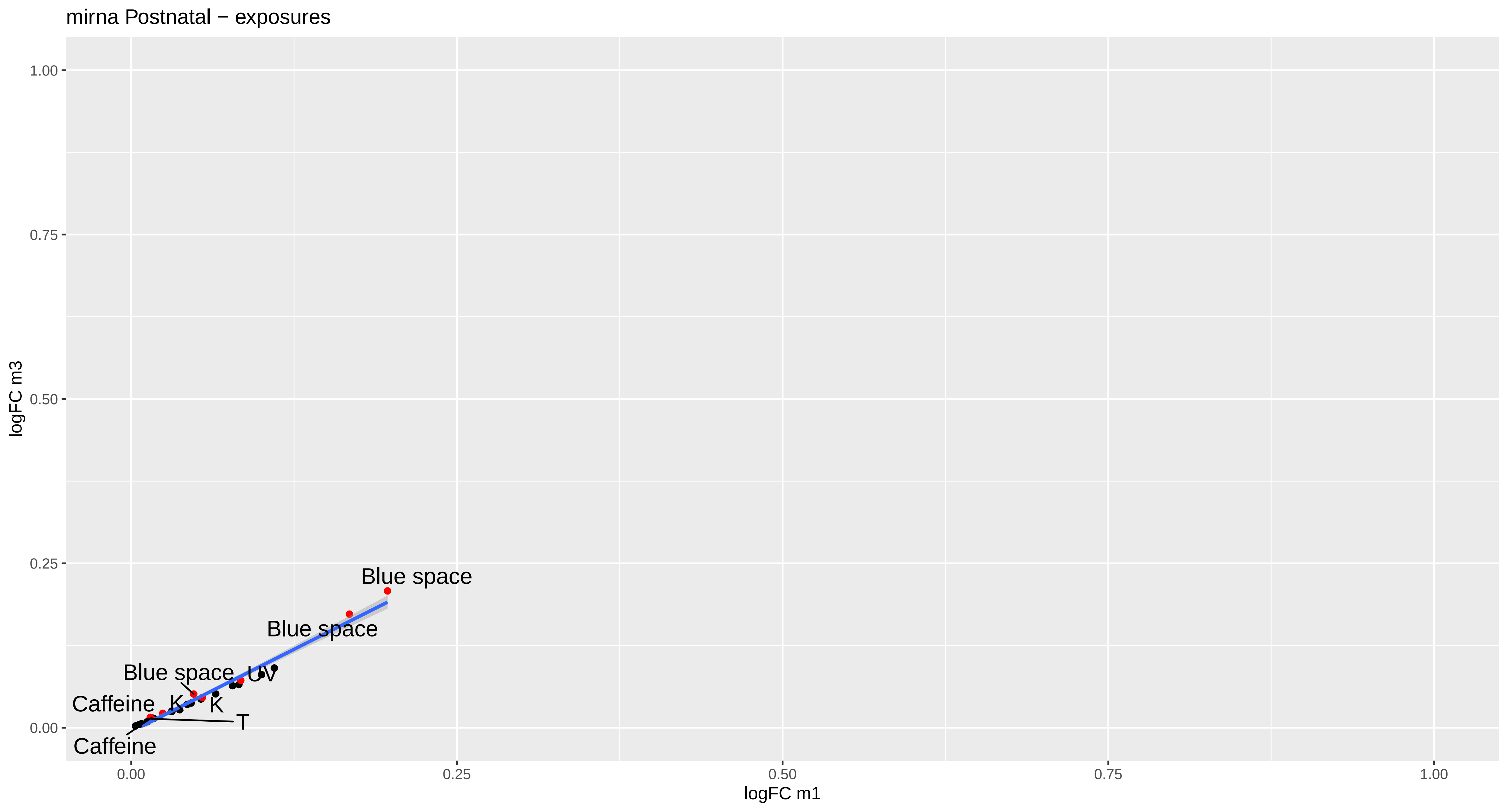

mirna Postnatal − exposures
logFC m3
1.00
0.75
0.50
0.25
0.00
Blue space
Blue space
Blue space
UV
Caffeine
K
K
T
Caffeine
0.00
0.25
0.50
0.75
1.00
logFC m1

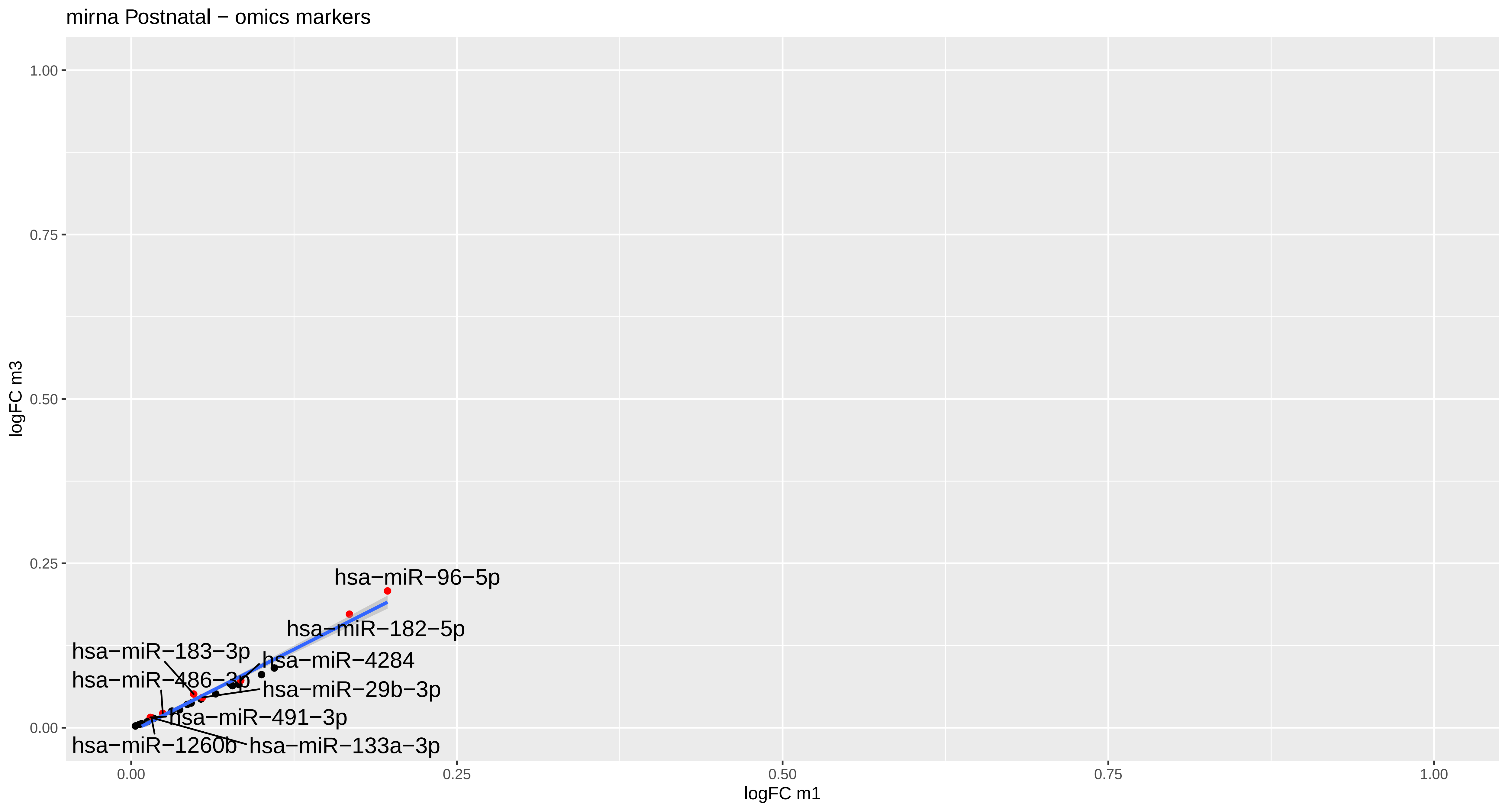

mirna Postnatal − omics markers
hsa−miR−96−5p
hsa−miR−182−5p
hsa−miR−183−3p
hsa−miR−4284
hsa−miR−486−3p
hsa−miR−29b−3p
hsa−miR−491−3p
hsa−miR−1260b
hsa−miR−133a−3p
logFC m3
logFC m1
0.00
0.25
0.50
0.75
1.00

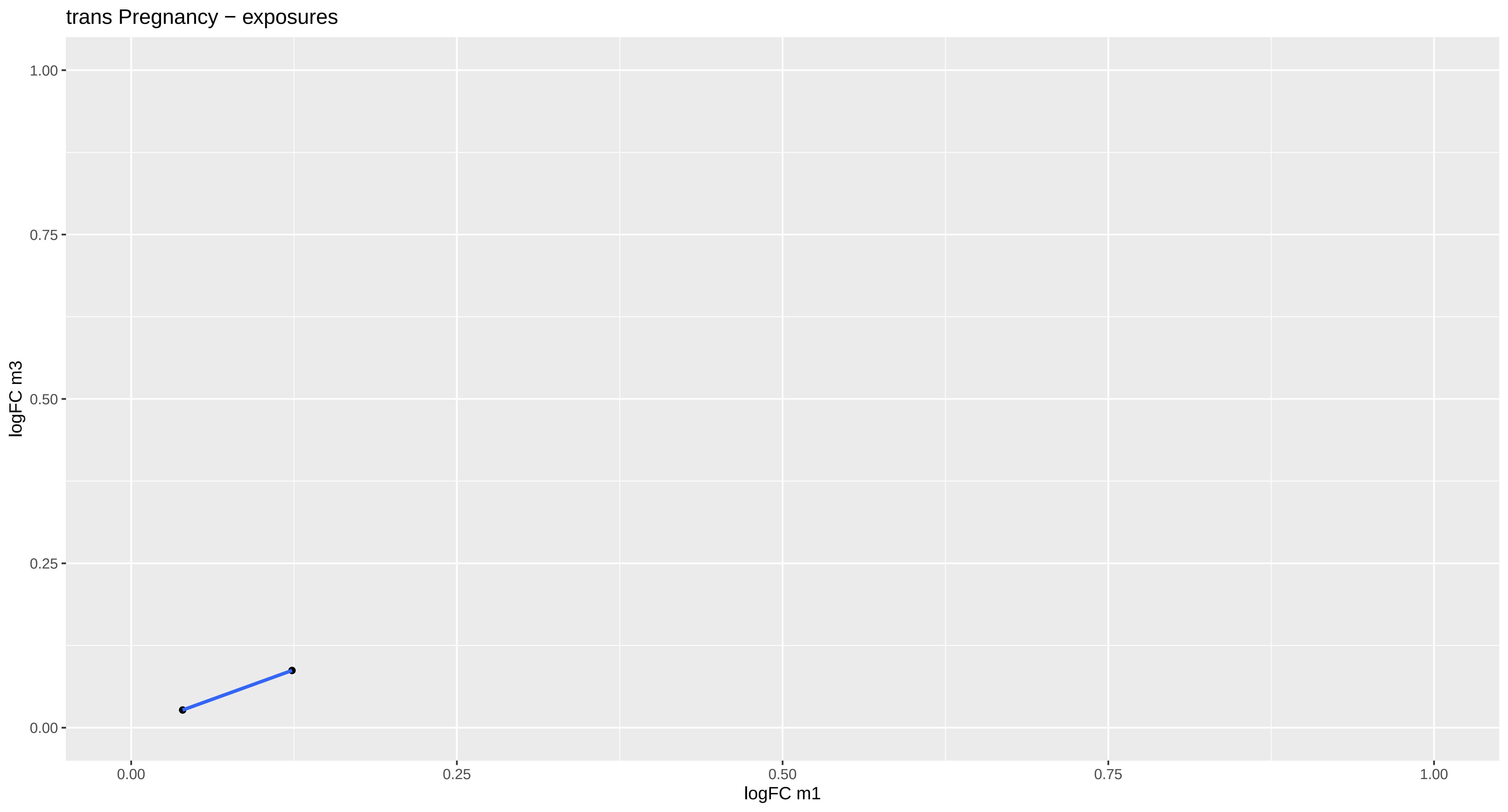

trans Pregnancy − exposures
1.00
0.75
0.50
0.25
0.00
logFC m3
0.00
0.25
0.50
0.75
1.00
logFC m1

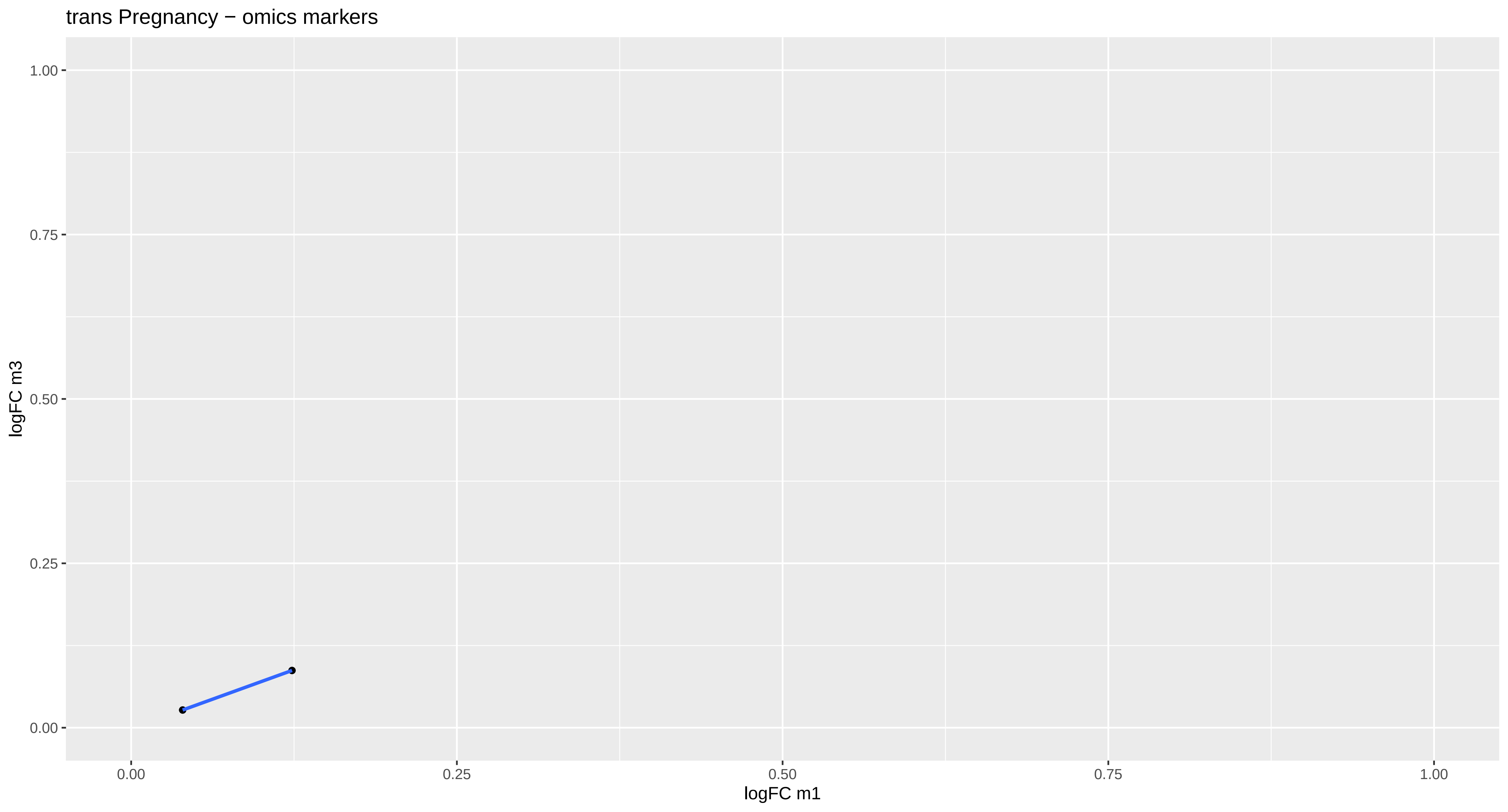

trans Pregnancy − omics markers
logFC m3
logFC m1
1.00
0.75
0.50
0.25
0.00
0.00
0.25
0.50
0.75
1.00

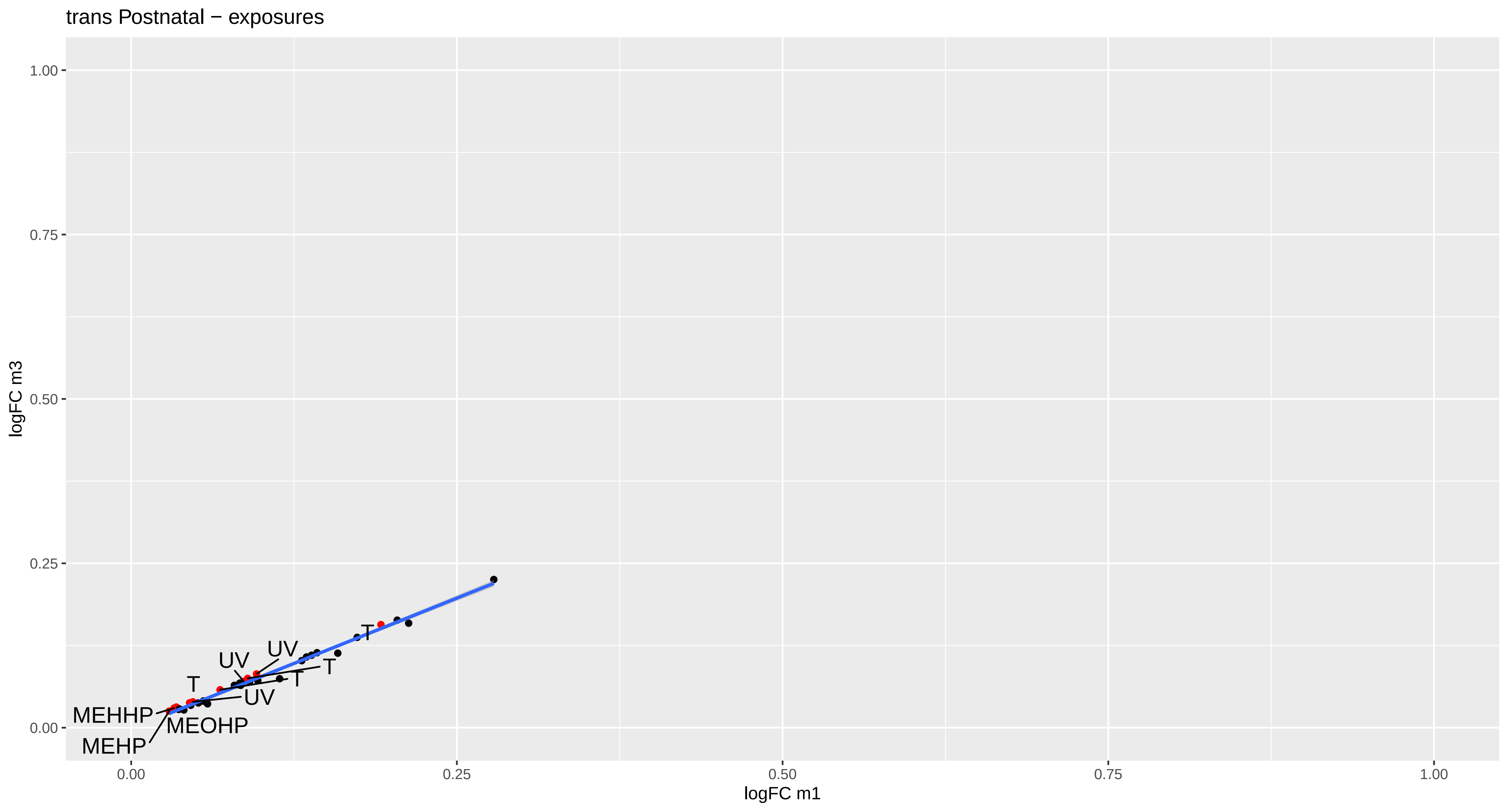
trans Postnatal − exposures
logFC m3
logFC m1
1.00
0.75
0.50
0.25
0.00
0.00
0.25
0.50
0.75
1.00
T
UV
UV
T
T
T
UV
MEHHP
MEOHP
MEHP

trans Postnatal − omics markers

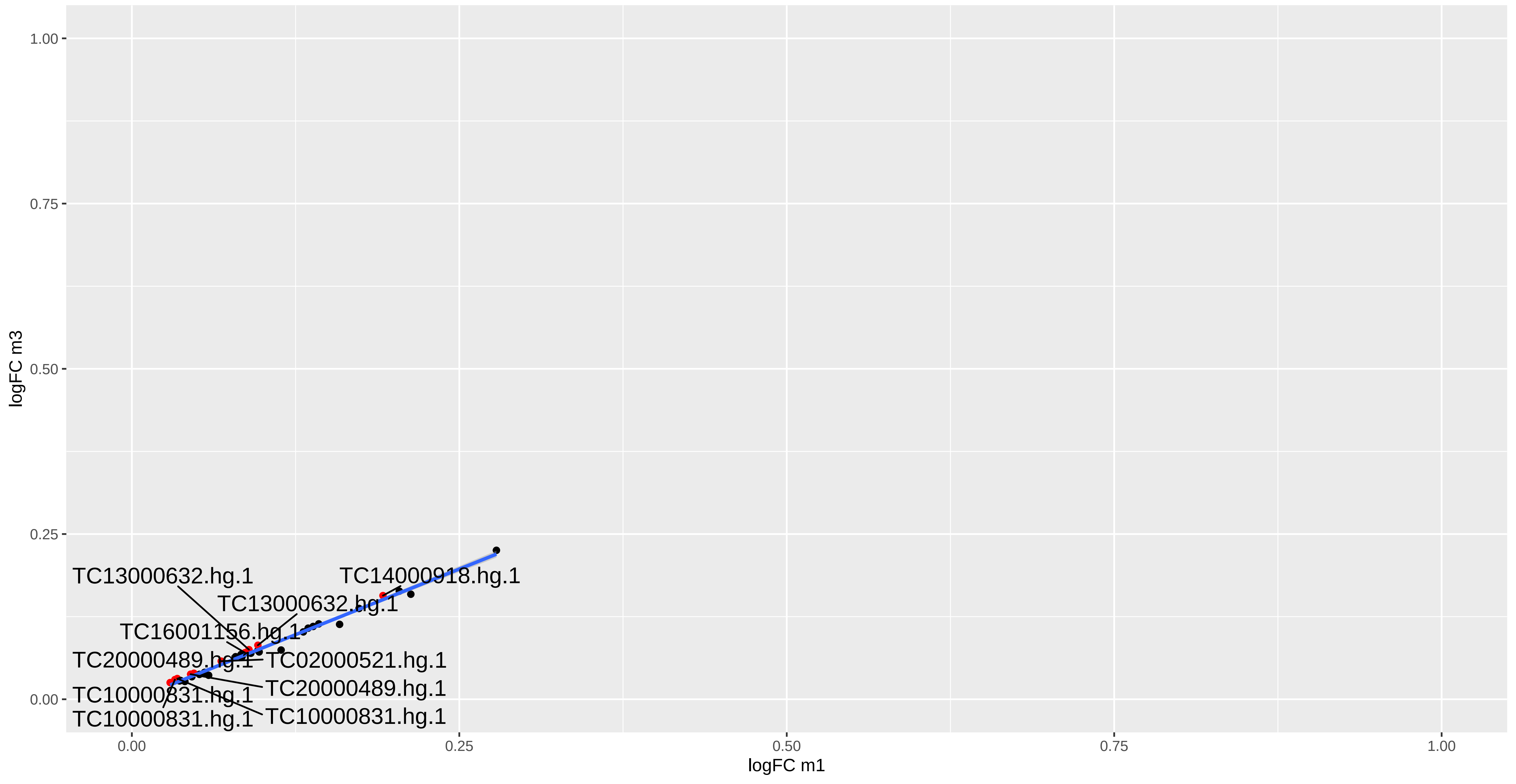

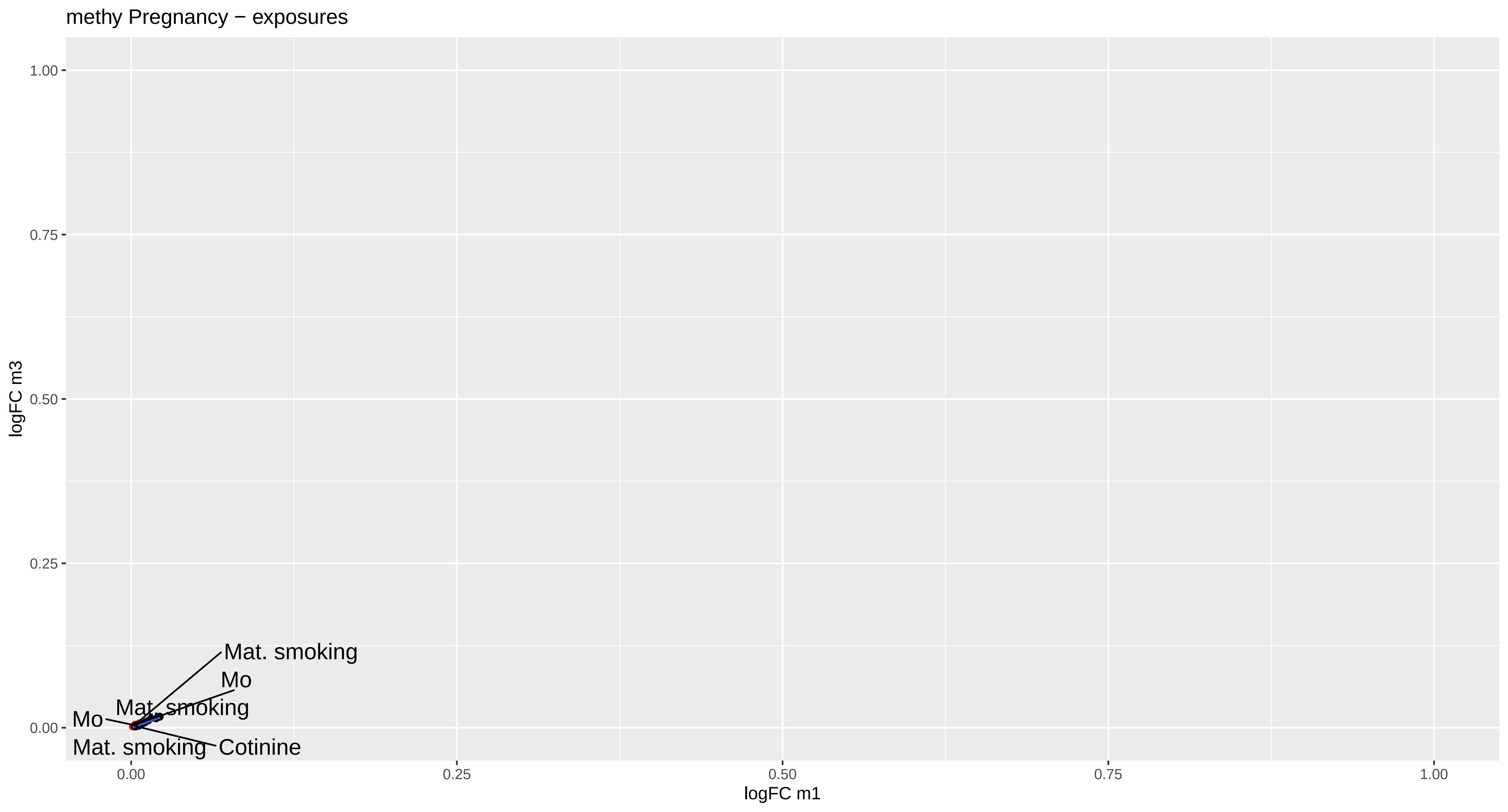
methy Pregnancy − exposures
logFC m3
1.00
0.75
0.50
0.25
0.00
logFC m1
0.00
0.25
0.50
0.75
1.00
Mat. smoking
Mo
Mat. smoking
Mo
Mat. smoking
Cotinine

methy Pregnancy − omics markers

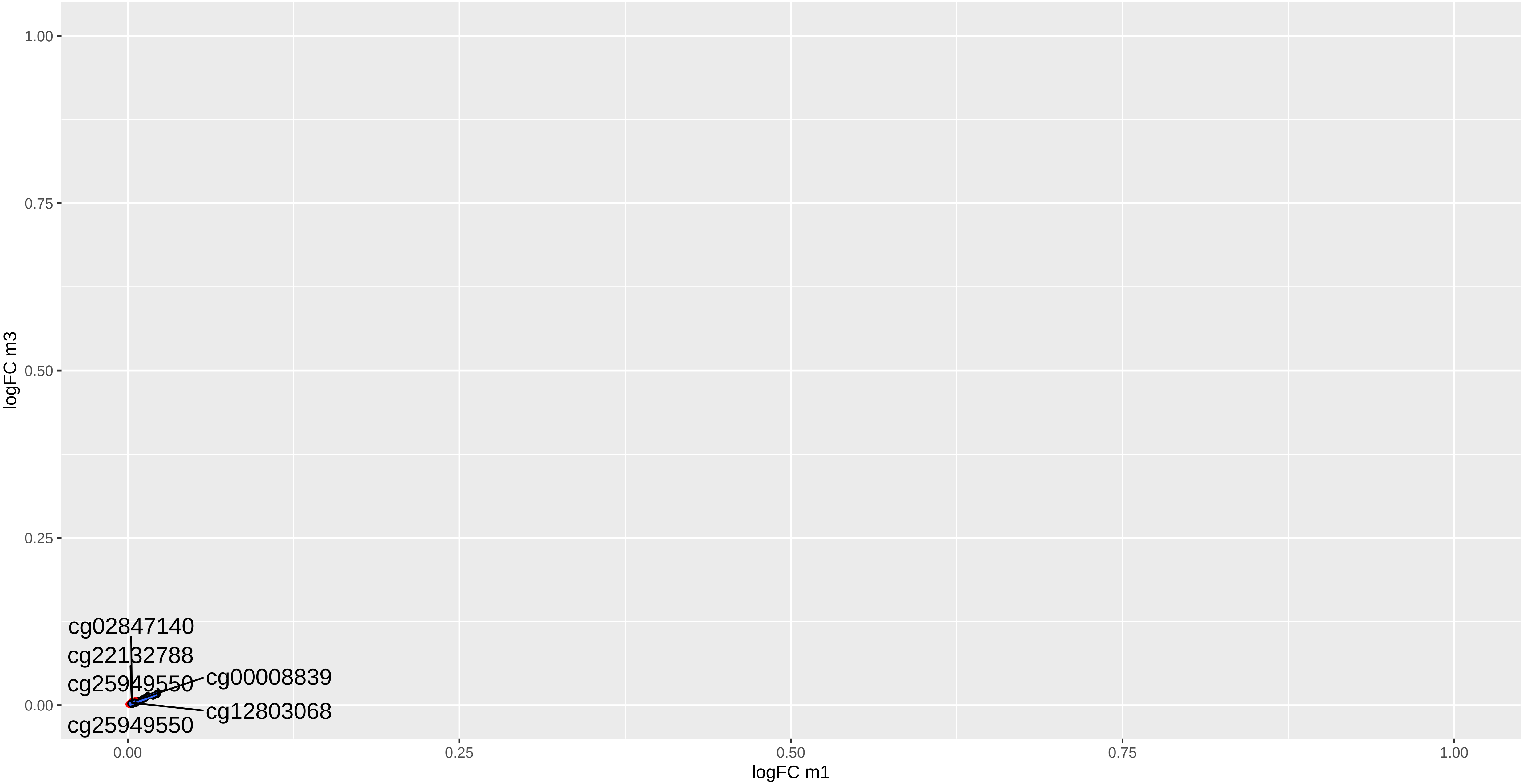

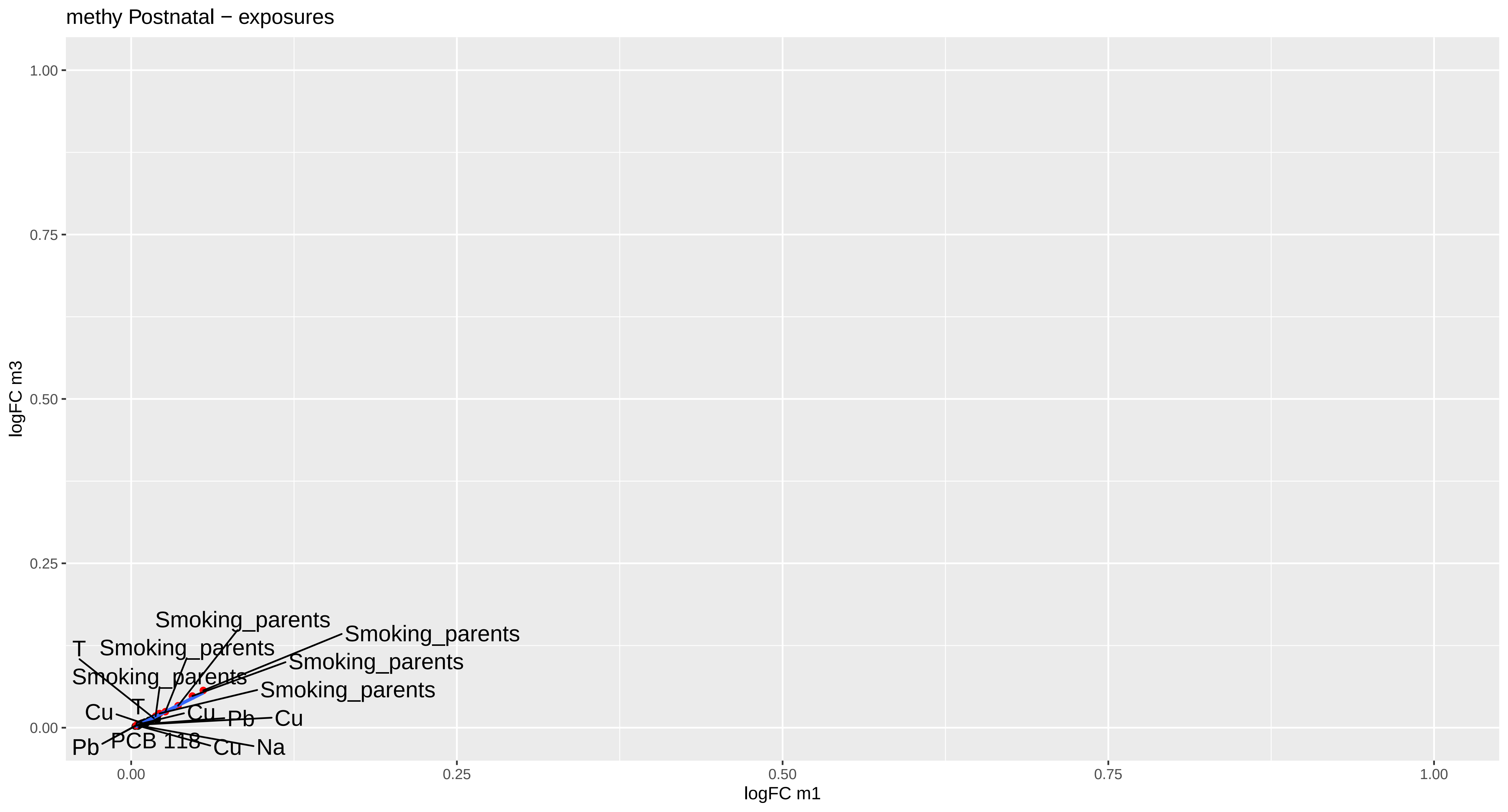

methy Postnatal − exposures
logFC m3
logFC m1
1.00
0.75
0.50
0.25
0.00
0.00
0.25
0.50
0.75
1.00
Smoking_parents
Smoking_parents
T
Smoking_parents
Smoking_parents
Smoking_parents
Smoking_parents
T
Cu
Cu
Pb
Cu
Pb
PCB 118
Cu
Na

methy Postnatal − omics markers

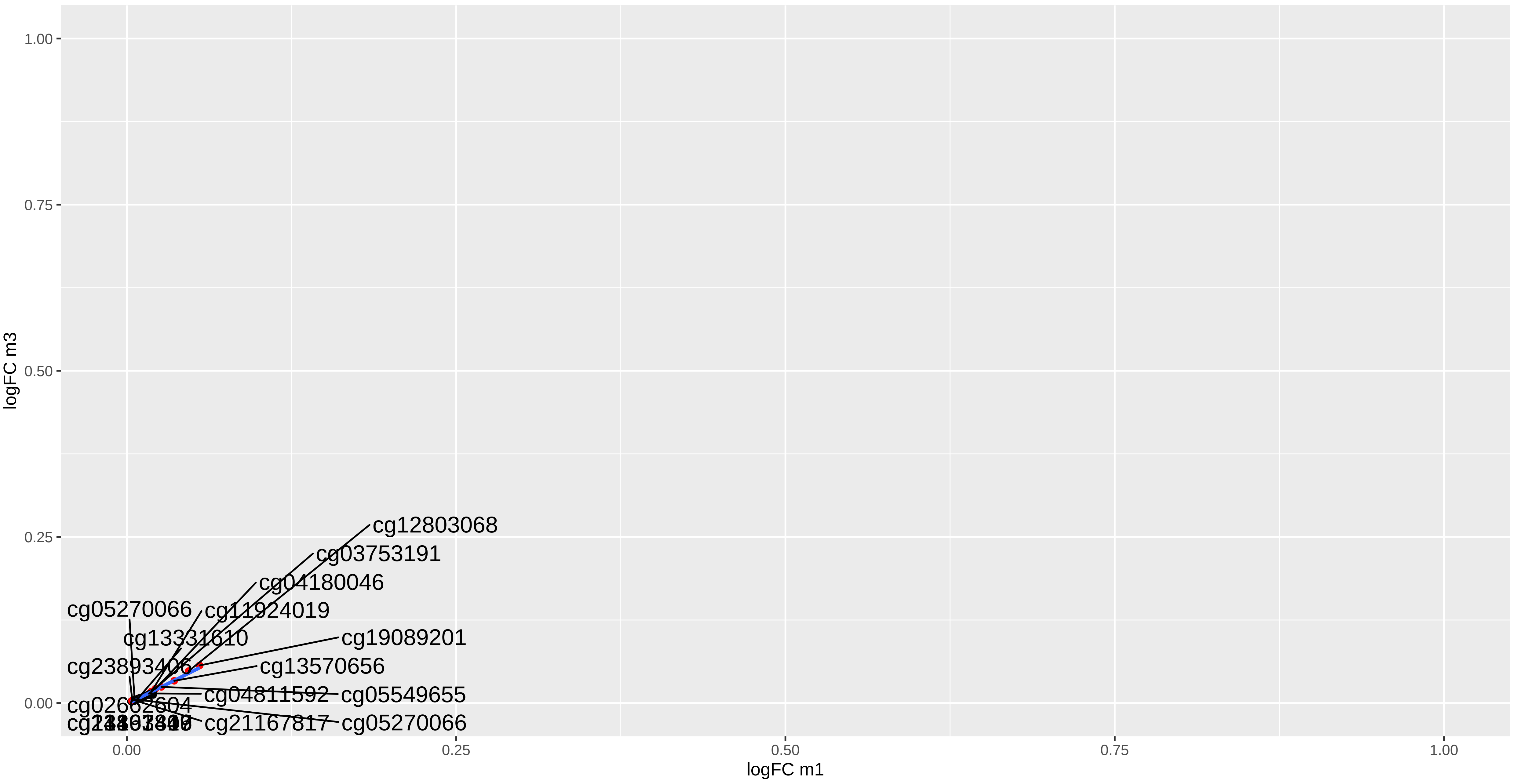
