## Supplementary figures and images for "Multi-omics signatures of the human early life exposome"

### Figure S1

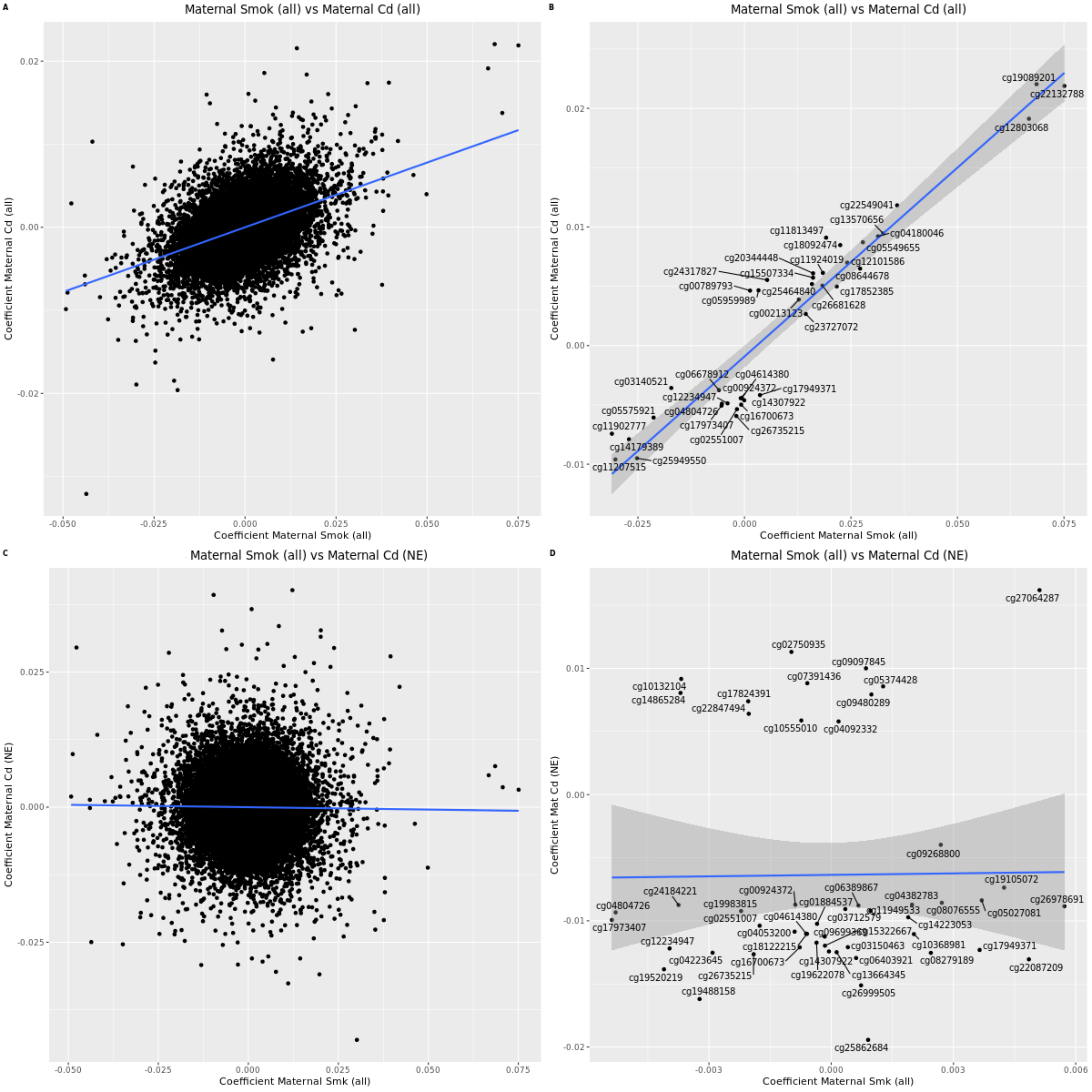

### Figure S4

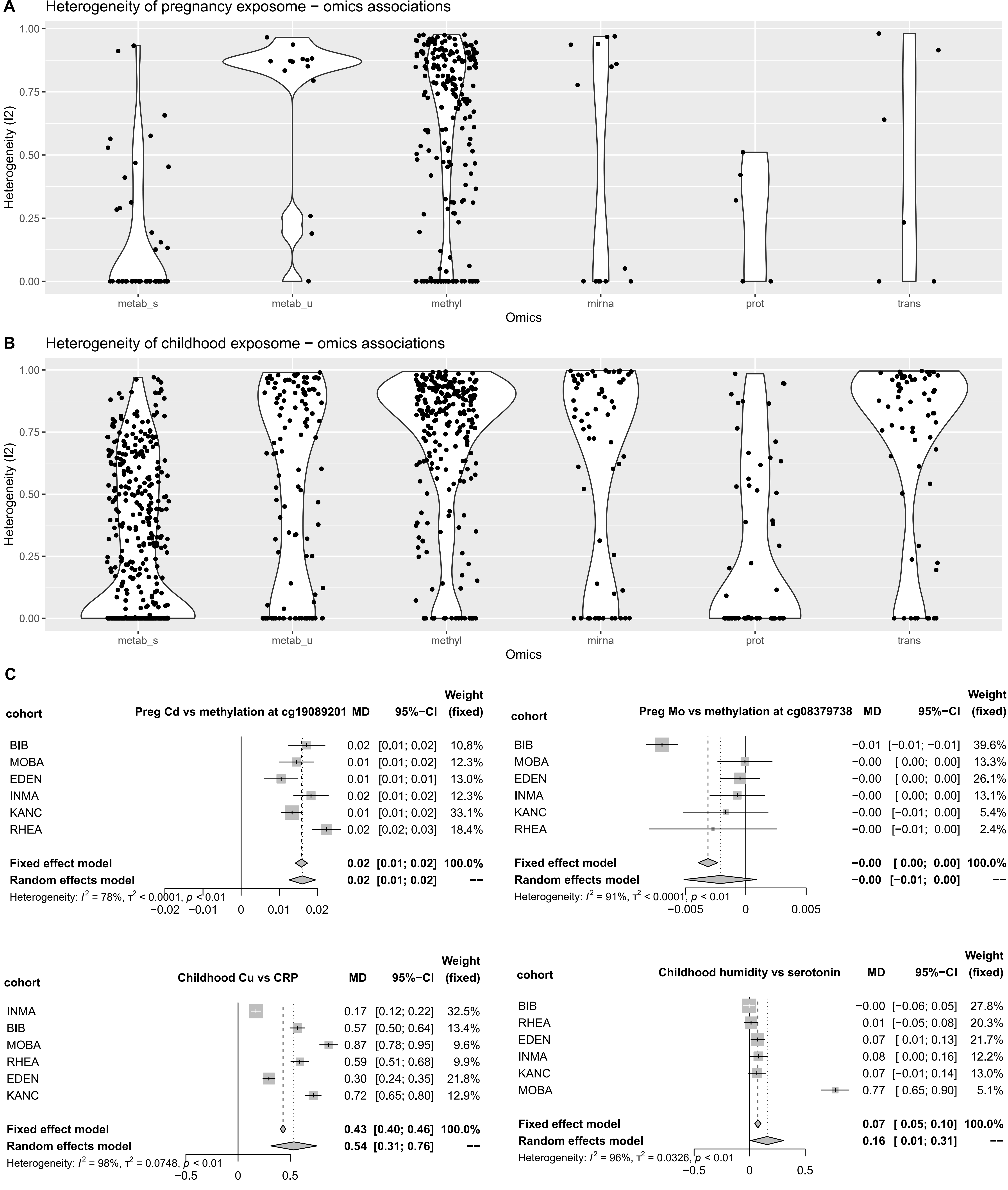
